## Supplemental materials for "Genomic epidemiology of COVID-19 in care homes in the East of England"

1. University of Cambridge, Department of Medicine, Cambridge, UK
2. Cambridge University Hospitals NHS Foundation Trust, Departments of Infectious Diseases and Microbiology, Cambridge UK
3. Wellcome Sanger Institute, Hinxton, UK
4. Cambridgeshire County Council, UK
5. Public Health England, Colindale, UK
6. University of Cambridge, Department of Pathology, Division of Virology, Cambridge, UK
7. Public Health England Clinical Microbiology and Public Health Laboratory, Cambridge UK
8. The Francis Crick Institute, London, UK
9. Department of Haematology, Hammersmith Hospital, Imperial College Healthcare NHS Trust, London, UK
10. [www.cogconsortium.uk](http://www.cogconsortium.uk)

**Correspondence**

William Hamilton

M. Estée Török

### Breakdown of organisations submitting samples to the Cambridge Clinical Microbiology and Public Health Laboratory over the study period

| **Submitting site** | **Number of positive cases** |
| --- | --- |
| Southend University Hospital | 888 |
| Luton and Dunstable University Hospital | 691 |
| Bedford Hospital | 668 |
| Watford General Hospital | 633 |
| PHE East of England HPT | 513 |
| Addenbrooke's Hospital | 512 |
| Lister Hospital | 506 |
| Colchester Hospital | 441 |
| Ipswich Hospital | 420 |
| Broomfield Hospital | 327 |
| West Suffolk Hospital | 199 |
| Basildon University Hospital | 196 |
| Herts Urgent Care Ltd | 97 |
| Princess Alexandra Hospital | 86 |
| Royal Papworth Hospital | 71 |
| Hinchingbrooke Hospital | 67 |
| Commisceo Primary Care Solutions | 63 |
| Peterborough City Hospital | 58 |
| Milton Keynes Hospital | 53 |
| EEAST COVID-19 Coordination Centre | 32 |
| PHL London | 19 |
| Norfolk and Norwich University Hospital | 17 |
| Unknown | 11 |
| Other | 32 |
| **Total** | **6600** |

To maintain patient anonymity, sites that submitted samples from ≤10 people with positive test results are not shown and are counted as “Other”. Note that over the course of the study, some sites changed testing provider from CMPHL as further testing sites became available around the region. This explains some of the variation in the relative proportion of cases submitted from each site. The numbers reported here do not necessarily reflect total case numbers for each hospital or submitting organisation, as tests may have been performed elsewhere or metadata not collected in this study; the numbers are included purely to indicate where the samples included in this study originated from. CMPHL = Clinical Microbiology and Public Health Laboratory.

### Demonstration of *transcluster* algorithm with anonymised data

Code to reproduce the *transcluster* transmission analysis using anonymised metadata is available via GitHub at: <https://github.com/gtonkinhill/SC2-care-homes-anonymised>. The genomes are the same as those used in the study, but sample names in the multiple sequence alignment and corresponding metadata have been changed from COG-UK sequence codes to anonymised sample codes.

Sequences generated through the COG-UK consortium have associated public metadata (available via the COG-UK website or GISAID), including patient age, sex, collection date (if available), and location to the level of UK county. COG-UK samples are sequenced under statutory powers granted to the UK Public Health Agencies. Matched patient data is securely released to the COG-UK consortium under a data sharing framework which strictly controls the handling of patient data. The status of individuals living in a care home and groups of such care home patients are both on the consortium restricted data list. This means that this data cannot be publicly released linked to their sequencing identifiers (eg. COG-UK sequence codes). This is because of the risk of deductive disclosure, potentially compromising study participant anonymity. However, the anonymised dataset released on GitHub can be used to fully replicate the *transcluster* analyses from the study without requiring public release of sample COG-UK codes linked to information on care home residency.

If a researcher requires access to restricted metadata (including care home residency status) linked to the COG-UK sequence codes, then this will require a formal data sharing agreement with the COG-UK Consortium. Access to patient outcome information for patients treated at Cambridge University Hospitals NHS Foundation Trust (CUH) requires a data sharing agreement with CUH. Data will only be shared for public health and research purposes, not for commercial enterprise, and only to individuals working at reputable research and public health institutions for which data security can be assured. Should this be required researchers should contact the study corresponding authors in the first instance.
Sampling date ranges for care home residents with genomic data: by care home

Table shows anonymised care home codes for the 292 care homes from 700 care home residents with genomic data. “Number of residents” shows the number of residents with genomic data for each care home. The date of the first and last samples collected from each care home and the date difference in days between these samples is shown. If there was only 1 sample from the care home, and/or all cases were from the same date, then “first sample date” and “last sample date” are the same. Collection date was used for sample dates; if collection date was missing then receive date in the laboratory was used instead. 170 / 292 (58%) care homes had 2 or more cases with genomic data, including 578 care home residents. Of these, the median date difference from first to last sampling dates was 9 days (interquartile range: 4 – 15; range: 0 – 50).

| **Care home code** | **Number of residents** | **First sample date** | **Last sample date** | **Date difference (days)** |
| --- | --- | --- | --- | --- |
| CARE0001 | 2 | 11/03/2020 | 17/03/2020 | 6 |
| CARE0002 | 2 | 12/03/2020 | 14/03/2020 | 2 |
| CARE0004 | 2 | 13/03/2020 | 13/03/2020 | 0 |
| CARE0005 | 2 | 16/03/2020 | 30/04/2020 | 45 |
| CARE0006 | 2 | 14/03/2020 | 16/03/2020 | 2 |
| CARE0007 | 1 | 18/03/2020 | 18/03/2020 | 0 |
| CARE0008 | 2 | 16/03/2020 | 05/04/2020 | 20 |
| CARE0010 | 2 | 19/03/2020 | 20/03/2020 | 1 |
| CARE0011 | 5 | 19/03/2020 | 31/03/2020 | 12 |
| CARE0012 | 4 | 20/03/2020 | 05/04/2020 | 16 |
| CARE0014 | 4 | 20/03/2020 | 09/05/2020 | 50 |
| CARE0015 | 2 | 20/03/2020 | 20/04/2020 | 31 |
| CARE0016 | 1 | 22/03/2020 | 22/03/2020 | 0 |
| CARE0017 | 2 | 21/03/2020 | 05/04/2020 | 15 |
| CARE0018 | 2 | 21/03/2020 | 06/04/2020 | 16 |
| CARE0019 | 1 | 21/03/2020 | 21/03/2020 | 0 |
| CARE0020 | 2 | 21/03/2020 | 31/03/2020 | 10 |
| CARE0021 | 2 | 21/03/2020 | 30/03/2020 | 9 |
| CARE0022 | 1 | 23/03/2020 | 23/03/2020 | 0 |
| CARE0023 | 2 | 22/03/2020 | 07/04/2020 | 16 |
| CARE0024 | 3 | 22/03/2020 | 20/04/2020 | 29 |
| CARE0025 | 2 | 23/03/2020 | 03/04/2020 | 11 |
| CARE0026 | 3 | 23/03/2020 | 02/04/2020 | 10 |
| CARE0027 | 1 | 02/04/2020 | 02/04/2020 | 0 |
| CARE0028 | 6 | 29/03/2020 | 06/04/2020 | 8 |
| CARE0029 | 4 | 05/04/2020 | 13/04/2020 | 8 |
| CARE0030 | 2 | 26/03/2020 | 27/04/2020 | 32 |
| CARE0031 | 1 | 09/04/2020 | 09/04/2020 | 0 |
| CARE0032 | 7 | 01/04/2020 | 10/05/2020 | 39 |
| CARE0033 | 1 | 31/03/2020 | 31/03/2020 | 0 |
| CARE0034 | 1 | 26/03/2020 | 26/03/2020 | 0 |
| CARE0036 | 6 | 03/04/2020 | 01/05/2020 | 28 |
| CARE0037 | 1 | 30/03/2020 | 30/03/2020 | 0 |
| CARE0038 | 2 | 04/04/2020 | 06/04/2020 | 2 |
| CARE0039 | 1 | 06/05/2020 | 06/05/2020 | 0 |
| CARE0040 | 1 | 27/03/2020 | 27/03/2020 | 0 |
| CARE0042 | 1 | 10/04/2020 | 10/04/2020 | 0 |
| CARE0043 | 3 | 09/04/2020 | 11/04/2020 | 2 |
| CARE0046 | 2 | 18/04/2020 | 07/05/2020 | 19 |
| CARE0047 | 5 | 02/04/2020 | 07/05/2020 | 35 |
| CARE0048 | 1 | 27/03/2020 | 27/03/2020 | 0 |
| CARE0049 | 1 | 31/03/2020 | 31/03/2020 | 0 |
| CARE0051 | 2 | 10/04/2020 | 21/04/2020 | 11 |
| CARE0054 | 4 | 05/04/2020 | 02/05/2020 | 27 |
| CARE0055 | 1 | 31/03/2020 | 31/03/2020 | 0 |
| CARE0056 | 1 | 29/03/2020 | 29/03/2020 | 0 |
| CARE0057 | 5 | 29/03/2020 | 12/04/2020 | 14 |
| CARE0058 | 2 | 28/03/2020 | 27/04/2020 | 30 |
| CARE0059 | 1 | 28/03/2020 | 28/03/2020 | 0 |
| CARE0060 | 2 | 29/03/2020 | 21/04/2020 | 23 |
| CARE0061 | 10 | 29/03/2020 | 06/05/2020 | 38 |
| CARE0062 | 2 | 30/03/2020 | 29/04/2020 | 30 |
| CARE0063 | 12 | 30/03/2020 | 20/04/2020 | 21 |
| CARE0064 | 1 | 29/03/2020 | 29/03/2020 | 0 |
| CARE0065 | 1 | 29/03/2020 | 29/03/2020 | 0 |
| CARE0066 | 6 | 29/03/2020 | 08/04/2020 | 10 |
| CARE0067 | 1 | 31/03/2020 | 31/03/2020 | 0 |
| CARE0068 | 4 | 31/03/2020 | 09/05/2020 | 39 |
| CARE0069 | 3 | 30/03/2020 | 07/04/2020 | 8 |
| CARE0070 | 1 | 29/03/2020 | 29/03/2020 | 0 |
| CARE0071 | 1 | 30/03/2020 | 30/03/2020 | 0 |
| CARE0073 | 3 | 30/03/2020 | 20/04/2020 | 21 |
| CARE0074 | 2 | 31/03/2020 | 08/04/2020 | 8 |
| CARE0075 | 2 | 30/03/2020 | 06/04/2020 | 7 |
| CARE0076 | 4 | 30/03/2020 | 12/04/2020 | 13 |
| CARE0077 | 1 | 31/03/2020 | 31/03/2020 | 0 |
| CARE0078 | 2 | 12/04/2020 | 06/05/2020 | 24 |
| CARE0079 | 2 | 12/04/2020 | 13/04/2020 | 1 |
| CARE0080 | 3 | 31/03/2020 | 04/04/2020 | 4 |
| CARE0081 | 2 | 31/03/2020 | 11/04/2020 | 11 |
| CARE0082 | 2 | 31/03/2020 | 21/04/2020 | 21 |
| CARE0083 | 2 | 31/03/2020 | 06/05/2020 | 36 |
| CARE0084 | 2 | 01/04/2020 | 10/04/2020 | 9 |
| CARE0085 | 1 | 31/03/2020 | 31/03/2020 | 0 |
| CARE0086 | 1 | 31/03/2020 | 31/03/2020 | 0 |
| CARE0087 | 1 | 30/03/2020 | 30/03/2020 | 0 |
| CARE0088 | 1 | 31/03/2020 | 31/03/2020 | 0 |
| CARE0089 | 1 | 02/04/2020 | 02/04/2020 | 0 |
| CARE0090 | 1 | 01/04/2020 | 01/04/2020 | 0 |
| CARE0091 | 5 | 02/04/2020 | 16/04/2020 | 14 |
| CARE0092 | 5 | 01/04/2020 | 22/04/2020 | 21 |
| CARE0093 | 4 | 01/04/2020 | 14/04/2020 | 13 |
| CARE0094 | 2 | 02/04/2020 | 16/04/2020 | 14 |
| CARE0095 | 2 | 01/04/2020 | 08/04/2020 | 7 |
| CARE0096 | 2 | 02/04/2020 | 13/04/2020 | 11 |
| CARE0097 | 7 | 03/04/2020 | 01/05/2020 | 28 |
| CARE0098 | 3 | 02/04/2020 | 06/04/2020 | 4 |
| CARE0099 | 2 | 03/04/2020 | 16/04/2020 | 13 |
| CARE0100 | 2 | 01/04/2020 | 02/04/2020 | 1 |
| CARE0101 | 2 | 02/04/2020 | 06/04/2020 | 4 |
| CARE0102 | 5 | 02/04/2020 | 04/05/2020 | 32 |
| CARE0103 | 2 | 08/04/2020 | 21/04/2020 | 13 |
| CARE0104 | 3 | 02/04/2020 | 10/04/2020 | 8 |
| CARE0105 | 2 | 03/04/2020 | 03/04/2020 | 0 |
| CARE0106 | 3 | 02/04/2020 | 12/04/2020 | 10 |
| CARE0107 | 1 | 03/04/2020 | 03/04/2020 | 0 |
| CARE0108 | 2 | 03/04/2020 | 29/04/2020 | 26 |
| CARE0109 | 2 | 04/04/2020 | 07/04/2020 | 3 |
| CARE0111 | 3 | 03/04/2020 | 07/04/2020 | 4 |
| CARE0112 | 1 | 03/04/2020 | 03/04/2020 | 0 |
| CARE0113 | 2 | 03/04/2020 | 12/04/2020 | 9 |
| CARE0114 | 3 | 07/04/2020 | 15/04/2020 | 8 |
| CARE0116 | 2 | 04/04/2020 | 10/04/2020 | 6 |
| CARE0117 | 2 | 04/04/2020 | 13/04/2020 | 9 |
| CARE0118 | 3 | 06/04/2020 | 18/04/2020 | 12 |
| CARE0119 | 3 | 03/04/2020 | 06/04/2020 | 3 |
| CARE0120 | 4 | 03/04/2020 | 01/05/2020 | 28 |
| CARE0121 | 1 | 03/04/2020 | 03/04/2020 | 0 |
| CARE0122 | 6 | 04/04/2020 | 18/04/2020 | 14 |
| CARE0123 | 1 | 05/04/2020 | 05/04/2020 | 0 |
| CARE0124 | 2 | 05/04/2020 | 11/04/2020 | 6 |
| CARE0125 | 6 | 05/04/2020 | 17/04/2020 | 12 |
| CARE0126 | 3 | 05/04/2020 | 14/04/2020 | 9 |
| CARE0127 | 2 | 04/04/2020 | 18/04/2020 | 14 |
| CARE0128 | 4 | 05/04/2020 | 11/04/2020 | 6 |
| CARE0129 | 1 | 05/04/2020 | 05/04/2020 | 0 |
| CARE0130 | 1 | 05/04/2020 | 05/04/2020 | 0 |
| CARE0131 | 1 | 04/04/2020 | 04/04/2020 | 0 |
| CARE0132 | 1 | 05/04/2020 | 05/04/2020 | 0 |
| CARE0133 | 1 | 04/04/2020 | 04/04/2020 | 0 |
| CARE0134 | 4 | 06/04/2020 | 19/04/2020 | 13 |
| CARE0135 | 2 | 05/04/2020 | 08/04/2020 | 3 |
| CARE0136 | 1 | 05/04/2020 | 05/04/2020 | 0 |
| CARE0137 | 1 | 05/04/2020 | 05/04/2020 | 0 |
| CARE0138 | 1 | 05/04/2020 | 05/04/2020 | 0 |
| CARE0139 | 4 | 05/04/2020 | 15/04/2020 | 10 |
| CARE0140 | 1 | 06/04/2020 | 06/04/2020 | 0 |
| CARE0141 | 1 | 07/04/2020 | 07/04/2020 | 0 |
| CARE0142 | 3 | 04/04/2020 | 19/04/2020 | 15 |
| CARE0143 | 1 | 05/04/2020 | 05/04/2020 | 0 |
| CARE0144 | 2 | 05/04/2020 | 07/04/2020 | 2 |
| CARE0145 | 2 | 07/04/2020 | 30/04/2020 | 23 |
| CARE0146 | 2 | 07/04/2020 | 09/04/2020 | 2 |
| CARE0147 | 1 | 07/04/2020 | 07/04/2020 | 0 |
| CARE0148 | 2 | 07/04/2020 | 07/04/2020 | 0 |
| CARE0149 | 5 | 07/04/2020 | 07/05/2020 | 30 |
| CARE0150 | 2 | 07/04/2020 | 09/04/2020 | 2 |
| CARE0151 | 7 | 07/04/2020 | 27/04/2020 | 20 |
| CARE0153 | 1 | 08/04/2020 | 08/04/2020 | 0 |
| CARE0154 | 1 | 08/04/2020 | 08/04/2020 | 0 |
| CARE0155 | 2 | 08/04/2020 | 21/04/2020 | 13 |
| CARE0156 | 3 | 07/04/2020 | 24/04/2020 | 17 |
| CARE0157 | 5 | 08/04/2020 | 26/04/2020 | 18 |
| CARE0158 | 1 | 08/04/2020 | 08/04/2020 | 0 |
| CARE0159 | 6 | 09/04/2020 | 06/05/2020 | 27 |
| CARE0160 | 1 | 09/04/2020 | 09/04/2020 | 0 |
| CARE0161 | 3 | 08/04/2020 | 14/04/2020 | 6 |
| CARE0162 | 4 | 09/04/2020 | 01/05/2020 | 22 |
| CARE0165 | 1 | 07/05/2020 | 07/05/2020 | 0 |
| CARE0166 | 4 | 11/04/2020 | 24/04/2020 | 13 |
| CARE0167 | 1 | 13/04/2020 | 13/04/2020 | 0 |
| CARE0169 | 4 | 18/04/2020 | 30/04/2020 | 12 |
| CARE0170 | 1 | 09/04/2020 | 09/04/2020 | 0 |
| CARE0171 | 2 | 09/04/2020 | 16/04/2020 | 7 |
| CARE0172 | 2 | 09/04/2020 | 13/04/2020 | 4 |
| CARE0173 | 7 | 10/04/2020 | 01/05/2020 | 21 |
| CARE0174 | 4 | 09/04/2020 | 17/04/2020 | 8 |
| CARE0175 | 2 | 09/04/2020 | 24/04/2020 | 15 |
| CARE0176 | 2 | 10/04/2020 | 18/04/2020 | 8 |
| CARE0177 | 3 | 10/04/2020 | 01/05/2020 | 21 |
| CARE0178 | 1 | 11/04/2020 | 11/04/2020 | 0 |
| CARE0179 | 1 | 10/04/2020 | 10/04/2020 | 0 |
| CARE0180 | 1 | 10/04/2020 | 10/04/2020 | 0 |
| CARE0181 | 2 | 10/04/2020 | 14/04/2020 | 4 |
| CARE0182 | 5 | 11/04/2020 | 21/04/2020 | 10 |
| CARE0183 | 1 | 11/04/2020 | 11/04/2020 | 0 |
| CARE0184 | 3 | 12/04/2020 | 17/04/2020 | 5 |
| CARE0185 | 1 | 11/04/2020 | 11/04/2020 | 0 |
| CARE0186 | 1 | 11/04/2020 | 11/04/2020 | 0 |
| CARE0187 | 1 | 12/04/2020 | 12/04/2020 | 0 |
| CARE0188 | 1 | 11/04/2020 | 11/04/2020 | 0 |
| CARE0189 | 4 | 11/04/2020 | 20/04/2020 | 9 |
| CARE0190 | 2 | 12/04/2020 | 21/04/2020 | 9 |
| CARE0191 | 2 | 11/04/2020 | 14/04/2020 | 3 |
| CARE0192 | 1 | 11/04/2020 | 11/04/2020 | 0 |
| CARE0193 | 1 | 11/04/2020 | 11/04/2020 | 0 |
| CARE0194 | 3 | 12/04/2020 | 28/04/2020 | 16 |
| CARE0195 | 1 | 13/04/2020 | 13/04/2020 | 0 |
| CARE0196 | 5 | 13/04/2020 | 28/04/2020 | 15 |
| CARE0197 | 1 | 13/04/2020 | 13/04/2020 | 0 |
| CARE0198 | 1 | 13/04/2020 | 13/04/2020 | 0 |
| CARE0199 | 4 | 13/04/2020 | 30/04/2020 | 17 |
| CARE0201 | 1 | 13/04/2020 | 13/04/2020 | 0 |
| CARE0202 | 1 | 14/04/2020 | 14/04/2020 | 0 |
| CARE0203 | 1 | 14/04/2020 | 14/04/2020 | 0 |
| CARE0204 | 3 | 12/04/2020 | 17/04/2020 | 5 |
| CARE0205 | 2 | 13/04/2020 | 17/04/2020 | 4 |
| CARE0206 | 1 | 13/04/2020 | 13/04/2020 | 0 |
| CARE0207 | 1 | 14/04/2020 | 14/04/2020 | 0 |
| CARE0208 | 3 | 14/04/2020 | 14/04/2020 | 0 |
| CARE0209 | 3 | 14/04/2020 | 02/05/2020 | 18 |
| CARE0210 | 3 | 14/04/2020 | 26/04/2020 | 12 |
| CARE0211 | 5 | 14/04/2020 | 20/04/2020 | 6 |
| CARE0212 | 1 | 14/04/2020 | 14/04/2020 | 0 |
| CARE0213 | 1 | 14/04/2020 | 14/04/2020 | 0 |
| CARE0214 | 3 | 14/04/2020 | 25/04/2020 | 11 |
| CARE0215 | 5 | 14/04/2020 | 21/04/2020 | 7 |
| CARE0216 | 1 | 14/04/2020 | 14/04/2020 | 0 |
| CARE0217 | 1 | 15/04/2020 | 15/04/2020 | 0 |
| CARE0218 | 1 | 15/04/2020 | 15/04/2020 | 0 |
| CARE0219 | 1 | 15/04/2020 | 15/04/2020 | 0 |
| CARE0220 | 1 | 16/04/2020 | 16/04/2020 | 0 |
| CARE0221 | 2 | 21/04/2020 | 23/04/2020 | 2 |
| CARE0222 | 1 | 15/04/2020 | 15/04/2020 | 0 |
| CARE0223 | 1 | 15/04/2020 | 15/04/2020 | 0 |
| CARE0224 | 2 | 16/04/2020 | 21/04/2020 | 5 |
| CARE0225 | 1 | 16/04/2020 | 16/04/2020 | 0 |
| CARE0226 | 1 | 16/04/2020 | 16/04/2020 | 0 |
| CARE0227 | 1 | 16/04/2020 | 16/04/2020 | 0 |
| CARE0228 | 4 | 15/04/2020 | 20/04/2020 | 5 |
| CARE0229 | 2 | 16/04/2020 | 21/04/2020 | 5 |
| CARE0230 | 1 | 15/04/2020 | 15/04/2020 | 0 |
| CARE0231 | 1 | 16/04/2020 | 16/04/2020 | 0 |
| CARE0232 | 1 | 17/04/2020 | 17/04/2020 | 0 |
| CARE0233 | 1 | 16/04/2020 | 16/04/2020 | 0 |
| CARE0234 | 1 | 17/04/2020 | 17/04/2020 | 0 |
| CARE0235 | 1 | 16/04/2020 | 16/04/2020 | 0 |
| CARE0236 | 2 | 16/04/2020 | 20/04/2020 | 4 |
| CARE0237 | 1 | 17/04/2020 | 17/04/2020 | 0 |
| CARE0239 | 2 | 18/04/2020 | 18/04/2020 | 0 |
| CARE0240 | 2 | 17/04/2020 | 17/04/2020 | 0 |
| CARE0241 | 1 | 17/04/2020 | 17/04/2020 | 0 |
| CARE0242 | 2 | 17/04/2020 | 28/04/2020 | 11 |
| CARE0243 | 2 | 18/04/2020 | 21/04/2020 | 3 |
| CARE0244 | 3 | 19/04/2020 | 07/05/2020 | 18 |
| CARE0245 | 1 | 18/04/2020 | 18/04/2020 | 0 |
| CARE0246 | 1 | 18/04/2020 | 18/04/2020 | 0 |
| CARE0247 | 1 | 19/04/2020 | 19/04/2020 | 0 |
| CARE0249 | 1 | 20/04/2020 | 20/04/2020 | 0 |
| CARE0250 | 2 | 19/04/2020 | 28/04/2020 | 9 |
| CARE0251 | 2 | 19/04/2020 | 01/05/2020 | 12 |
| CARE0253 | 4 | 19/04/2020 | 04/05/2020 | 15 |
| CARE0254 | 2 | 20/04/2020 | 29/04/2020 | 9 |
| CARE0255 | 1 | 20/04/2020 | 20/04/2020 | 0 |
| CARE0257 | 1 | 20/04/2020 | 20/04/2020 | 0 |
| CARE0258 | 4 | 20/04/2020 | 01/05/2020 | 11 |
| CARE0259 | 1 | 20/04/2020 | 20/04/2020 | 0 |
| CARE0260 | 1 | 20/04/2020 | 20/04/2020 | 0 |
| CARE0261 | 6 | 20/04/2020 | 01/05/2020 | 11 |
| CARE0262 | 1 | 21/04/2020 | 21/04/2020 | 0 |
| CARE0263 | 12 | 07/05/2020 | 10/05/2020 | 3 |
| CARE0264 | 9 | 22/04/2020 | 06/05/2020 | 14 |
| CARE0265 | 4 | 21/04/2020 | 21/04/2020 | 0 |
| CARE0266 | 4 | 21/04/2020 | 21/04/2020 | 0 |
| CARE0270 | 2 | 02/05/2020 | 03/05/2020 | 1 |
| CARE0271 | 1 | 22/04/2020 | 22/04/2020 | 0 |
| CARE0272 | 1 | 27/04/2020 | 27/04/2020 | 0 |
| CARE0273 | 2 | 23/04/2020 | 24/04/2020 | 1 |
| CARE0274 | 3 | 24/04/2020 | 09/05/2020 | 15 |
| CARE0276 | 1 | 04/05/2020 | 04/05/2020 | 0 |
| CARE0277 | 13 | 25/04/2020 | 08/05/2020 | 13 |
| CARE0279 | 4 | 29/04/2020 | 30/04/2020 | 1 |
| CARE0281 | 1 | 01/05/2020 | 01/05/2020 | 0 |
| CARE0284 | 2 | 26/04/2020 | 01/05/2020 | 5 |
| CARE0289 | 3 | 02/05/2020 | 05/05/2020 | 3 |
| CARE0293 | 2 | 22/04/2020 | 30/04/2020 | 8 |
| CARE0294 | 1 | 26/04/2020 | 26/04/2020 | 0 |
| CARE0296 | 1 | 25/04/2020 | 25/04/2020 | 0 |
| CARE0297 | 1 | 26/04/2020 | 26/04/2020 | 0 |
| CARE0298 | 1 | 26/04/2020 | 26/04/2020 | 0 |
| CARE0299 | 3 | 26/04/2020 | 26/04/2020 | 0 |
| CARE0300 | 1 | 26/04/2020 | 26/04/2020 | 0 |
| CARE0301 | 2 | 26/04/2020 | 26/04/2020 | 0 |
| CARE0302 | 1 | 26/04/2020 | 26/04/2020 | 0 |
| CARE0304 | 1 | 04/05/2020 | 04/05/2020 | 0 |
| CARE0305 | 1 | 27/04/2020 | 27/04/2020 | 0 |
| CARE0306 | 3 | 14/04/2020 | 27/04/2020 | 13 |
| CARE0307 | 1 | 26/04/2020 | 26/04/2020 | 0 |
| CARE0308 | 5 | 22/04/2020 | 28/04/2020 | 6 |
| CARE0309 | 5 | 01/05/2020 | 08/05/2020 | 7 |
| CARE0310 | 2 | 28/04/2020 | 29/04/2020 | 1 |
| CARE0311 | 1 | 27/04/2020 | 27/04/2020 | 0 |
| CARE0312 | 4 | 05/05/2020 | 05/05/2020 | 0 |
| CARE0314 | 18 | 04/05/2020 | 09/05/2020 | 5 |
| CARE0315 | 1 | 29/04/2020 | 29/04/2020 | 0 |
| CARE0316 | 1 | 30/04/2020 | 30/04/2020 | 0 |
| CARE0317 | 1 | 30/04/2020 | 30/04/2020 | 0 |
| CARE0318 | 1 | 30/04/2020 | 30/04/2020 | 0 |
| CARE0320 | 4 | 01/05/2020 | 07/05/2020 | 6 |
| CARE0322 | 3 | 01/05/2020 | 01/05/2020 | 0 |
| CARE0323 | 2 | 02/05/2020 | 07/05/2020 | 5 |
| CARE0324 | 1 | 01/05/2020 | 01/05/2020 | 0 |
| CARE0325 | 2 | 02/05/2020 | 02/05/2020 | 0 |
| CARE0326 | 2 | 02/05/2020 | 03/05/2020 | 1 |
| CARE0327 | 3 | 01/05/2020 | 01/05/2020 | 0 |
| CARE0328 | 1 | 02/05/2020 | 02/05/2020 | 0 |
| CARE0329 | 2 | 01/05/2020 | 07/05/2020 | 6 |
| CARE0330 | 5 | 07/05/2020 | 07/05/2020 | 0 |
| CARE0331 | 1 | 10/05/2020 | 10/05/2020 | 0 |
| CARE0332 | 1 | 07/05/2020 | 07/05/2020 | 0 |
| CARE0333 | 3 | 06/05/2020 | 06/05/2020 | 0 |
| CARE0335 | 1 | 06/05/2020 | 06/05/2020 | 0 |

### Sampling date ranges for care home residents with genomic data: by cluster defined by the *transcluster* algorithm

Table shows anonymised cluster codes for the 409 clusters from 700 care home residents with genomic data, defined using the *transcluster* algorithm (described in Methods). Each cluster code consists of the anonymised care home code with an increasing numeric suffix. “Number of residents” shows the number of residents with genomic data for each cluster. The date of the first and last samples collected from each cluster and the date difference in days between these samples is shown. If there was only 1 sample from the cluster, and/or all cases were from the same date, then “first sample date” and “last sample date” are the same. Collection date was used for sample dates; if collection date was missing then receive date in the laboratory was used instead. 133 / 409 (33%) clusters had 2 or more cases with genomic data, including 424 care home residents. Of these, the median date difference from first to last sampling dates was 5 days (interquartile range, IQR: 1 – 11; range: 0 – 22). This date range was smaller than for care homes with ≥2 genomes (9 days, IQR: 4 – 15, range: 0 – 50) (*P* = 9.2e-06, Wilcoxon rank sum test), comparison shown in Figure 7, Supplement 6.

| **Care home code** | **Cluster code** | **Number of residents** | **First sample date** | **Last sample date** | **Date difference (days)** |
| --- | --- | --- | --- | --- | --- |
| CARE0001 | CARE0001_1 | 2 | 11/03/2020 | 17/03/2020 | 6 |
| CARE0002 | CARE0002_1 | 2 | 12/03/2020 | 14/03/2020 | 2 |
| CARE0004 | CARE0004_1 | 2 | 13/03/2020 | 13/03/2020 | 0 |
| CARE0005 | CARE0005_1 | 1 | 16/03/2020 | 16/03/2020 | 0 |
| CARE0005 | CARE0005_2 | 1 | 30/04/2020 | 30/04/2020 | 0 |
| CARE0006 | CARE0006_1 | 1 | 14/03/2020 | 14/03/2020 | 0 |
| CARE0006 | CARE0006_2 | 1 | 16/03/2020 | 16/03/2020 | 0 |
| CARE0007 | CARE0007_1 | 1 | 18/03/2020 | 18/03/2020 | 0 |
| CARE0008 | CARE0008_1 | 1 | 16/03/2020 | 16/03/2020 | 0 |
| CARE0008 | CARE0008_2 | 1 | 05/04/2020 | 05/04/2020 | 0 |
| CARE0010 | CARE0010_1 | 1 | 19/03/2020 | 19/03/2020 | 0 |
| CARE0010 | CARE0010_2 | 1 | 20/03/2020 | 20/03/2020 | 0 |
| CARE0011 | CARE0011_1 | 5 | 19/03/2020 | 31/03/2020 | 12 |
| CARE0012 | CARE0012_1 | 1 | 20/03/2020 | 20/03/2020 | 0 |
| CARE0012 | CARE0012_2 | 3 | 31/03/2020 | 05/04/2020 | 5 |
| CARE0014 | CARE0014_1 | 1 | 20/03/2020 | 20/03/2020 | 0 |
| CARE0014 | CARE0014_2 | 3 | 07/05/2020 | 09/05/2020 | 2 |
| CARE0015 | CARE0015_1 | 1 | 20/03/2020 | 20/03/2020 | 0 |
| CARE0015 | CARE0015_2 | 1 | 20/04/2020 | 20/04/2020 | 0 |
| CARE0016 | CARE0016_1 | 1 | 22/03/2020 | 22/03/2020 | 0 |
| CARE0017 | CARE0017_1 | 1 | 21/03/2020 | 21/03/2020 | 0 |
| CARE0017 | CARE0017_2 | 1 | 05/04/2020 | 05/04/2020 | 0 |
| CARE0018 | CARE0018_1 | 1 | 21/03/2020 | 21/03/2020 | 0 |
| CARE0018 | CARE0018_2 | 1 | 06/04/2020 | 06/04/2020 | 0 |
| CARE0019 | CARE0019_1 | 1 | 21/03/2020 | 21/03/2020 | 0 |
| CARE0020 | CARE0020_1 | 2 | 21/03/2020 | 31/03/2020 | 10 |
| CARE0021 | CARE0021_1 | 1 | 21/03/2020 | 21/03/2020 | 0 |
| CARE0021 | CARE0021_2 | 1 | 30/03/2020 | 30/03/2020 | 0 |
| CARE0022 | CARE0022_1 | 1 | 23/03/2020 | 23/03/2020 | 0 |
| CARE0023 | CARE0023_1 | 1 | 22/03/2020 | 22/03/2020 | 0 |
| CARE0023 | CARE0023_2 | 1 | 07/04/2020 | 07/04/2020 | 0 |
| CARE0024 | CARE0024_1 | 1 | 22/03/2020 | 22/03/2020 | 0 |
| CARE0024 | CARE0024_2 | 1 | 05/04/2020 | 05/04/2020 | 0 |
| CARE0024 | CARE0024_3 | 1 | 20/04/2020 | 20/04/2020 | 0 |
| CARE0025 | CARE0025_1 | 1 | 23/03/2020 | 23/03/2020 | 0 |
| CARE0025 | CARE0025_2 | 1 | 03/04/2020 | 03/04/2020 | 0 |
| CARE0026 | CARE0026_2 | 1 | 31/03/2020 | 31/03/2020 | 0 |
| CARE0026 | CARE0026_1 | 2 | 23/03/2020 | 02/04/2020 | 10 |
| CARE0027 | CARE0027_1 | 1 | 02/04/2020 | 02/04/2020 | 0 |
| CARE0028 | CARE0028_1 | 6 | 29/03/2020 | 06/04/2020 | 8 |
| CARE0029 | CARE0029_1 | 2 | 05/04/2020 | 10/04/2020 | 5 |
| CARE0029 | CARE0029_2 | 2 | 06/04/2020 | 13/04/2020 | 7 |
| CARE0030 | CARE0030_1 | 1 | 26/03/2020 | 26/03/2020 | 0 |
| CARE0030 | CARE0030_2 | 1 | 27/04/2020 | 27/04/2020 | 0 |
| CARE0031 | CARE0031_1 | 1 | 09/04/2020 | 09/04/2020 | 0 |
| CARE0032 | CARE0032_1 | 1 | 01/04/2020 | 01/04/2020 | 0 |
| CARE0032 | CARE0032_2 | 6 | 30/04/2020 | 10/05/2020 | 10 |
| CARE0033 | CARE0033_1 | 1 | 31/03/2020 | 31/03/2020 | 0 |
| CARE0034 | CARE0034_1 | 1 | 26/03/2020 | 26/03/2020 | 0 |
| CARE0036 | CARE0036_1 | 1 | 03/04/2020 | 03/04/2020 | 0 |
| CARE0036 | CARE0036_4 | 1 | 19/04/2020 | 19/04/2020 | 0 |
| CARE0036 | CARE0036_2 | 2 | 05/04/2020 | 08/04/2020 | 3 |
| CARE0036 | CARE0036_3 | 2 | 20/04/2020 | 01/05/2020 | 11 |
| CARE0037 | CARE0037_1 | 1 | 30/03/2020 | 30/03/2020 | 0 |
| CARE0038 | CARE0038_1 | 1 | 04/04/2020 | 04/04/2020 | 0 |
| CARE0038 | CARE0038_2 | 1 | 06/04/2020 | 06/04/2020 | 0 |
| CARE0039 | CARE0039_1 | 1 | 06/05/2020 | 06/05/2020 | 0 |
| CARE0040 | CARE0040_1 | 1 | 27/03/2020 | 27/03/2020 | 0 |
| CARE0042 | CARE0042_1 | 1 | 10/04/2020 | 10/04/2020 | 0 |
| CARE0043 | CARE0043_1 | 3 | 09/04/2020 | 11/04/2020 | 2 |
| CARE0046 | CARE0046_1 | 1 | 18/04/2020 | 18/04/2020 | 0 |
| CARE0046 | CARE0046_2 | 1 | 07/05/2020 | 07/05/2020 | 0 |
| CARE0047 | CARE0047_3 | 1 | 07/05/2020 | 07/05/2020 | 0 |
| CARE0047 | CARE0047_1 | 2 | 02/04/2020 | 16/04/2020 | 14 |
| CARE0047 | CARE0047_2 | 2 | 02/04/2020 | 14/04/2020 | 12 |
| CARE0048 | CARE0048_1 | 1 | 27/03/2020 | 27/03/2020 | 0 |
| CARE0049 | CARE0049_1 | 1 | 31/03/2020 | 31/03/2020 | 0 |
| CARE0051 | CARE0051_1 | 2 | 10/04/2020 | 21/04/2020 | 11 |
| CARE0054 | CARE0054_1 | 1 | 05/04/2020 | 05/04/2020 | 0 |
| CARE0054 | CARE0054_2 | 1 | 14/04/2020 | 14/04/2020 | 0 |
| CARE0054 | CARE0054_3 | 2 | 02/05/2020 | 02/05/2020 | 0 |
| CARE0055 | CARE0055_1 | 1 | 31/03/2020 | 31/03/2020 | 0 |
| CARE0056 | CARE0056_1 | 1 | 29/03/2020 | 29/03/2020 | 0 |
| CARE0057 | CARE0057_2 | 1 | 30/03/2020 | 30/03/2020 | 0 |
| CARE0057 | CARE0057_3 | 1 | 12/04/2020 | 12/04/2020 | 0 |
| CARE0057 | CARE0057_1 | 3 | 29/03/2020 | 30/03/2020 | 1 |
| CARE0058 | CARE0058_1 | 1 | 28/03/2020 | 28/03/2020 | 0 |
| CARE0058 | CARE0058_2 | 1 | 27/04/2020 | 27/04/2020 | 0 |
| CARE0059 | CARE0059_1 | 1 | 28/03/2020 | 28/03/2020 | 0 |
| CARE0060 | CARE0060_1 | 1 | 29/03/2020 | 29/03/2020 | 0 |
| CARE0060 | CARE0060_2 | 1 | 21/04/2020 | 21/04/2020 | 0 |
| CARE0061 | CARE0061_1 | 1 | 29/03/2020 | 29/03/2020 | 0 |
| CARE0061 | CARE0061_3 | 1 | 22/04/2020 | 22/04/2020 | 0 |
| CARE0061 | CARE0061_4 | 1 | 06/05/2020 | 06/05/2020 | 0 |
| CARE0061 | CARE0061_2 | 7 | 14/04/2020 | 06/05/2020 | 22 |
| CARE0062 | CARE0062_1 | 1 | 30/03/2020 | 30/03/2020 | 0 |
| CARE0062 | CARE0062_2 | 1 | 29/04/2020 | 29/04/2020 | 0 |
| CARE0063 | CARE0063_2 | 1 | 20/04/2020 | 20/04/2020 | 0 |
| CARE0063 | CARE0063_1 | 11 | 30/03/2020 | 17/04/2020 | 18 |
| CARE0064 | CARE0064_1 | 1 | 29/03/2020 | 29/03/2020 | 0 |
| CARE0065 | CARE0065_1 | 1 | 29/03/2020 | 29/03/2020 | 0 |
| CARE0066 | CARE0066_1 | 6 | 29/03/2020 | 08/04/2020 | 10 |
| CARE0067 | CARE0067_1 | 1 | 31/03/2020 | 31/03/2020 | 0 |
| CARE0068 | CARE0068_1 | 1 | 31/03/2020 | 31/03/2020 | 0 |
| CARE0068 | CARE0068_3 | 1 | 08/05/2020 | 08/05/2020 | 0 |
| CARE0068 | CARE0068_2 | 2 | 08/05/2020 | 09/05/2020 | 1 |
| CARE0069 | CARE0069_1 | 1 | 30/03/2020 | 30/03/2020 | 0 |
| CARE0069 | CARE0069_2 | 2 | 06/04/2020 | 07/04/2020 | 1 |
| CARE0070 | CARE0070_1 | 1 | 29/03/2020 | 29/03/2020 | 0 |
| CARE0071 | CARE0071_1 | 1 | 30/03/2020 | 30/03/2020 | 0 |
| CARE0073 | CARE0073_1 | 1 | 30/03/2020 | 30/03/2020 | 0 |
| CARE0073 | CARE0073_2 | 2 | 17/04/2020 | 20/04/2020 | 3 |
| CARE0074 | CARE0074_1 | 2 | 31/03/2020 | 08/04/2020 | 8 |
| CARE0075 | CARE0075_1 | 1 | 30/03/2020 | 30/03/2020 | 0 |
| CARE0075 | CARE0075_2 | 1 | 06/04/2020 | 06/04/2020 | 0 |
| CARE0076 | CARE0076_2 | 1 | 02/04/2020 | 02/04/2020 | 0 |
| CARE0076 | CARE0076_1 | 3 | 30/03/2020 | 12/04/2020 | 13 |
| CARE0077 | CARE0077_1 | 1 | 31/03/2020 | 31/03/2020 | 0 |
| CARE0078 | CARE0078_1 | 1 | 12/04/2020 | 12/04/2020 | 0 |
| CARE0078 | CARE0078_2 | 1 | 06/05/2020 | 06/05/2020 | 0 |
| CARE0079 | CARE0079_1 | 2 | 12/04/2020 | 13/04/2020 | 1 |
| CARE0080 | CARE0080_1 | 3 | 31/03/2020 | 04/04/2020 | 4 |
| CARE0081 | CARE0081_1 | 2 | 31/03/2020 | 11/04/2020 | 11 |
| CARE0082 | CARE0082_1 | 1 | 31/03/2020 | 31/03/2020 | 0 |
| CARE0082 | CARE0082_2 | 1 | 21/04/2020 | 21/04/2020 | 0 |
| CARE0083 | CARE0083_1 | 1 | 31/03/2020 | 31/03/2020 | 0 |
| CARE0083 | CARE0083_2 | 1 | 06/05/2020 | 06/05/2020 | 0 |
| CARE0084 | CARE0084_1 | 2 | 01/04/2020 | 10/04/2020 | 9 |
| CARE0085 | CARE0085_1 | 1 | 31/03/2020 | 31/03/2020 | 0 |
| CARE0086 | CARE0086_1 | 1 | 31/03/2020 | 31/03/2020 | 0 |
| CARE0087 | CARE0087_1 | 1 | 30/03/2020 | 30/03/2020 | 0 |
| CARE0088 | CARE0088_1 | 1 | 31/03/2020 | 31/03/2020 | 0 |
| CARE0089 | CARE0089_1 | 1 | 02/04/2020 | 02/04/2020 | 0 |
| CARE0090 | CARE0090_1 | 1 | 01/04/2020 | 01/04/2020 | 0 |
| CARE0091 | CARE0091_1 | 5 | 02/04/2020 | 16/04/2020 | 14 |
| CARE0092 | CARE0092_1 | 5 | 01/04/2020 | 22/04/2020 | 21 |
| CARE0093 | CARE0093_1 | 4 | 01/04/2020 | 14/04/2020 | 13 |
| CARE0094 | CARE0094_1 | 1 | 02/04/2020 | 02/04/2020 | 0 |
| CARE0094 | CARE0094_2 | 1 | 16/04/2020 | 16/04/2020 | 0 |
| CARE0095 | CARE0095_1 | 1 | 01/04/2020 | 01/04/2020 | 0 |
| CARE0095 | CARE0095_2 | 1 | 08/04/2020 | 08/04/2020 | 0 |
| CARE0096 | CARE0096_1 | 2 | 02/04/2020 | 13/04/2020 | 11 |
| CARE0097 | CARE0097_1 | 1 | 03/04/2020 | 03/04/2020 | 0 |
| CARE0097 | CARE0097_2 | 6 | 17/04/2020 | 01/05/2020 | 14 |
| CARE0098 | CARE0098_1 | 3 | 02/04/2020 | 06/04/2020 | 4 |
| CARE0099 | CARE0099_1 | 2 | 03/04/2020 | 16/04/2020 | 13 |
| CARE0100 | CARE0100_1 | 1 | 02/04/2020 | 02/04/2020 | 0 |
| CARE0100 | CARE0100_2 | 1 | 01/04/2020 | 01/04/2020 | 0 |
| CARE0101 | CARE0101_1 | 2 | 02/04/2020 | 06/04/2020 | 4 |
| CARE0102 | CARE0102_1 | 2 | 02/04/2020 | 16/04/2020 | 14 |
| CARE0102 | CARE0102_2 | 3 | 02/05/2020 | 04/05/2020 | 2 |
| CARE0103 | CARE0103_1 | 1 | 08/04/2020 | 08/04/2020 | 0 |
| CARE0103 | CARE0103_2 | 1 | 21/04/2020 | 21/04/2020 | 0 |
| CARE0104 | CARE0104_1 | 1 | 02/04/2020 | 02/04/2020 | 0 |
| CARE0104 | CARE0104_2 | 1 | 10/04/2020 | 10/04/2020 | 0 |
| CARE0104 | CARE0104_3 | 1 | 10/04/2020 | 10/04/2020 | 0 |
| CARE0105 | CARE0105_1 | 2 | 03/04/2020 | 03/04/2020 | 0 |
| CARE0106 | CARE0106_1 | 3 | 02/04/2020 | 12/04/2020 | 10 |
| CARE0107 | CARE0107_1 | 1 | 03/04/2020 | 03/04/2020 | 0 |
| CARE0108 | CARE0108_1 | 1 | 03/04/2020 | 03/04/2020 | 0 |
| CARE0108 | CARE0108_2 | 1 | 29/04/2020 | 29/04/2020 | 0 |
| CARE0109 | CARE0109_1 | 1 | 04/04/2020 | 04/04/2020 | 0 |
| CARE0109 | CARE0109_2 | 1 | 07/04/2020 | 07/04/2020 | 0 |
| CARE0111 | CARE0111_1 | 3 | 03/04/2020 | 07/04/2020 | 4 |
| CARE0112 | CARE0112_1 | 1 | 03/04/2020 | 03/04/2020 | 0 |
| CARE0113 | CARE0113_1 | 1 | 03/04/2020 | 03/04/2020 | 0 |
| CARE0113 | CARE0113_2 | 1 | 12/04/2020 | 12/04/2020 | 0 |
| CARE0114 | CARE0114_1 | 3 | 07/04/2020 | 15/04/2020 | 8 |
| CARE0116 | CARE0116_1 | 1 | 04/04/2020 | 04/04/2020 | 0 |
| CARE0116 | CARE0116_2 | 1 | 10/04/2020 | 10/04/2020 | 0 |
| CARE0117 | CARE0117_1 | 1 | 04/04/2020 | 04/04/2020 | 0 |
| CARE0117 | CARE0117_2 | 1 | 13/04/2020 | 13/04/2020 | 0 |
| CARE0118 | CARE0118_1 | 3 | 06/04/2020 | 18/04/2020 | 12 |
| CARE0119 | CARE0119_2 | 1 | 06/04/2020 | 06/04/2020 | 0 |
| CARE0119 | CARE0119_1 | 2 | 03/04/2020 | 06/04/2020 | 3 |
| CARE0120 | CARE0120_2 | 1 | 13/04/2020 | 13/04/2020 | 0 |
| CARE0120 | CARE0120_3 | 1 | 01/05/2020 | 01/05/2020 | 0 |
| CARE0120 | CARE0120_1 | 2 | 03/04/2020 | 16/04/2020 | 13 |
| CARE0121 | CARE0121_1 | 1 | 03/04/2020 | 03/04/2020 | 0 |
| CARE0122 | CARE0122_1 | 1 | 04/04/2020 | 04/04/2020 | 0 |
| CARE0122 | CARE0122_2 | 5 | 06/04/2020 | 18/04/2020 | 12 |
| CARE0123 | CARE0123_1 | 1 | 05/04/2020 | 05/04/2020 | 0 |
| CARE0124 | CARE0124_1 | 2 | 05/04/2020 | 11/04/2020 | 6 |
| CARE0125 | CARE0125_2 | 1 | 07/04/2020 | 07/04/2020 | 0 |
| CARE0125 | CARE0125_1 | 5 | 05/04/2020 | 17/04/2020 | 12 |
| CARE0126 | CARE0126_1 | 1 | 05/04/2020 | 05/04/2020 | 0 |
| CARE0126 | CARE0126_2 | 2 | 10/04/2020 | 14/04/2020 | 4 |
| CARE0127 | CARE0127_1 | 1 | 04/04/2020 | 04/04/2020 | 0 |
| CARE0127 | CARE0127_2 | 1 | 18/04/2020 | 18/04/2020 | 0 |
| CARE0128 | CARE0128_1 | 4 | 05/04/2020 | 11/04/2020 | 6 |
| CARE0129 | CARE0129_1 | 1 | 05/04/2020 | 05/04/2020 | 0 |
| CARE0130 | CARE0130_1 | 1 | 05/04/2020 | 05/04/2020 | 0 |
| CARE0131 | CARE0131_1 | 1 | 04/04/2020 | 04/04/2020 | 0 |
| CARE0132 | CARE0132_1 | 1 | 05/04/2020 | 05/04/2020 | 0 |
| CARE0133 | CARE0133_1 | 1 | 04/04/2020 | 04/04/2020 | 0 |
| CARE0134 | CARE0134_1 | 2 | 06/04/2020 | 18/04/2020 | 12 |
| CARE0134 | CARE0134_2 | 2 | 17/04/2020 | 19/04/2020 | 2 |
| CARE0135 | CARE0135_1 | 1 | 05/04/2020 | 05/04/2020 | 0 |
| CARE0135 | CARE0135_2 | 1 | 08/04/2020 | 08/04/2020 | 0 |
| CARE0136 | CARE0136_1 | 1 | 05/04/2020 | 05/04/2020 | 0 |
| CARE0137 | CARE0137_1 | 1 | 05/04/2020 | 05/04/2020 | 0 |
| CARE0138 | CARE0138_1 | 1 | 05/04/2020 | 05/04/2020 | 0 |
| CARE0139 | CARE0139_2 | 1 | 13/04/2020 | 13/04/2020 | 0 |
| CARE0139 | CARE0139_3 | 1 | 15/04/2020 | 15/04/2020 | 0 |
| CARE0139 | CARE0139_1 | 2 | 05/04/2020 | 06/04/2020 | 1 |
| CARE0140 | CARE0140_1 | 1 | 06/04/2020 | 06/04/2020 | 0 |
| CARE0141 | CARE0141_1 | 1 | 07/04/2020 | 07/04/2020 | 0 |
| CARE0142 | CARE0142_1 | 3 | 04/04/2020 | 19/04/2020 | 15 |
| CARE0143 | CARE0143_1 | 1 | 05/04/2020 | 05/04/2020 | 0 |
| CARE0144 | CARE0144_1 | 2 | 05/04/2020 | 07/04/2020 | 2 |
| CARE0145 | CARE0145_1 | 1 | 07/04/2020 | 07/04/2020 | 0 |
| CARE0145 | CARE0145_2 | 1 | 30/04/2020 | 30/04/2020 | 0 |
| CARE0146 | CARE0146_1 | 1 | 09/04/2020 | 09/04/2020 | 0 |
| CARE0146 | CARE0146_2 | 1 | 07/04/2020 | 07/04/2020 | 0 |
| CARE0147 | CARE0147_1 | 1 | 07/04/2020 | 07/04/2020 | 0 |
| CARE0148 | CARE0148_1 | 2 | 07/04/2020 | 07/04/2020 | 0 |
| CARE0149 | CARE0149_2 | 1 | 19/04/2020 | 19/04/2020 | 0 |
| CARE0149 | CARE0149_3 | 1 | 29/04/2020 | 29/04/2020 | 0 |
| CARE0149 | CARE0149_4 | 1 | 07/05/2020 | 07/05/2020 | 0 |
| CARE0149 | CARE0149_1 | 2 | 07/04/2020 | 07/04/2020 | 0 |
| CARE0150 | CARE0150_1 | 2 | 07/04/2020 | 09/04/2020 | 2 |
| CARE0151 | CARE0151_1 | 1 | 07/04/2020 | 07/04/2020 | 0 |
| CARE0151 | CARE0151_3 | 1 | 27/04/2020 | 27/04/2020 | 0 |
| CARE0151 | CARE0151_4 | 1 | 27/04/2020 | 27/04/2020 | 0 |
| CARE0151 | CARE0151_2 | 4 | 27/04/2020 | 27/04/2020 | 0 |
| CARE0153 | CARE0153_1 | 1 | 08/04/2020 | 08/04/2020 | 0 |
| CARE0154 | CARE0154_1 | 1 | 08/04/2020 | 08/04/2020 | 0 |
| CARE0155 | CARE0155_1 | 2 | 08/04/2020 | 21/04/2020 | 13 |
| CARE0156 | CARE0156_2 | 1 | 24/04/2020 | 24/04/2020 | 0 |
| CARE0156 | CARE0156_1 | 2 | 07/04/2020 | 08/04/2020 | 1 |
| CARE0157 | CARE0157_1 | 1 | 08/04/2020 | 08/04/2020 | 0 |
| CARE0157 | CARE0157_3 | 1 | 26/04/2020 | 26/04/2020 | 0 |
| CARE0157 | CARE0157_2 | 3 | 10/04/2020 | 12/04/2020 | 2 |
| CARE0158 | CARE0158_1 | 1 | 08/04/2020 | 08/04/2020 | 0 |
| CARE0159 | CARE0159_2 | 1 | 06/05/2020 | 06/05/2020 | 0 |
| CARE0159 | CARE0159_1 | 5 | 09/04/2020 | 21/04/2020 | 12 |
| CARE0160 | CARE0160_1 | 1 | 09/04/2020 | 09/04/2020 | 0 |
| CARE0161 | CARE0161_2 | 1 | 09/04/2020 | 09/04/2020 | 0 |
| CARE0161 | CARE0161_1 | 2 | 08/04/2020 | 14/04/2020 | 6 |
| CARE0162 | CARE0162_1 | 4 | 09/04/2020 | 01/05/2020 | 22 |
| CARE0165 | CARE0165_1 | 1 | 07/05/2020 | 07/05/2020 | 0 |
| CARE0166 | CARE0166_1 | 4 | 11/04/2020 | 24/04/2020 | 13 |
| CARE0167 | CARE0167_1 | 1 | 13/04/2020 | 13/04/2020 | 0 |
| CARE0169 | CARE0169_1 | 4 | 18/04/2020 | 30/04/2020 | 12 |
| CARE0170 | CARE0170_1 | 1 | 09/04/2020 | 09/04/2020 | 0 |
| CARE0171 | CARE0171_1 | 2 | 09/04/2020 | 16/04/2020 | 7 |
| CARE0172 | CARE0172_1 | 1 | 09/04/2020 | 09/04/2020 | 0 |
| CARE0172 | CARE0172_2 | 1 | 13/04/2020 | 13/04/2020 | 0 |
| CARE0173 | CARE0173_1 | 1 | 10/04/2020 | 10/04/2020 | 0 |
| CARE0173 | CARE0173_2 | 3 | 28/04/2020 | 01/05/2020 | 3 |
| CARE0173 | CARE0173_3 | 3 | 01/05/2020 | 01/05/2020 | 0 |
| CARE0174 | CARE0174_1 | 4 | 09/04/2020 | 17/04/2020 | 8 |
| CARE0175 | CARE0175_1 | 1 | 09/04/2020 | 09/04/2020 | 0 |
| CARE0175 | CARE0175_2 | 1 | 24/04/2020 | 24/04/2020 | 0 |
| CARE0176 | CARE0176_1 | 2 | 10/04/2020 | 18/04/2020 | 8 |
| CARE0177 | CARE0177_1 | 1 | 10/04/2020 | 10/04/2020 | 0 |
| CARE0177 | CARE0177_2 | 2 | 28/04/2020 | 01/05/2020 | 3 |
| CARE0178 | CARE0178_1 | 1 | 11/04/2020 | 11/04/2020 | 0 |
| CARE0179 | CARE0179_1 | 1 | 10/04/2020 | 10/04/2020 | 0 |
| CARE0180 | CARE0180_1 | 1 | 10/04/2020 | 10/04/2020 | 0 |
| CARE0181 | CARE0181_1 | 1 | 10/04/2020 | 10/04/2020 | 0 |
| CARE0181 | CARE0181_2 | 1 | 14/04/2020 | 14/04/2020 | 0 |
| CARE0182 | CARE0182_2 | 1 | 21/04/2020 | 21/04/2020 | 0 |
| CARE0182 | CARE0182_1 | 4 | 11/04/2020 | 21/04/2020 | 10 |
| CARE0183 | CARE0183_1 | 1 | 11/04/2020 | 11/04/2020 | 0 |
| CARE0184 | CARE0184_1 | 3 | 12/04/2020 | 17/04/2020 | 5 |
| CARE0185 | CARE0185_1 | 1 | 11/04/2020 | 11/04/2020 | 0 |
| CARE0186 | CARE0186_1 | 1 | 11/04/2020 | 11/04/2020 | 0 |
| CARE0187 | CARE0187_1 | 1 | 12/04/2020 | 12/04/2020 | 0 |
| CARE0188 | CARE0188_1 | 1 | 11/04/2020 | 11/04/2020 | 0 |
| CARE0189 | CARE0189_1 | 4 | 11/04/2020 | 20/04/2020 | 9 |
| CARE0190 | CARE0190_1 | 2 | 12/04/2020 | 21/04/2020 | 9 |
| CARE0191 | CARE0191_1 | 2 | 11/04/2020 | 14/04/2020 | 3 |
| CARE0192 | CARE0192_1 | 1 | 11/04/2020 | 11/04/2020 | 0 |
| CARE0193 | CARE0193_1 | 1 | 11/04/2020 | 11/04/2020 | 0 |
| CARE0194 | CARE0194_2 | 1 | 28/04/2020 | 28/04/2020 | 0 |
| CARE0194 | CARE0194_1 | 2 | 12/04/2020 | 13/04/2020 | 1 |
| CARE0195 | CARE0195_1 | 1 | 13/04/2020 | 13/04/2020 | 0 |
| CARE0196 | CARE0196_1 | 1 | 13/04/2020 | 13/04/2020 | 0 |
| CARE0196 | CARE0196_2 | 1 | 18/04/2020 | 18/04/2020 | 0 |
| CARE0196 | CARE0196_4 | 1 | 28/04/2020 | 28/04/2020 | 0 |
| CARE0196 | CARE0196_3 | 2 | 28/04/2020 | 28/04/2020 | 0 |
| CARE0197 | CARE0197_1 | 1 | 13/04/2020 | 13/04/2020 | 0 |
| CARE0198 | CARE0198_1 | 1 | 13/04/2020 | 13/04/2020 | 0 |
| CARE0199 | CARE0199_1 | 2 | 13/04/2020 | 21/04/2020 | 8 |
| CARE0199 | CARE0199_2 | 2 | 26/04/2020 | 30/04/2020 | 4 |
| CARE0201 | CARE0201_1 | 1 | 13/04/2020 | 13/04/2020 | 0 |
| CARE0202 | CARE0202_1 | 1 | 14/04/2020 | 14/04/2020 | 0 |
| CARE0203 | CARE0203_1 | 1 | 14/04/2020 | 14/04/2020 | 0 |
| CARE0204 | CARE0204_1 | 1 | 12/04/2020 | 12/04/2020 | 0 |
| CARE0204 | CARE0204_2 | 2 | 13/04/2020 | 17/04/2020 | 4 |
| CARE0205 | CARE0205_1 | 2 | 13/04/2020 | 17/04/2020 | 4 |
| CARE0206 | CARE0206_1 | 1 | 13/04/2020 | 13/04/2020 | 0 |
| CARE0207 | CARE0207_1 | 1 | 14/04/2020 | 14/04/2020 | 0 |
| CARE0208 | CARE0208_2 | 1 | 14/04/2020 | 14/04/2020 | 0 |
| CARE0208 | CARE0208_1 | 2 | 14/04/2020 | 14/04/2020 | 0 |
| CARE0209 | CARE0209_2 | 1 | 02/05/2020 | 02/05/2020 | 0 |
| CARE0209 | CARE0209_1 | 2 | 14/04/2020 | 15/04/2020 | 1 |
| CARE0210 | CARE0210_1 | 3 | 14/04/2020 | 26/04/2020 | 12 |
| CARE0211 | CARE0211_2 | 1 | 15/04/2020 | 15/04/2020 | 0 |
| CARE0211 | CARE0211_3 | 1 | 15/04/2020 | 15/04/2020 | 0 |
| CARE0211 | CARE0211_1 | 3 | 14/04/2020 | 20/04/2020 | 6 |
| CARE0212 | CARE0212_1 | 1 | 14/04/2020 | 14/04/2020 | 0 |
| CARE0213 | CARE0213_1 | 1 | 14/04/2020 | 14/04/2020 | 0 |
| CARE0214 | CARE0214_1 | 3 | 14/04/2020 | 25/04/2020 | 11 |
| CARE0215 | CARE0215_1 | 5 | 14/04/2020 | 21/04/2020 | 7 |
| CARE0216 | CARE0216_1 | 1 | 14/04/2020 | 14/04/2020 | 0 |
| CARE0217 | CARE0217_1 | 1 | 15/04/2020 | 15/04/2020 | 0 |
| CARE0218 | CARE0218_1 | 1 | 15/04/2020 | 15/04/2020 | 0 |
| CARE0219 | CARE0219_1 | 1 | 15/04/2020 | 15/04/2020 | 0 |
| CARE0220 | CARE0220_1 | 1 | 16/04/2020 | 16/04/2020 | 0 |
| CARE0221 | CARE0221_1 | 1 | 21/04/2020 | 21/04/2020 | 0 |
| CARE0221 | CARE0221_2 | 1 | 23/04/2020 | 23/04/2020 | 0 |
| CARE0222 | CARE0222_1 | 1 | 15/04/2020 | 15/04/2020 | 0 |
| CARE0223 | CARE0223_1 | 1 | 15/04/2020 | 15/04/2020 | 0 |
| CARE0224 | CARE0224_1 | 2 | 16/04/2020 | 21/04/2020 | 5 |
| CARE0225 | CARE0225_1 | 1 | 16/04/2020 | 16/04/2020 | 0 |
| CARE0226 | CARE0226_1 | 1 | 16/04/2020 | 16/04/2020 | 0 |
| CARE0227 | CARE0227_1 | 1 | 16/04/2020 | 16/04/2020 | 0 |
| CARE0228 | CARE0228_1 | 4 | 15/04/2020 | 20/04/2020 | 5 |
| CARE0229 | CARE0229_1 | 1 | 16/04/2020 | 16/04/2020 | 0 |
| CARE0229 | CARE0229_2 | 1 | 21/04/2020 | 21/04/2020 | 0 |
| CARE0230 | CARE0230_1 | 1 | 15/04/2020 | 15/04/2020 | 0 |
| CARE0231 | CARE0231_1 | 1 | 16/04/2020 | 16/04/2020 | 0 |
| CARE0232 | CARE0232_1 | 1 | 17/04/2020 | 17/04/2020 | 0 |
| CARE0233 | CARE0233_1 | 1 | 16/04/2020 | 16/04/2020 | 0 |
| CARE0234 | CARE0234_1 | 1 | 17/04/2020 | 17/04/2020 | 0 |
| CARE0235 | CARE0235_1 | 1 | 16/04/2020 | 16/04/2020 | 0 |
| CARE0236 | CARE0236_1 | 1 | 16/04/2020 | 16/04/2020 | 0 |
| CARE0236 | CARE0236_2 | 1 | 20/04/2020 | 20/04/2020 | 0 |
| CARE0237 | CARE0237_1 | 1 | 17/04/2020 | 17/04/2020 | 0 |
| CARE0239 | CARE0239_1 | 1 | 18/04/2020 | 18/04/2020 | 0 |
| CARE0239 | CARE0239_2 | 1 | 18/04/2020 | 18/04/2020 | 0 |
| CARE0240 | CARE0240_1 | 2 | 17/04/2020 | 17/04/2020 | 0 |
| CARE0241 | CARE0241_1 | 1 | 17/04/2020 | 17/04/2020 | 0 |
| CARE0242 | CARE0242_1 | 2 | 17/04/2020 | 28/04/2020 | 11 |
| CARE0243 | CARE0243_1 | 2 | 18/04/2020 | 21/04/2020 | 3 |
| CARE0244 | CARE0244_1 | 1 | 19/04/2020 | 19/04/2020 | 0 |
| CARE0244 | CARE0244_2 | 1 | 07/05/2020 | 07/05/2020 | 0 |
| CARE0244 | CARE0244_3 | 1 | 07/05/2020 | 07/05/2020 | 0 |
| CARE0245 | CARE0245_1 | 1 | 18/04/2020 | 18/04/2020 | 0 |
| CARE0246 | CARE0246_1 | 1 | 18/04/2020 | 18/04/2020 | 0 |
| CARE0247 | CARE0247_1 | 1 | 19/04/2020 | 19/04/2020 | 0 |
| CARE0249 | CARE0249_1 | 1 | 20/04/2020 | 20/04/2020 | 0 |
| CARE0250 | CARE0250_1 | 2 | 19/04/2020 | 28/04/2020 | 9 |
| CARE0251 | CARE0251_1 | 1 | 19/04/2020 | 19/04/2020 | 0 |
| CARE0251 | CARE0251_2 | 1 | 01/05/2020 | 01/05/2020 | 0 |
| CARE0253 | CARE0253_1 | 4 | 19/04/2020 | 04/05/2020 | 15 |
| CARE0254 | CARE0254_1 | 2 | 20/04/2020 | 29/04/2020 | 9 |
| CARE0255 | CARE0255_1 | 1 | 20/04/2020 | 20/04/2020 | 0 |
| CARE0257 | CARE0257_1 | 1 | 20/04/2020 | 20/04/2020 | 0 |
| CARE0258 | CARE0258_1 | 4 | 20/04/2020 | 01/05/2020 | 11 |
| CARE0259 | CARE0259_1 | 1 | 20/04/2020 | 20/04/2020 | 0 |
| CARE0260 | CARE0260_1 | 1 | 20/04/2020 | 20/04/2020 | 0 |
| CARE0261 | CARE0261_1 | 6 | 20/04/2020 | 01/05/2020 | 11 |
| CARE0262 | CARE0262_1 | 1 | 21/04/2020 | 21/04/2020 | 0 |
| CARE0263 | CARE0263_3 | 1 | 07/05/2020 | 07/05/2020 | 0 |
| CARE0263 | CARE0263_2 | 2 | 07/05/2020 | 07/05/2020 | 0 |
| CARE0263 | CARE0263_1 | 9 | 07/05/2020 | 10/05/2020 | 3 |
| CARE0264 | CARE0264_1 | 9 | 22/04/2020 | 06/05/2020 | 14 |
| CARE0265 | CARE0265_2 | 1 | 21/04/2020 | 21/04/2020 | 0 |
| CARE0265 | CARE0265_1 | 3 | 21/04/2020 | 21/04/2020 | 0 |
| CARE0266 | CARE0266_2 | 1 | 21/04/2020 | 21/04/2020 | 0 |
| CARE0266 | CARE0266_1 | 3 | 21/04/2020 | 21/04/2020 | 0 |
| CARE0270 | CARE0270_1 | 2 | 02/05/2020 | 03/05/2020 | 1 |
| CARE0271 | CARE0271_1 | 1 | 22/04/2020 | 22/04/2020 | 0 |
| CARE0272 | CARE0272_1 | 1 | 27/04/2020 | 27/04/2020 | 0 |
| CARE0273 | CARE0273_1 | 2 | 23/04/2020 | 24/04/2020 | 1 |
| CARE0274 | CARE0274_2 | 1 | 09/05/2020 | 09/05/2020 | 0 |
| CARE0274 | CARE0274_1 | 2 | 24/04/2020 | 06/05/2020 | 12 |
| CARE0276 | CARE0276_1 | 1 | 04/05/2020 | 04/05/2020 | 0 |
| CARE0277 | CARE0277_2 | 1 | 07/05/2020 | 07/05/2020 | 0 |
| CARE0277 | CARE0277_1 | 12 | 25/04/2020 | 08/05/2020 | 13 |
| CARE0279 | CARE0279_1 | 4 | 29/04/2020 | 30/04/2020 | 1 |
| CARE0281 | CARE0281_1 | 1 | 01/05/2020 | 01/05/2020 | 0 |
| CARE0284 | CARE0284_1 | 1 | 26/04/2020 | 26/04/2020 | 0 |
| CARE0284 | CARE0284_2 | 1 | 01/05/2020 | 01/05/2020 | 0 |
| CARE0289 | CARE0289_1 | 3 | 02/05/2020 | 05/05/2020 | 3 |
| CARE0293 | CARE0293_1 | 2 | 22/04/2020 | 30/04/2020 | 8 |
| CARE0294 | CARE0294_1 | 1 | 26/04/2020 | 26/04/2020 | 0 |
| CARE0296 | CARE0296_1 | 1 | 25/04/2020 | 25/04/2020 | 0 |
| CARE0297 | CARE0297_1 | 1 | 26/04/2020 | 26/04/2020 | 0 |
| CARE0298 | CARE0298_1 | 1 | 26/04/2020 | 26/04/2020 | 0 |
| CARE0299 | CARE0299_1 | 3 | 26/04/2020 | 26/04/2020 | 0 |
| CARE0300 | CARE0300_1 | 1 | 26/04/2020 | 26/04/2020 | 0 |
| CARE0301 | CARE0301_1 | 2 | 26/04/2020 | 26/04/2020 | 0 |
| CARE0302 | CARE0302_1 | 1 | 26/04/2020 | 26/04/2020 | 0 |
| CARE0304 | CARE0304_1 | 1 | 04/05/2020 | 04/05/2020 | 0 |
| CARE0305 | CARE0305_1 | 1 | 27/04/2020 | 27/04/2020 | 0 |
| CARE0306 | CARE0306_1 | 3 | 14/04/2020 | 27/04/2020 | 13 |
| CARE0307 | CARE0307_1 | 1 | 26/04/2020 | 26/04/2020 | 0 |
| CARE0308 | CARE0308_1 | 5 | 22/04/2020 | 28/04/2020 | 6 |
| CARE0309 | CARE0309_1 | 5 | 01/05/2020 | 08/05/2020 | 7 |
| CARE0310 | CARE0310_1 | 2 | 28/04/2020 | 29/04/2020 | 1 |
| CARE0311 | CARE0311_1 | 1 | 27/04/2020 | 27/04/2020 | 0 |
| CARE0312 | CARE0312_2 | 1 | 05/05/2020 | 05/05/2020 | 0 |
| CARE0312 | CARE0312_1 | 3 | 05/05/2020 | 05/05/2020 | 0 |
| CARE0314 | CARE0314_1 | 18 | 04/05/2020 | 09/05/2020 | 5 |
| CARE0315 | CARE0315_1 | 1 | 29/04/2020 | 29/04/2020 | 0 |
| CARE0316 | CARE0316_1 | 1 | 30/04/2020 | 30/04/2020 | 0 |
| CARE0317 | CARE0317_1 | 1 | 30/04/2020 | 30/04/2020 | 0 |
| CARE0318 | CARE0318_1 | 1 | 30/04/2020 | 30/04/2020 | 0 |
| CARE0320 | CARE0320_1 | 2 | 01/05/2020 | 01/05/2020 | 0 |
| CARE0320 | CARE0320_2 | 2 | 01/05/2020 | 07/05/2020 | 6 |
| CARE0322 | CARE0322_2 | 1 | 01/05/2020 | 01/05/2020 | 0 |
| CARE0322 | CARE0322_1 | 2 | 01/05/2020 | 01/05/2020 | 0 |
| CARE0323 | CARE0323_1 | 2 | 02/05/2020 | 07/05/2020 | 5 |
| CARE0324 | CARE0324_1 | 1 | 01/05/2020 | 01/05/2020 | 0 |
| CARE0325 | CARE0325_1 | 2 | 02/05/2020 | 02/05/2020 | 0 |
| CARE0326 | CARE0326_1 | 2 | 02/05/2020 | 03/05/2020 | 1 |
| CARE0327 | CARE0327_1 | 3 | 01/05/2020 | 01/05/2020 | 0 |
| CARE0328 | CARE0328_1 | 1 | 02/05/2020 | 02/05/2020 | 0 |
| CARE0329 | CARE0329_1 | 1 | 01/05/2020 | 01/05/2020 | 0 |
| CARE0329 | CARE0329_2 | 1 | 07/05/2020 | 07/05/2020 | 0 |
| CARE0330 | CARE0330_1 | 5 | 07/05/2020 | 07/05/2020 | 0 |
| CARE0331 | CARE0331_1 | 1 | 10/05/2020 | 10/05/2020 | 0 |
| CARE0332 | CARE0332_1 | 1 | 07/05/2020 | 07/05/2020 | 0 |
| CARE0333 | CARE0333_1 | 3 | 06/05/2020 | 06/05/2020 | 0 |
| CARE0335 | CARE0335_1 | 1 | 06/05/2020 | 06/05/2020 | 0 |

### Cambridge COG-UK IDs for all samples analysed in this study

The main analysis set comprised 700 genomes from care home residents. Additionally, a randomised selection of 700 genomes from non-care home residents was used for comparing lineage composition, and genomes from 76 healthcare workers tested at CUH were included for the analysis of care home resident-HCW transmission. Consensus fasta sequences for the 1,476 genomes are publicly accessible through the COG-UK website data section (<https://www.cogconsortium.uk/data/>). COG-UK also regularly deposits data into public databases such as GISAID (<https://www.gisaid.org/>). COG-UK sequence codes, GISAID accession IDs and virus names for the 1,476 analysed genomes are included below. “Sample date” refers to date of sampling collection; if this is missing then sample receive date in the diagnostic laboratory is used instead.

| **COG-UK ID** | **Virus name** | **Accession ID** | **Sample date** |
| --- | --- | --- | --- |
| CAMB-8473C | hCoV-19/England/CAMB-8473C/2020 | EPI_ISL_440499 | 26/02/2020 |
| CAMB-84778 | hCoV-19/England/CAMB-84778/2020 | EPI_ISL_440514 | 28/02/2020 |
| CAMB-84875 | hCoV-19/England/CAMB-84875/2020 | EPI_ISL_440554 | 04/03/2020 |
| CAMB-84945 | hCoV-19/England/CAMB-84945/2020 | EPI_ISL_440535 | 07/03/2020 |
| CAMB-84954 | hCoV-19/England/CAMB-84954/2020 | EPI_ISL_440474 | 07/03/2020 |
| CAMB-849CD | hCoV-19/England/CAMB-849CD/2020 | EPI_ISL_440479 | 09/03/2020 |
| CAMB-846E4 | hCoV-19/England/CAMB-846E4/2020 | EPI_ISL_440534 | 10/03/2020 |
| CAMB-84A06 | hCoV-19/England/CAMB-84A06/2020 | EPI_ISL_440543 | 10/03/2020 |
| CAMB-84AAC | hCoV-19/England/CAMB-84AAC/2020 | EPI_ISL_440545 | 10/03/2020 |
| CAMB-84AD9 | hCoV-19/England/CAMB-84AD9/2020 | EPI_ISL_440490 | 10/03/2020 |
| CAMB-84AF7 | hCoV-19/England/CAMB-84AF7/2020 | EPI_ISL_440523 | 10/03/2020 |
| CAMB-84B6D | hCoV-19/England/CAMB-84B6D/2020 | EPI_ISL_440525 | 10/03/2020 |
| CAMB-84B12 | hCoV-19/England/CAMB-84B12/2020 | EPI_ISL_440477 | 11/03/2020 |
| CAMB-84BB8 | hCoV-19/England/CAMB-84BB8/2020 | EPI_ISL_440485 | 11/03/2020 |
| CAMB-84C3D | hCoV-19/England/CAMB-84C3D/2020 | EPI_ISL_440547 | 11/03/2020 |
| CAMB-84F70 | hCoV-19/England/CAMB-84F70/2020 | EPI_ISL_440451 | 11/03/2020 |
| CAMB-7524C | hCoV-19/England/CAMB-7524C/2020 | EPI_ISL_440375 | 12/03/2020 |
| CAMB-753B2 | hCoV-19/England/CAMB-753B2/2020 | EPI_ISL_440378 | 12/03/2020 |
| CAMB-75622 | hCoV-19/England/CAMB-75622/2020 | EPI_ISL_425415 | 12/03/2020 |
| CAMB-84B8B | hCoV-19/England/CAMB-84B8B/2020 | EPI_ISL_440516 | 12/03/2020 |
| CAMB-84C1F | hCoV-19/England/CAMB-84C1F/2020 | EPI_ISL_440539 | 12/03/2020 |
| CAMB-84D76 | hCoV-19/England/CAMB-84D76/2020 | EPI_ISL_440470 | 12/03/2020 |
| CAMB-84E73 | hCoV-19/England/CAMB-84E73/2020 | EPI_ISL_440465 | 12/03/2020 |
| CAMB-754FB | hCoV-19/England/CAMB-754FB/2020 | EPI_ISL_425399 | 13/03/2020 |
| CAMB-75516 | hCoV-19/England/CAMB-75516/2020 | EPI_ISL_425401 | 13/03/2020 |
| CAMB-75525 | hCoV-19/England/CAMB-75525/2020 | EPI_ISL_425402 | 13/03/2020 |
| CAMB-84E37 | hCoV-19/England/CAMB-84E37/2020 | EPI_ISL_440459 | 13/03/2020 |
| CAMB-84F9E | hCoV-19/England/CAMB-84F9E/2020 | EPI_ISL_440464 | 13/03/2020 |
| CAMB-84FBC | hCoV-19/England/CAMB-84FBC/2020 | EPI_ISL_440443 | 13/03/2020 |
| CAMB-74FBF | hCoV-19/England/CAMB-74FBF/2020 | EPI_ISL_440380 | 14/03/2020 |
| CAMB-751E5 | hCoV-19/England/CAMB-751E5/2020 | EPI_ISL_440396 | 14/03/2020 |
| CAMB-7525B | hCoV-19/England/CAMB-7525B/2020 | EPI_ISL_440403 | 14/03/2020 |
| CAMB-75437 | hCoV-19/England/CAMB-75437/2020 | EPI_ISL_425388 | 14/03/2020 |
| CAMB-84E19 | hCoV-19/England/CAMB-84E19/2020 | EPI_ISL_440439 | 14/03/2020 |
| CAMB-84EBF | hCoV-19/England/CAMB-84EBF/2020 | EPI_ISL_440462 | 14/03/2020 |
| CAMB-750CA | hCoV-19/England/CAMB-750CA/2020 | EPI_ISL_440424 | 15/03/2020 |
| CAMB-7537E | hCoV-19/England/CAMB-7537E/2020 | EPI_ISL_425386 | 15/03/2020 |
| CAMB-75491 | hCoV-19/England/CAMB-75491/2020 | EPI_ISL_425393 | 15/03/2020 |
| CAMB-755BC | hCoV-19/England/CAMB-755BC/2020 | EPI_ISL_425408 | 15/03/2020 |
| CAMB-74C21 | hCoV-19/England/CAMB-74C21/2020 | EPI_ISL_425358 | 16/03/2020 |
| CAMB-74CA9 | hCoV-19/England/CAMB-74CA9/2020 | EPI_ISL_425363 | 16/03/2020 |
| CAMB-74FDD | hCoV-19/England/CAMB-74FDD/2020 | EPI_ISL_440398 | 16/03/2020 |
| CAMB-75130 | hCoV-19/England/CAMB-75130/2020 | EPI_ISL_440400 | 16/03/2020 |
| CAMB-751A9 | hCoV-19/England/CAMB-751A9/2020 | EPI_ISL_440384 | 16/03/2020 |
| CAMB-75367 | hCoV-19/England/CAMB-75367/2020 | EPI_ISL_440382 | 16/03/2020 |
| CAMB-75455 | hCoV-19/England/CAMB-75455/2020 | EPI_ISL_425389 | 16/03/2020 |
| CAMB-754B7 | hCoV-19/England/CAMB-754B7/2020 | EPI_ISL_425395 | 16/03/2020 |
| CAMB-7565F | hCoV-19/England/CAMB-7565F/2020 | EPI_ISL_425418 | 16/03/2020 |
| CAMB-7567D | hCoV-19/England/CAMB-7567D/2020 | EPI_ISL_425420 | 16/03/2020 |
| CAMB-74BAC | hCoV-19/England/CAMB-74BAC/2020 | EPI_ISL_425352 | 17/03/2020 |
| CAMB-74BF7 | hCoV-19/England/CAMB-74BF7/2020 | EPI_ISL_425355 | 17/03/2020 |
| CAMB-74C12 | hCoV-19/England/CAMB-74C12/2020 | EPI_ISL_425357 | 17/03/2020 |
| CAMB-74CF4 | hCoV-19/England/CAMB-74CF4/2020 | EPI_ISL_425366 | 17/03/2020 |
| CAMB-74F82 | hCoV-19/England/CAMB-74F82/2020 | EPI_ISL_440392 | 17/03/2020 |
| CAMB-74F91 | hCoV-19/England/CAMB-74F91/2020 | EPI_ISL_440385 | 17/03/2020 |
| CAMB-75024 | hCoV-19/England/CAMB-75024/2020 | EPI_ISL_440401 | 17/03/2020 |
| CAMB-7522E | hCoV-19/England/CAMB-7522E/2020 | EPI_ISL_440422 | 17/03/2020 |
| CAMB-754DD | hCoV-19/England/CAMB-754DD/2020 | EPI_ISL_425397 | 17/03/2020 |
| CAMB-73E1D | hCoV-19/England/CAMB-73E1D/2020 | EPI_ISL_440330 | 18/03/2020 |
| CAMB-73FA1 | hCoV-19/England/CAMB-73FA1/2020 | EPI_ISL_440315 | 18/03/2020 |
| CAMB-74016 | hCoV-19/England/CAMB-74016/2020 | EPI_ISL_440303 | 18/03/2020 |
| CAMB-7419B | hCoV-19/England/CAMB-7419B/2020 | EPI_ISL_440328 | 18/03/2020 |
| CAMB-74395 | hCoV-19/England/CAMB-74395/2020 | EPI_ISL_425284 | 18/03/2020 |
| CAMB-749A2 | hCoV-19/England/CAMB-749A2/2020 | EPI_ISL_425341 | 18/03/2020 |
| CAMB-74A09 | hCoV-19/England/CAMB-74A09/2020 | EPI_ISL_425342 | 18/03/2020 |
| CAMB-74B33 | hCoV-19/England/CAMB-74B33/2020 | EPI_ISL_425347 | 18/03/2020 |
| CAMB-74B42 | hCoV-19/England/CAMB-74B42/2020 | EPI_ISL_425348 | 18/03/2020 |
| CAMB-74B60 | hCoV-19/England/CAMB-74B60/2020 | EPI_ISL_425350 | 18/03/2020 |
| CAMB-74E58 | hCoV-19/England/CAMB-74E58/2020 | EPI_ISL_425379 | 18/03/2020 |
| CAMB-74E85 | hCoV-19/England/CAMB-74E85/2020 | EPI_ISL_425382 | 18/03/2020 |
| CAMB-74F37 | hCoV-19/England/CAMB-74F37/2020 | EPI_ISL_440394 | 18/03/2020 |
| CAMB-7517C | hCoV-19/England/CAMB-7517C/2020 | EPI_ISL_440420 | 18/03/2020 |
| CAMB-75200 | hCoV-19/England/CAMB-75200/2020 | EPI_ISL_440434 | 18/03/2020 |
| CAMB-7566E | hCoV-19/England/CAMB-7566E/2020 | EPI_ISL_425419 | 18/03/2020 |
| CAMB-73E77 | hCoV-19/England/CAMB-73E77/2020 | EPI_ISL_440357 | 19/03/2020 |
| CAMB-73E86 | hCoV-19/England/CAMB-73E86/2020 | EPI_ISL_440316 | 19/03/2020 |
| CAMB-73F47 | hCoV-19/England/CAMB-73F47/2020 | EPI_ISL_440372 | 19/03/2020 |
| CAMB-73FB0 | hCoV-19/England/CAMB-73FB0/2020 | EPI_ISL_440355 | 19/03/2020 |
| CAMB-740CB | hCoV-19/England/CAMB-740CB/2020 | EPI_ISL_440306 | 19/03/2020 |
| CAMB-740F8 | hCoV-19/England/CAMB-740F8/2020 | EPI_ISL_440305 | 19/03/2020 |
| CAMB-74465 | hCoV-19/England/CAMB-74465/2020 | EPI_ISL_425292 | 19/03/2020 |
| CAMB-74B24 | hCoV-19/England/CAMB-74B24/2020 | EPI_ISL_425346 | 19/03/2020 |
| CAMB-74C5E | hCoV-19/England/CAMB-74C5E/2020 | EPI_ISL_425361 | 19/03/2020 |
| CAMB-74D2E | hCoV-19/England/CAMB-74D2E/2020 | EPI_ISL_425369 | 19/03/2020 |
| CAMB-74D3D | hCoV-19/England/CAMB-74D3D/2020 | EPI_ISL_425370 | 19/03/2020 |
| CAMB-74D6A | hCoV-19/England/CAMB-74D6A/2020 | EPI_ISL_425374 | 19/03/2020 |
| CAMB-75121 | hCoV-19/England/CAMB-75121/2020 | EPI_ISL_440435 | 19/03/2020 |
| CAMB-73527 | hCoV-19/England/CAMB-73527/2020 | EPI_ISL_440211 | 20/03/2020 |
| CAMB-73581 | hCoV-19/England/CAMB-73581/2020 | EPI_ISL_440147 | 20/03/2020 |
| CAMB-735AF | hCoV-19/England/CAMB-735AF/2020 | EPI_ISL_440164 | 20/03/2020 |
| CAMB-735CD | hCoV-19/England/CAMB-735CD/2020 | EPI_ISL_440073 | 20/03/2020 |
| CAMB-735DC | hCoV-19/England/CAMB-735DC/2020 | EPI_ISL_440111 | 20/03/2020 |
| CAMB-73F29 | hCoV-19/England/CAMB-73F29/2020 | EPI_ISL_440324 | 20/03/2020 |
| CAMB-74104 | hCoV-19/England/CAMB-74104/2020 | EPI_ISL_440347 | 20/03/2020 |
| CAMB-74140 | hCoV-19/England/CAMB-74140/2020 | EPI_ISL_440369 | 20/03/2020 |
| CAMB-741AA | hCoV-19/England/CAMB-741AA/2020 | EPI_ISL_440364 | 20/03/2020 |
| CAMB-7424D | hCoV-19/England/CAMB-7424D/2020 | EPI_ISL_440371 | 20/03/2020 |
| CAMB-7427A | hCoV-19/England/CAMB-7427A/2020 | EPI_ISL_440362 | 20/03/2020 |
| CAMB-74377 | hCoV-19/England/CAMB-74377/2020 | EPI_ISL_425282 | 20/03/2020 |
| CAMB-744A1 | hCoV-19/England/CAMB-744A1/2020 | EPI_ISL_425296 | 20/03/2020 |
| CAMB-758A4 | hCoV-19/England/CAMB-758A4/2020 | EPI_ISL_440351 | 20/03/2020 |
| CAMB-72476 | hCoV-19/England/CAMB-72476/2020 | EPI_ISL_439552 | 21/03/2020 |
| CAMB-724FE | hCoV-19/England/CAMB-724FE/2020 | EPI_ISL_439600 | 21/03/2020 |
| CAMB-72EB4 | hCoV-19/England/CAMB-72EB4/2020 | EPI_ISL_440080 | 21/03/2020 |
| CAMB-72FEE | hCoV-19/England/CAMB-72FEE/2020 | EPI_ISL_440144 | 21/03/2020 |
| CAMB-73017 | hCoV-19/England/CAMB-73017/2020 | EPI_ISL_440239 | 21/03/2020 |
| CAMB-73053 | hCoV-19/England/CAMB-73053/2020 | EPI_ISL_440166 | 21/03/2020 |
| CAMB-73071 | hCoV-19/England/CAMB-73071/2020 | EPI_ISL_440208 | 21/03/2020 |
| CAMB-73114 | hCoV-19/England/CAMB-73114/2020 | EPI_ISL_440240 | 21/03/2020 |
| CAMB-7317E | hCoV-19/England/CAMB-7317E/2020 | EPI_ISL_440264 | 21/03/2020 |
| CAMB-731AB | hCoV-19/England/CAMB-731AB/2020 | EPI_ISL_440203 | 21/03/2020 |
| CAMB-731D8 | hCoV-19/England/CAMB-731D8/2020 | EPI_ISL_440143 | 21/03/2020 |
| CAMB-731F6 | hCoV-19/England/CAMB-731F6/2020 | EPI_ISL_440282 | 21/03/2020 |
| CAMB-73211 | hCoV-19/England/CAMB-73211/2020 | EPI_ISL_440287 | 21/03/2020 |
| CAMB-7333C | hCoV-19/England/CAMB-7333C/2020 | EPI_ISL_440109 | 21/03/2020 |
| CAMB-734EE | hCoV-19/England/CAMB-734EE/2020 | EPI_ISL_440007 | 21/03/2020 |
| CAMB-736AC | hCoV-19/England/CAMB-736AC/2020 | EPI_ISL_440159 | 21/03/2020 |
| CAMB-7282F | hCoV-19/England/CAMB-7282F/2020 | EPI_ISL_439606 | 22/03/2020 |
| CAMB-728B6 | hCoV-19/England/CAMB-728B6/2020 | EPI_ISL_439941 | 22/03/2020 |
| CAMB-7292C | hCoV-19/England/CAMB-7292C/2020 | EPI_ISL_439909 | 22/03/2020 |
| CAMB-72AB0 | hCoV-19/England/CAMB-72AB0/2020 | EPI_ISL_439865 | 22/03/2020 |
| CAMB-72B44 | hCoV-19/England/CAMB-72B44/2020 | EPI_ISL_440064 | 22/03/2020 |
| CAMB-72BEA | hCoV-19/England/CAMB-72BEA/2020 | EPI_ISL_439996 | 22/03/2020 |
| CAMB-72C23 | hCoV-19/England/CAMB-72C23/2020 | EPI_ISL_439963 | 22/03/2020 |
| CAMB-72C41 | hCoV-19/England/CAMB-72C41/2020 | EPI_ISL_439975 | 22/03/2020 |
| CAMB-72C6F | hCoV-19/England/CAMB-72C6F/2020 | EPI_ISL_440046 | 22/03/2020 |
| CAMB-72C9C | hCoV-19/England/CAMB-72C9C/2020 | EPI_ISL_440040 | 22/03/2020 |
| CAMB-72D11 | hCoV-19/England/CAMB-72D11/2020 | EPI_ISL_440072 | 22/03/2020 |
| CAMB-72D7B | hCoV-19/England/CAMB-72D7B/2020 | EPI_ISL_440009 | 22/03/2020 |
| CAMB-72F93 | hCoV-19/England/CAMB-72F93/2020 | EPI_ISL_440263 | 22/03/2020 |
| CAMB-7319C | hCoV-19/England/CAMB-7319C/2020 | EPI_ISL_440271 | 22/03/2020 |
| CAMB-7323F | hCoV-19/England/CAMB-7323F/2020 | EPI_ISL_440217 | 22/03/2020 |
| CAMB-7327B | hCoV-19/England/CAMB-7327B/2020 | EPI_ISL_440184 | 22/03/2020 |
| CAMB-733A5 | hCoV-19/England/CAMB-733A5/2020 | EPI_ISL_440259 | 22/03/2020 |
| CAMB-733D2 | hCoV-19/England/CAMB-733D2/2020 | EPI_ISL_439961 | 22/03/2020 |
| CAMB-73712 | hCoV-19/England/CAMB-73712/2020 | EPI_ISL_440163 | 22/03/2020 |
| CAMB-74614 | hCoV-19/England/CAMB-74614/2020 | EPI_ISL_425316 | 22/03/2020 |
| CAMB-74623 | hCoV-19/England/CAMB-74623/2020 | EPI_ISL_425317 | 22/03/2020 |
| CAMB-72546 | hCoV-19/England/CAMB-72546/2020 | EPI_ISL_439541 | 23/03/2020 |
| CAMB-725EC | hCoV-19/England/CAMB-725EC/2020 | EPI_ISL_439898 | 23/03/2020 |
| CAMB-7269E | hCoV-19/England/CAMB-7269E/2020 | EPI_ISL_439897 | 23/03/2020 |
| CAMB-72740 | hCoV-19/England/CAMB-72740/2020 | EPI_ISL_439644 | 23/03/2020 |
| CAMB-727C8 | hCoV-19/England/CAMB-727C8/2020 | EPI_ISL_439538 | 23/03/2020 |
| CAMB-72889 | hCoV-19/England/CAMB-72889/2020 | EPI_ISL_439912 | 23/03/2020 |
| CAMB-72986 | hCoV-19/England/CAMB-72986/2020 | EPI_ISL_439566 | 23/03/2020 |
| CAMB-729A4 | hCoV-19/England/CAMB-729A4/2020 | EPI_ISL_439568 | 23/03/2020 |
| CAMB-72A1A | hCoV-19/England/CAMB-72A1A/2020 | EPI_ISL_439556 | 23/03/2020 |
| CAMB-72A56 | hCoV-19/England/CAMB-72A56/2020 | EPI_ISL_439868 | 23/03/2020 |
| CAMB-72B62 | hCoV-19/England/CAMB-72B62/2020 | EPI_ISL_440096 | 23/03/2020 |
| CAMB-72BAE | hCoV-19/England/CAMB-72BAE/2020 | EPI_ISL_440017 | 23/03/2020 |
| CAMB-72C14 | hCoV-19/England/CAMB-72C14/2020 | EPI_ISL_440077 | 23/03/2020 |
| CAMB-72E96 | hCoV-19/England/CAMB-72E96/2020 | EPI_ISL_440078 | 23/03/2020 |
| CAMB-72EA5 | hCoV-19/England/CAMB-72EA5/2020 | EPI_ISL_440074 | 23/03/2020 |
| CAMB-72F0C | hCoV-19/England/CAMB-72F0C/2020 | EPI_ISL_439954 | 23/03/2020 |
| CAMB-73008 | hCoV-19/England/CAMB-73008/2020 | EPI_ISL_440226 | 23/03/2020 |
| CAMB-730AE | hCoV-19/England/CAMB-730AE/2020 | EPI_ISL_440179 | 23/03/2020 |
| CAMB-731C9 | hCoV-19/England/CAMB-731C9/2020 | EPI_ISL_440230 | 23/03/2020 |
| CAMB-7243A | hCoV-19/England/CAMB-7243A/2020 | EPI_ISL_439584 | 24/03/2020 |
| CAMB-72458 | hCoV-19/England/CAMB-72458/2020 | EPI_ISL_439940 | 24/03/2020 |
| CAMB-72591 | hCoV-19/England/CAMB-72591/2020 | EPI_ISL_439889 | 24/03/2020 |
| CAMB-7233D | hCoV-19/England/CAMB-7233D/2020 | EPI_ISL_439651 | 25/03/2020 |
| CAMB-784CB | hCoV-19/England/CAMB-784CB/2020 | EPI_ISL_433741 | 26/03/2020 |
| CAMB-784E9 | hCoV-19/England/CAMB-784E9/2020 | EPI_ISL_433743 | 26/03/2020 |
| CAMB-79A04 | hCoV-19/England/CAMB-79A04/2020 | EPI_ISL_433813 | 26/03/2020 |
| CAMB-79A40 | hCoV-19/England/CAMB-79A40/2020 | EPI_ISL_433815 | 26/03/2020 |
| CAMB-72018 | hCoV-19/England/CAMB-72018/2020 | EPI_ISL_439548 | 27/03/2020 |
| CAMB-74517 | hCoV-19/England/CAMB-74517/2020 | EPI_ISL_425301 | 27/03/2020 |
| CAMB-74526 | hCoV-19/England/CAMB-74526/2020 | EPI_ISL_425302 | 27/03/2020 |
| CAMB-78504 | hCoV-19/England/CAMB-78504/2020 | EPI_ISL_433744 | 27/03/2020 |
| CAMB-79A9B | hCoV-19/England/CAMB-79A9B/2020 | EPI_ISL_433818 | 27/03/2020 |
| CAMB-79AB9 | hCoV-19/England/CAMB-79AB9/2020 | EPI_ISL_433819 | 27/03/2020 |
| CAMB-79AF5 | hCoV-19/England/CAMB-79AF5/2020 | EPI_ISL_433823 | 27/03/2020 |
| CAMB-71E6A | hCoV-19/England/CAMB-71E6A/2020 | EPI_ISL_439654 | 28/03/2020 |
| CAMB-71F67 | hCoV-19/England/CAMB-71F67/2020 | EPI_ISL_439936 | 28/03/2020 |
| CAMB-71FB2 | hCoV-19/England/CAMB-71FB2/2020 | EPI_ISL_439873 | 28/03/2020 |
| CAMB-72045 | hCoV-19/England/CAMB-72045/2020 | EPI_ISL_439631 | 28/03/2020 |
| CAMB-72063 | hCoV-19/England/CAMB-72063/2020 | EPI_ISL_439942 | 28/03/2020 |
| CAMB-72072 | hCoV-19/England/CAMB-72072/2020 | EPI_ISL_439894 | 28/03/2020 |
| CAMB-72160 | hCoV-19/England/CAMB-72160/2020 | EPI_ISL_439887 | 28/03/2020 |
| CAMB-74508 | hCoV-19/England/CAMB-74508/2020 | EPI_ISL_425300 | 28/03/2020 |
| CAMB-74571 | hCoV-19/England/CAMB-74571/2020 | EPI_ISL_425307 | 28/03/2020 |
| CAMB-75A71 | hCoV-19/England/CAMB-75A71/2020 | EPI_ISL_438388 | 28/03/2020 |
| CAMB-75B9C | hCoV-19/England/CAMB-75B9C/2020 | EPI_ISL_438439 | 28/03/2020 |
| CAMB-75BAB | hCoV-19/England/CAMB-75BAB/2020 | EPI_ISL_438307 | 28/03/2020 |
| CAMB-75C6C | hCoV-19/England/CAMB-75C6C/2020 | EPI_ISL_438463 | 28/03/2020 |
| CAMB-76005 | hCoV-19/England/CAMB-76005/2020 | EPI_ISL_438415 | 28/03/2020 |
| CAMB-762D2 | hCoV-19/England/CAMB-762D2/2020 | EPI_ISL_438362 | 28/03/2020 |
| CAMB-71E88 | hCoV-19/England/CAMB-71E88/2020 | EPI_ISL_439907 | 29/03/2020 |
| CAMB-71F49 | hCoV-19/England/CAMB-71F49/2020 | EPI_ISL_439626 | 29/03/2020 |
| CAMB-72054 | hCoV-19/England/CAMB-72054/2020 | EPI_ISL_439916 | 29/03/2020 |
| CAMB-7218E | hCoV-19/England/CAMB-7218E/2020 | EPI_ISL_439598 | 29/03/2020 |
| CAMB-73994 | hCoV-19/England/CAMB-73994/2020 | EPI_ISL_425260 | 29/03/2020 |
| CAMB-7468D | hCoV-19/England/CAMB-7468D/2020 | EPI_ISL_425321 | 29/03/2020 |
| CAMB-7469C | hCoV-19/England/CAMB-7469C/2020 | EPI_ISL_425322 | 29/03/2020 |
| CAMB-74711 | hCoV-19/England/CAMB-74711/2020 | EPI_ISL_425327 | 29/03/2020 |
| CAMB-74720 | hCoV-19/England/CAMB-74720/2020 | EPI_ISL_425328 | 29/03/2020 |
| CAMB-7473F | hCoV-19/England/CAMB-7473F/2020 | EPI_ISL_425329 | 29/03/2020 |
| CAMB-7474E | hCoV-19/England/CAMB-7474E/2020 | EPI_ISL_425330 | 29/03/2020 |
| CAMB-759FC | hCoV-19/England/CAMB-759FC/2020 | EPI_ISL_438258 | 29/03/2020 |
| CAMB-75ADB | hCoV-19/England/CAMB-75ADB/2020 | EPI_ISL_438448 | 29/03/2020 |
| CAMB-75B41 | hCoV-19/England/CAMB-75B41/2020 | EPI_ISL_438378 | 29/03/2020 |
| CAMB-75C8A | hCoV-19/England/CAMB-75C8A/2020 | EPI_ISL_438262 | 29/03/2020 |
| CAMB-75D1E | hCoV-19/England/CAMB-75D1E/2020 | EPI_ISL_438355 | 29/03/2020 |
| CAMB-75D2D | hCoV-19/England/CAMB-75D2D/2020 | EPI_ISL_438423 | 29/03/2020 |
| CAMB-75D4B | hCoV-19/England/CAMB-75D4B/2020 | EPI_ISL_438420 | 29/03/2020 |
| CAMB-75D69 | hCoV-19/England/CAMB-75D69/2020 | EPI_ISL_438284 | 29/03/2020 |
| CAMB-75D96 | hCoV-19/England/CAMB-75D96/2020 | EPI_ISL_438334 | 29/03/2020 |
| CAMB-75DD2 | hCoV-19/England/CAMB-75DD2/2020 | EPI_ISL_438270 | 29/03/2020 |
| CAMB-75F27 | hCoV-19/England/CAMB-75F27/2020 | EPI_ISL_438367 | 29/03/2020 |
| CAMB-75F36 | hCoV-19/England/CAMB-75F36/2020 | EPI_ISL_438271 | 29/03/2020 |
| CAMB-75F81 | hCoV-19/England/CAMB-75F81/2020 | EPI_ISL_438410 | 29/03/2020 |
| CAMB-762A5 | hCoV-19/England/CAMB-762A5/2020 | EPI_ISL_438437 | 29/03/2020 |
| CAMB-764CD | hCoV-19/England/CAMB-764CD/2020 | EPI_ISL_438429 | 29/03/2020 |
| CAMB-765CA | hCoV-19/England/CAMB-765CA/2020 | EPI_ISL_439520 | 29/03/2020 |
| CAMB-76612 | hCoV-19/England/CAMB-76612/2020 | EPI_ISL_438357 | 29/03/2020 |
| CAMB-76894 | hCoV-19/England/CAMB-76894/2020 | EPI_ISL_438286 | 29/03/2020 |
| CAMB-768B2 | hCoV-19/England/CAMB-768B2/2020 | EPI_ISL_438347 | 29/03/2020 |
| CAMB-76973 | hCoV-19/England/CAMB-76973/2020 | EPI_ISL_438449 | 29/03/2020 |
| CAMB-76ABC | hCoV-19/England/CAMB-76ABC/2020 | EPI_ISL_438309 | 29/03/2020 |
| CAMB-76C4D | hCoV-19/England/CAMB-76C4D/2020 | EPI_ISL_438281 | 29/03/2020 |
| CAMB-71B54 | hCoV-19/England/CAMB-71B54/2020 | EPI_ISL_439664 | 30/03/2020 |
| CAMB-71C8E | hCoV-19/England/CAMB-71C8E/2020 | EPI_ISL_439567 | 30/03/2020 |
| CAMB-71CBB | hCoV-19/England/CAMB-71CBB/2020 | EPI_ISL_439947 | 30/03/2020 |
| CAMB-71D30 | hCoV-19/England/CAMB-71D30/2020 | EPI_ISL_439554 | 30/03/2020 |
| CAMB-737B8 | hCoV-19/England/CAMB-737B8/2020 | EPI_ISL_425238 | 30/03/2020 |
| CAMB-73A82 | hCoV-19/England/CAMB-73A82/2020 | EPI_ISL_425273 | 30/03/2020 |
| CAMB-75B7E | hCoV-19/England/CAMB-75B7E/2020 | EPI_ISL_438278 | 30/03/2020 |
| CAMB-75CF3 | hCoV-19/England/CAMB-75CF3/2020 | EPI_ISL_438451 | 30/03/2020 |
| CAMB-76032 | hCoV-19/England/CAMB-76032/2020 | EPI_ISL_439507 | 30/03/2020 |
| CAMB-760F6 | hCoV-19/England/CAMB-760F6/2020 | EPI_ISL_438268 | 30/03/2020 |
| CAMB-76102 | hCoV-19/England/CAMB-76102/2020 | EPI_ISL_439442 | 30/03/2020 |
| CAMB-7614E | hCoV-19/England/CAMB-7614E/2020 | EPI_ISL_439469 | 30/03/2020 |
| CAMB-761D5 | hCoV-19/England/CAMB-761D5/2020 | EPI_ISL_438452 | 30/03/2020 |
| CAMB-762E1 | hCoV-19/England/CAMB-762E1/2020 | EPI_ISL_438413 | 30/03/2020 |
| CAMB-76393 | hCoV-19/England/CAMB-76393/2020 | EPI_ISL_438274 | 30/03/2020 |
| CAMB-763A2 | hCoV-19/England/CAMB-763A2/2020 | EPI_ISL_438400 | 30/03/2020 |
| CAMB-763EE | hCoV-19/England/CAMB-763EE/2020 | EPI_ISL_438354 | 30/03/2020 |
| CAMB-76445 | hCoV-19/England/CAMB-76445/2020 | EPI_ISL_438273 | 30/03/2020 |
| CAMB-76454 | hCoV-19/England/CAMB-76454/2020 | EPI_ISL_438287 | 30/03/2020 |
| CAMB-76472 | hCoV-19/England/CAMB-76472/2020 | EPI_ISL_438526 | 30/03/2020 |
| CAMB-76481 | hCoV-19/England/CAMB-76481/2020 | EPI_ISL_438267 | 30/03/2020 |
| CAMB-7658E | hCoV-19/England/CAMB-7658E/2020 | EPI_ISL_439515 | 30/03/2020 |
| CAMB-765BB | hCoV-19/England/CAMB-765BB/2020 | EPI_ISL_438538 | 30/03/2020 |
| CAMB-766E5 | hCoV-19/England/CAMB-766E5/2020 | EPI_ISL_438506 | 30/03/2020 |
| CAMB-767C4 | hCoV-19/England/CAMB-767C4/2020 | EPI_ISL_438427 | 30/03/2020 |
| CAMB-7680D | hCoV-19/England/CAMB-7680D/2020 | EPI_ISL_438457 | 30/03/2020 |
| CAMB-7681C | hCoV-19/England/CAMB-7681C/2020 | EPI_ISL_438487 | 30/03/2020 |
| CAMB-768C1 | hCoV-19/England/CAMB-768C1/2020 | EPI_ISL_438443 | 30/03/2020 |
| CAMB-76991 | hCoV-19/England/CAMB-76991/2020 | EPI_ISL_438424 | 30/03/2020 |
| CAMB-769A0 | hCoV-19/England/CAMB-769A0/2020 | EPI_ISL_438488 | 30/03/2020 |
| CAMB-769EC | hCoV-19/England/CAMB-769EC/2020 | EPI_ISL_438359 | 30/03/2020 |
| CAMB-76AE9 | hCoV-19/England/CAMB-76AE9/2020 | EPI_ISL_438465 | 30/03/2020 |
| CAMB-76B40 | hCoV-19/England/CAMB-76B40/2020 | EPI_ISL_438372 | 30/03/2020 |
| CAMB-76BF5 | hCoV-19/England/CAMB-76BF5/2020 | EPI_ISL_438516 | 30/03/2020 |
| CAMB-76C10 | hCoV-19/England/CAMB-76C10/2020 | EPI_ISL_439398 | 30/03/2020 |
| CAMB-76C6B | hCoV-19/England/CAMB-76C6B/2020 | EPI_ISL_438257 | 30/03/2020 |
| CAMB-76D0E | hCoV-19/England/CAMB-76D0E/2020 | EPI_ISL_438479 | 30/03/2020 |
| CAMB-76DD1 | hCoV-19/England/CAMB-76DD1/2020 | EPI_ISL_425429 | 30/03/2020 |
| CAMB-76DFF | hCoV-19/England/CAMB-76DFF/2020 | EPI_ISL_425430 | 30/03/2020 |
| CAMB-770C8 | hCoV-19/England/CAMB-770C8/2020 | EPI_ISL_425461 | 30/03/2020 |
| CAMB-77286 | hCoV-19/England/CAMB-77286/2020 | EPI_ISL_439385 | 30/03/2020 |
| CAMB-7731A | hCoV-19/England/CAMB-7731A/2020 | EPI_ISL_439384 | 30/03/2020 |
| CAMB-77796 | hCoV-19/England/CAMB-77796/2020 | EPI_ISL_439500 | 30/03/2020 |
| CAMB-71AEE | hCoV-19/England/CAMB-71AEE/2020 | EPI_ISL_439578 | 31/03/2020 |
| CAMB-71AFD | hCoV-19/England/CAMB-71AFD/2020 | EPI_ISL_439929 | 31/03/2020 |
| CAMB-71B09 | hCoV-19/England/CAMB-71B09/2020 | EPI_ISL_439950 | 31/03/2020 |
| CAMB-71B18 | hCoV-19/England/CAMB-71B18/2020 | EPI_ISL_439619 | 31/03/2020 |
| CAMB-71C42 | hCoV-19/England/CAMB-71C42/2020 | EPI_ISL_439880 | 31/03/2020 |
| CAMB-71C60 | hCoV-19/England/CAMB-71C60/2020 | EPI_ISL_439607 | 31/03/2020 |
| CAMB-71C9D | hCoV-19/England/CAMB-71C9D/2020 | EPI_ISL_439637 | 31/03/2020 |
| CAMB-737C7 | hCoV-19/England/CAMB-737C7/2020 | EPI_ISL_425239 | 31/03/2020 |
| CAMB-737D6 | hCoV-19/England/CAMB-737D6/2020 | EPI_ISL_425240 | 31/03/2020 |
| CAMB-739B2 | hCoV-19/England/CAMB-739B2/2020 | EPI_ISL_425262 | 31/03/2020 |
| CAMB-73A64 | hCoV-19/England/CAMB-73A64/2020 | EPI_ISL_425271 | 31/03/2020 |
| CAMB-75929 | hCoV-19/England/CAMB-75929/2020 | EPI_ISL_438291 | 31/03/2020 |
| CAMB-75956 | hCoV-19/England/CAMB-75956/2020 | EPI_ISL_438483 | 31/03/2020 |
| CAMB-759A1 | hCoV-19/England/CAMB-759A1/2020 | EPI_ISL_438507 | 31/03/2020 |
| CAMB-76AF8 | hCoV-19/England/CAMB-76AF8/2020 | EPI_ISL_438473 | 31/03/2020 |
| CAMB-76B31 | hCoV-19/England/CAMB-76B31/2020 | EPI_ISL_438398 | 31/03/2020 |
| CAMB-76B9B | hCoV-19/England/CAMB-76B9B/2020 | EPI_ISL_438265 | 31/03/2020 |
| CAMB-76C7A | hCoV-19/England/CAMB-76C7A/2020 | EPI_ISL_438459 | 31/03/2020 |
| CAMB-76E47 | hCoV-19/England/CAMB-76E47/2020 | EPI_ISL_439415 | 31/03/2020 |
| CAMB-76FAE | hCoV-19/England/CAMB-76FAE/2020 | EPI_ISL_425444 | 31/03/2020 |
| CAMB-76FEA | hCoV-19/England/CAMB-76FEA/2020 | EPI_ISL_425448 | 31/03/2020 |
| CAMB-77022 | hCoV-19/England/CAMB-77022/2020 | EPI_ISL_425452 | 31/03/2020 |
| CAMB-77040 | hCoV-19/England/CAMB-77040/2020 | EPI_ISL_425454 | 31/03/2020 |
| CAMB-7705F | hCoV-19/England/CAMB-7705F/2020 | EPI_ISL_425455 | 31/03/2020 |
| CAMB-770E6 | hCoV-19/England/CAMB-770E6/2020 | EPI_ISL_439380 | 31/03/2020 |
| CAMB-771B6 | hCoV-19/England/CAMB-771B6/2020 | EPI_ISL_439492 | 31/03/2020 |
| CAMB-77277 | hCoV-19/England/CAMB-77277/2020 | EPI_ISL_438509 | 31/03/2020 |
| CAMB-772A4 | hCoV-19/England/CAMB-772A4/2020 | EPI_ISL_439428 | 31/03/2020 |
| CAMB-772B3 | hCoV-19/England/CAMB-772B3/2020 | EPI_ISL_439381 | 31/03/2020 |
| CAMB-772D1 | hCoV-19/England/CAMB-772D1/2020 | EPI_ISL_439420 | 31/03/2020 |
| CAMB-77365 | hCoV-19/England/CAMB-77365/2020 | EPI_ISL_439513 | 31/03/2020 |
| CAMB-77383 | hCoV-19/England/CAMB-77383/2020 | EPI_ISL_438544 | 31/03/2020 |
| CAMB-77392 | hCoV-19/England/CAMB-77392/2020 | EPI_ISL_439456 | 31/03/2020 |
| CAMB-773A1 | hCoV-19/England/CAMB-773A1/2020 | EPI_ISL_438527 | 31/03/2020 |
| CAMB-773CF | hCoV-19/England/CAMB-773CF/2020 | EPI_ISL_439463 | 31/03/2020 |
| CAMB-77471 | hCoV-19/England/CAMB-77471/2020 | EPI_ISL_438535 | 31/03/2020 |
| CAMB-7749F | hCoV-19/England/CAMB-7749F/2020 | EPI_ISL_439424 | 31/03/2020 |
| CAMB-774AE | hCoV-19/England/CAMB-774AE/2020 | EPI_ISL_439451 | 31/03/2020 |
| CAMB-774F9 | hCoV-19/England/CAMB-774F9/2020 | EPI_ISL_439416 | 31/03/2020 |
| CAMB-77514 | hCoV-19/England/CAMB-77514/2020 | EPI_ISL_438517 | 31/03/2020 |
| CAMB-7766C | hCoV-19/England/CAMB-7766C/2020 | EPI_ISL_439368 | 31/03/2020 |
| CAMB-7767B | hCoV-19/England/CAMB-7767B/2020 | EPI_ISL_439430 | 31/03/2020 |
| CAMB-776D5 | hCoV-19/England/CAMB-776D5/2020 | EPI_ISL_439413 | 31/03/2020 |
| CAMB-7774B | hCoV-19/England/CAMB-7774B/2020 | EPI_ISL_439496 | 31/03/2020 |
| CAMB-777A5 | hCoV-19/England/CAMB-777A5/2020 | EPI_ISL_439404 | 31/03/2020 |
| CAMB-777E1 | hCoV-19/England/CAMB-777E1/2020 | EPI_ISL_439481 | 31/03/2020 |
| CAMB-77893 | hCoV-19/England/CAMB-77893/2020 | EPI_ISL_439509 | 31/03/2020 |
| CAMB-778B1 | hCoV-19/England/CAMB-778B1/2020 | EPI_ISL_439441 | 31/03/2020 |
| CAMB-778DF | hCoV-19/England/CAMB-778DF/2020 | EPI_ISL_439445 | 31/03/2020 |
| CAMB-77954 | hCoV-19/England/CAMB-77954/2020 | EPI_ISL_438525 | 31/03/2020 |
| CAMB-77990 | hCoV-19/England/CAMB-77990/2020 | EPI_ISL_439524 | 31/03/2020 |
| CAMB-77A9D | hCoV-19/England/CAMB-77A9D/2020 | EPI_ISL_439464 | 31/03/2020 |
| CAMB-77AAC | hCoV-19/England/CAMB-77AAC/2020 | EPI_ISL_591093 | 31/03/2020 |
| CAMB-7809A | hCoV-19/England/CAMB-7809A/2020 | EPI_ISL_441152 | 31/03/2020 |
| CAMB-73800 | hCoV-19/England/CAMB-73800/2020 | EPI_ISL_425243 | 01/04/2020 |
| CAMB-739D0 | hCoV-19/England/CAMB-739D0/2020 | EPI_ISL_425263 | 01/04/2020 |
| CAMB-73D2F | hCoV-19/England/CAMB-73D2F/2020 | EPI_ISL_433688 | 01/04/2020 |
| CAMB-759CF | hCoV-19/England/CAMB-759CF/2020 | EPI_ISL_591082 | 01/04/2020 |
| CAMB-76E65 | hCoV-19/England/CAMB-76E65/2020 | EPI_ISL_439389 | 01/04/2020 |
| CAMB-76FCC | hCoV-19/England/CAMB-76FCC/2020 | EPI_ISL_425446 | 01/04/2020 |
| CAMB-770F5 | hCoV-19/England/CAMB-770F5/2020 | EPI_ISL_439397 | 01/04/2020 |
| CAMB-7717A | hCoV-19/England/CAMB-7717A/2020 | EPI_ISL_439459 | 01/04/2020 |
| CAMB-77198 | hCoV-19/England/CAMB-77198/2020 | EPI_ISL_438536 | 01/04/2020 |
| CAMB-77374 | hCoV-19/England/CAMB-77374/2020 | EPI_ISL_439378 | 01/04/2020 |
| CAMB-7768A | hCoV-19/England/CAMB-7768A/2020 | EPI_ISL_439443 | 01/04/2020 |
| CAMB-777F0 | hCoV-19/England/CAMB-777F0/2020 | EPI_ISL_438514 | 01/04/2020 |
| CAMB-77A33 | hCoV-19/England/CAMB-77A33/2020 | EPI_ISL_439372 | 01/04/2020 |
| CAMB-77A51 | hCoV-19/England/CAMB-77A51/2020 | EPI_ISL_439401 | 01/04/2020 |
| CAMB-77ABB | hCoV-19/England/CAMB-77ABB/2020 | EPI_ISL_439435 | 01/04/2020 |
| CAMB-77D2B | hCoV-19/England/CAMB-77D2B/2020 | EPI_ISL_440601 | 01/04/2020 |
| CAMB-77DB2 | hCoV-19/England/CAMB-77DB2/2020 | EPI_ISL_440831 | 01/04/2020 |
| CAMB-78012 | hCoV-19/England/CAMB-78012/2020 | EPI_ISL_441123 | 01/04/2020 |
| CAMB-7804F | hCoV-19/England/CAMB-7804F/2020 | EPI_ISL_441803 | 01/04/2020 |
| CAMB-7812E | hCoV-19/England/CAMB-7812E/2020 | EPI_ISL_441149 | 01/04/2020 |
| CAMB-79785 | hCoV-19/England/CAMB-79785/2020 | EPI_ISL_441121 | 01/04/2020 |
| CAMB-7980A | hCoV-19/England/CAMB-7980A/2020 | EPI_ISL_441841 | 01/04/2020 |
| CAMB-79855 | hCoV-19/England/CAMB-79855/2020 | EPI_ISL_441137 | 01/04/2020 |
| CAMB-79864 | hCoV-19/England/CAMB-79864/2020 | EPI_ISL_441811 | 01/04/2020 |
| CAMB-79873 | hCoV-19/England/CAMB-79873/2020 | EPI_ISL_441788 | 01/04/2020 |
| CAMB-798CE | hCoV-19/England/CAMB-798CE/2020 | EPI_ISL_441796 | 01/04/2020 |
| CAMB-79907 | hCoV-19/England/CAMB-79907/2020 | EPI_ISL_441307 | 01/04/2020 |
| CAMB-79BD4 | hCoV-19/England/CAMB-79BD4/2020 | EPI_ISL_441241 | 01/04/2020 |
| CAMB-79C1D | hCoV-19/England/CAMB-79C1D/2020 | EPI_ISL_591081 | 01/04/2020 |
| CAMB-79E9F | hCoV-19/England/CAMB-79E9F/2020 | EPI_ISL_440617 | 01/04/2020 |
| CAMB-7A0E3 | hCoV-19/England/CAMB-7A0E3/2020 | EPI_ISL_441290 | 01/04/2020 |
| CAMB-7A292 | hCoV-19/England/CAMB-7A292/2020 | EPI_ISL_441304 | 01/04/2020 |
| CAMB-7A2FC | hCoV-19/England/CAMB-7A2FC/2020 | EPI_ISL_441844 | 01/04/2020 |
| CAMB-7A4F6 | hCoV-19/England/CAMB-7A4F6/2020 | EPI_ISL_441235 | 01/04/2020 |
| CAMB-7A511 | hCoV-19/England/CAMB-7A511/2020 | EPI_ISL_441257 | 01/04/2020 |
| CAMB-7A55D | hCoV-19/England/CAMB-7A55D/2020 | EPI_ISL_441068 | 01/04/2020 |
| CAMB-73D01 | hCoV-19/England/CAMB-73D01/2020 | EPI_ISL_433686 | 02/04/2020 |
| CAMB-73D10 | hCoV-19/England/CAMB-73D10/2020 | EPI_ISL_433687 | 02/04/2020 |
| CAMB-73D98 | hCoV-19/England/CAMB-73D98/2020 | EPI_ISL_433695 | 02/04/2020 |
| CAMB-76F35 | hCoV-19/England/CAMB-76F35/2020 | EPI_ISL_425440 | 02/04/2020 |
| CAMB-7720E | hCoV-19/England/CAMB-7720E/2020 | EPI_ISL_439383 | 02/04/2020 |
| CAMB-78D39 | hCoV-19/England/CAMB-78D39/2020 | EPI_ISL_441216 | 02/04/2020 |
| CAMB-78DB1 | hCoV-19/England/CAMB-78DB1/2020 | EPI_ISL_441115 | 02/04/2020 |
| CAMB-78E72 | hCoV-19/England/CAMB-78E72/2020 | EPI_ISL_441330 | 02/04/2020 |
| CAMB-79767 | hCoV-19/England/CAMB-79767/2020 | EPI_ISL_441143 | 02/04/2020 |
| CAMB-797C1 | hCoV-19/England/CAMB-797C1/2020 | EPI_ISL_441217 | 02/04/2020 |
| CAMB-79B89 | hCoV-19/England/CAMB-79B89/2020 | EPI_ISL_440621 | 02/04/2020 |
| CAMB-79C0E | hCoV-19/England/CAMB-79C0E/2020 | EPI_ISL_441284 | 02/04/2020 |
| CAMB-79C59 | hCoV-19/England/CAMB-79C59/2020 | EPI_ISL_441233 | 02/04/2020 |
| CAMB-79CB3 | hCoV-19/England/CAMB-79CB3/2020 | EPI_ISL_441301 | 02/04/2020 |
| CAMB-79CE0 | hCoV-19/England/CAMB-79CE0/2020 | EPI_ISL_440576 | 02/04/2020 |
| CAMB-79D38 | hCoV-19/England/CAMB-79D38/2020 | EPI_ISL_440615 | 02/04/2020 |
| CAMB-79E80 | hCoV-19/England/CAMB-79E80/2020 | EPI_ISL_441258 | 02/04/2020 |
| CAMB-79F32 | hCoV-19/England/CAMB-79F32/2020 | EPI_ISL_441247 | 02/04/2020 |
| CAMB-79FD8 | hCoV-19/England/CAMB-79FD8/2020 | EPI_ISL_441238 | 02/04/2020 |
| CAMB-79FF6 | hCoV-19/England/CAMB-79FF6/2020 | EPI_ISL_440600 | 02/04/2020 |
| CAMB-7A010 | hCoV-19/England/CAMB-7A010/2020 | EPI_ISL_441324 | 02/04/2020 |
| CAMB-7A03E | hCoV-19/England/CAMB-7A03E/2020 | EPI_ISL_441309 | 02/04/2020 |
| CAMB-7A04D | hCoV-19/England/CAMB-7A04D/2020 | EPI_ISL_441337 | 02/04/2020 |
| CAMB-7A07A | hCoV-19/England/CAMB-7A07A/2020 | EPI_ISL_441261 | 02/04/2020 |
| CAMB-7A098 | hCoV-19/England/CAMB-7A098/2020 | EPI_ISL_440610 | 02/04/2020 |
| CAMB-7A0C5 | hCoV-19/England/CAMB-7A0C5/2020 | EPI_ISL_440612 | 02/04/2020 |
| CAMB-7A0F2 | hCoV-19/England/CAMB-7A0F2/2020 | EPI_ISL_441300 | 02/04/2020 |
| CAMB-7A14A | hCoV-19/England/CAMB-7A14A/2020 | EPI_ISL_441289 | 02/04/2020 |
| CAMB-7A21A | hCoV-19/England/CAMB-7A21A/2020 | EPI_ISL_440583 | 02/04/2020 |
| CAMB-7A405 | hCoV-19/England/CAMB-7A405/2020 | EPI_ISL_441334 | 02/04/2020 |
| CAMB-7A4BA | hCoV-19/England/CAMB-7A4BA/2020 | EPI_ISL_441275 | 02/04/2020 |
| CAMB-7A5A8 | hCoV-19/England/CAMB-7A5A8/2020 | EPI_ISL_441825 | 02/04/2020 |
| CAMB-7A5F3 | hCoV-19/England/CAMB-7A5F3/2020 | EPI_ISL_441845 | 02/04/2020 |
| CAMB-7A739 | hCoV-19/England/CAMB-7A739/2020 | EPI_ISL_441135 | 02/04/2020 |
| CAMB-7A793 | hCoV-19/England/CAMB-7A793/2020 | EPI_ISL_441271 | 02/04/2020 |
| CAMB-7A7C0 | hCoV-19/England/CAMB-7A7C0/2020 | EPI_ISL_440570 | 02/04/2020 |
| CAMB-7A7DF | hCoV-19/England/CAMB-7A7DF/2020 | EPI_ISL_441294 | 02/04/2020 |
| CAMB-7A7FD | hCoV-19/England/CAMB-7A7FD/2020 | EPI_ISL_440591 | 02/04/2020 |
| CAMB-7A836 | hCoV-19/England/CAMB-7A836/2020 | EPI_ISL_441345 | 02/04/2020 |
| CAMB-7A881 | hCoV-19/England/CAMB-7A881/2020 | EPI_ISL_441069 | 02/04/2020 |
| CAMB-7A890 | hCoV-19/England/CAMB-7A890/2020 | EPI_ISL_441172 | 02/04/2020 |
| CAMB-7A99D | hCoV-19/England/CAMB-7A99D/2020 | EPI_ISL_441344 | 02/04/2020 |
| CAMB-7A9E8 | hCoV-19/England/CAMB-7A9E8/2020 | EPI_ISL_440565 | 02/04/2020 |
| CAMB-7B7ED | hCoV-19/England/CAMB-7B7ED/2020 | EPI_ISL_441591 | 02/04/2020 |
| CAMB-7D29F | hCoV-19/England/CAMB-7D29F/2020 | EPI_ISL_442073 | 02/04/2020 |
| CAMB-73B16 | hCoV-19/England/CAMB-73B16/2020 | EPI_ISL_433666 | 03/04/2020 |
| CAMB-73B9E | hCoV-19/England/CAMB-73B9E/2020 | EPI_ISL_433670 | 03/04/2020 |
| CAMB-73BDA | hCoV-19/England/CAMB-73BDA/2020 | EPI_ISL_433674 | 03/04/2020 |
| CAMB-77B9A | hCoV-19/England/CAMB-77B9A/2020 | EPI_ISL_440830 | 03/04/2020 |
| CAMB-78346 | hCoV-19/England/CAMB-78346/2020 | EPI_ISL_433720 | 03/04/2020 |
| CAMB-78364 | hCoV-19/England/CAMB-78364/2020 | EPI_ISL_433722 | 03/04/2020 |
| CAMB-78B02 | hCoV-19/England/CAMB-78B02/2020 | EPI_ISL_441111 | 03/04/2020 |
| CAMB-78B11 | hCoV-19/England/CAMB-78B11/2020 | EPI_ISL_441808 | 03/04/2020 |
| CAMB-78B3F | hCoV-19/England/CAMB-78B3F/2020 | EPI_ISL_441062 | 03/04/2020 |
| CAMB-78B99 | hCoV-19/England/CAMB-78B99/2020 | EPI_ISL_441054 | 03/04/2020 |
| CAMB-78CA5 | hCoV-19/England/CAMB-78CA5/2020 | EPI_ISL_441220 | 03/04/2020 |
| CAMB-78D48 | hCoV-19/England/CAMB-78D48/2020 | EPI_ISL_441336 | 03/04/2020 |
| CAMB-78D75 | hCoV-19/England/CAMB-78D75/2020 | EPI_ISL_441804 | 03/04/2020 |
| CAMB-78D93 | hCoV-19/England/CAMB-78D93/2020 | EPI_ISL_441084 | 03/04/2020 |
| CAMB-78FD9 | hCoV-19/England/CAMB-78FD9/2020 | EPI_ISL_441798 | 03/04/2020 |
| CAMB-790E4 | hCoV-19/England/CAMB-790E4/2020 | EPI_ISL_441839 | 03/04/2020 |
| CAMB-7912D | hCoV-19/England/CAMB-7912D/2020 | EPI_ISL_441159 | 03/04/2020 |
| CAMB-7913C | hCoV-19/England/CAMB-7913C/2020 | EPI_ISL_441081 | 03/04/2020 |
| CAMB-791A5 | hCoV-19/England/CAMB-791A5/2020 | EPI_ISL_441113 | 03/04/2020 |
| CAMB-79318 | hCoV-19/England/CAMB-79318/2020 | EPI_ISL_470536 | 03/04/2020 |
| CAMB-79336 | hCoV-19/England/CAMB-79336/2020 | EPI_ISL_441171 | 03/04/2020 |
| CAMB-793DC | hCoV-19/England/CAMB-793DC/2020 | EPI_ISL_441075 | 03/04/2020 |
| CAMB-79415 | hCoV-19/England/CAMB-79415/2020 | EPI_ISL_441147 | 03/04/2020 |
| CAMB-7957C | hCoV-19/England/CAMB-7957C/2020 | EPI_ISL_441200 | 03/04/2020 |
| CAMB-79679 | hCoV-19/England/CAMB-79679/2020 | EPI_ISL_441815 | 03/04/2020 |
| CAMB-796B5 | hCoV-19/England/CAMB-796B5/2020 | EPI_ISL_441214 | 03/04/2020 |
| CAMB-7A001 | hCoV-19/England/CAMB-7A001/2020 | EPI_ISL_440573 | 03/04/2020 |
| CAMB-7A12C | hCoV-19/England/CAMB-7A12C/2020 | EPI_ISL_441244 | 03/04/2020 |
| CAMB-7A159 | hCoV-19/England/CAMB-7A159/2020 | EPI_ISL_440586 | 03/04/2020 |
| CAMB-7A775 | hCoV-19/England/CAMB-7A775/2020 | EPI_ISL_440578 | 03/04/2020 |
| CAMB-7A98E | hCoV-19/England/CAMB-7A98E/2020 | EPI_ISL_441205 | 03/04/2020 |
| CAMB-7A9AC | hCoV-19/England/CAMB-7A9AC/2020 | EPI_ISL_441234 | 03/04/2020 |
| CAMB-7A9CA | hCoV-19/England/CAMB-7A9CA/2020 | EPI_ISL_440580 | 03/04/2020 |
| CAMB-7B64A | hCoV-19/England/CAMB-7B64A/2020 | EPI_ISL_441122 | 03/04/2020 |
| CAMB-7B6A4 | hCoV-19/England/CAMB-7B6A4/2020 | EPI_ISL_441087 | 03/04/2020 |
| CAMB-7B6E0 | hCoV-19/England/CAMB-7B6E0/2020 | EPI_ISL_441654 | 03/04/2020 |
| CAMB-7B6FF | hCoV-19/England/CAMB-7B6FF/2020 | EPI_ISL_442259 | 03/04/2020 |
| CAMB-7B70B | hCoV-19/England/CAMB-7B70B/2020 | EPI_ISL_442312 | 03/04/2020 |
| CAMB-7B747 | hCoV-19/England/CAMB-7B747/2020 | EPI_ISL_441650 | 03/04/2020 |
| CAMB-7B783 | hCoV-19/England/CAMB-7B783/2020 | EPI_ISL_442283 | 03/04/2020 |
| CAMB-7B7CF | hCoV-19/England/CAMB-7B7CF/2020 | EPI_ISL_442218 | 03/04/2020 |
| CAMB-7B7FC | hCoV-19/England/CAMB-7B7FC/2020 | EPI_ISL_442331 | 03/04/2020 |
| CAMB-7BA7B | hCoV-19/England/CAMB-7BA7B/2020 | EPI_ISL_442315 | 03/04/2020 |
| CAMB-7BB1E | hCoV-19/England/CAMB-7BB1E/2020 | EPI_ISL_441593 | 03/04/2020 |
| CAMB-781A6 | hCoV-19/England/CAMB-781A6/2020 | EPI_ISL_433703 | 04/04/2020 |
| CAMB-78328 | hCoV-19/England/CAMB-78328/2020 | EPI_ISL_433718 | 04/04/2020 |
| CAMB-78337 | hCoV-19/England/CAMB-78337/2020 | EPI_ISL_433719 | 04/04/2020 |
| CAMB-795F4 | hCoV-19/England/CAMB-795F4/2020 | EPI_ISL_441102 | 04/04/2020 |
| CAMB-796F1 | hCoV-19/England/CAMB-796F1/2020 | EPI_ISL_441215 | 04/04/2020 |
| CAMB-7970D | hCoV-19/England/CAMB-7970D/2020 | EPI_ISL_441148 | 04/04/2020 |
| CAMB-7B8F9 | hCoV-19/England/CAMB-7B8F9/2020 | EPI_ISL_442174 | 04/04/2020 |
| CAMB-7B9BA | hCoV-19/England/CAMB-7B9BA/2020 | EPI_ISL_442336 | 04/04/2020 |
| CAMB-7B9C9 | hCoV-19/England/CAMB-7B9C9/2020 | EPI_ISL_442060 | 04/04/2020 |
| CAMB-7BA5D | hCoV-19/England/CAMB-7BA5D/2020 | EPI_ISL_442165 | 04/04/2020 |
| CAMB-7BA6C | hCoV-19/England/CAMB-7BA6C/2020 | EPI_ISL_441609 | 04/04/2020 |
| CAMB-7BAC6 | hCoV-19/England/CAMB-7BAC6/2020 | EPI_ISL_441557 | 04/04/2020 |
| CAMB-7BB3C | hCoV-19/England/CAMB-7BB3C/2020 | EPI_ISL_442227 | 04/04/2020 |
| CAMB-7BC1B | hCoV-19/England/CAMB-7BC1B/2020 | EPI_ISL_442207 | 04/04/2020 |
| CAMB-7BC57 | hCoV-19/England/CAMB-7BC57/2020 | EPI_ISL_442176 | 04/04/2020 |
| CAMB-7BCB1 | hCoV-19/England/CAMB-7BCB1/2020 | EPI_ISL_442087 | 04/04/2020 |
| CAMB-7BF30 | hCoV-19/England/CAMB-7BF30/2020 | EPI_ISL_442101 | 04/04/2020 |
| CAMB-7C412 | hCoV-19/England/CAMB-7C412/2020 | EPI_ISL_441584 | 04/04/2020 |
| CAMB-7C47C | hCoV-19/England/CAMB-7C47C/2020 | EPI_ISL_442278 | 04/04/2020 |
| CAMB-7C4B8 | hCoV-19/England/CAMB-7C4B8/2020 | EPI_ISL_442335 | 04/04/2020 |
| CAMB-7C4F4 | hCoV-19/England/CAMB-7C4F4/2020 | EPI_ISL_442248 | 04/04/2020 |
| CAMB-7C5E2 | hCoV-19/England/CAMB-7C5E2/2020 | EPI_ISL_442286 | 04/04/2020 |
| CAMB-7C6C1 | hCoV-19/England/CAMB-7C6C1/2020 | EPI_ISL_441628 | 04/04/2020 |
| CAMB-7C6FE | hCoV-19/England/CAMB-7C6FE/2020 | EPI_ISL_442300 | 04/04/2020 |
| CAMB-7C737 | hCoV-19/England/CAMB-7C737/2020 | EPI_ISL_442214 | 04/04/2020 |
| CAMB-7C791 | hCoV-19/England/CAMB-7C791/2020 | EPI_ISL_442282 | 04/04/2020 |
| CAMB-7C931 | hCoV-19/England/CAMB-7C931/2020 | EPI_ISL_442050 | 04/04/2020 |
| CAMB-7D39C | hCoV-19/England/CAMB-7D39C/2020 | EPI_ISL_442253 | 04/04/2020 |
| CAMB-80CB9 | hCoV-19/England/CAMB-80CB9/2020 | EPI_ISL_442954 | 04/04/2020 |
| CAMB-781E2 | hCoV-19/England/CAMB-781E2/2020 | EPI_ISL_433705 | 05/04/2020 |
| CAMB-7830A | hCoV-19/England/CAMB-7830A/2020 | EPI_ISL_433716 | 05/04/2020 |
| CAMB-78391 | hCoV-19/England/CAMB-78391/2020 | EPI_ISL_433725 | 05/04/2020 |
| CAMB-78407 | hCoV-19/England/CAMB-78407/2020 | EPI_ISL_433731 | 05/04/2020 |
| CAMB-785AA | hCoV-19/England/CAMB-785AA/2020 | EPI_ISL_433752 | 05/04/2020 |
| CAMB-785C8 | hCoV-19/England/CAMB-785C8/2020 | EPI_ISL_433753 | 05/04/2020 |
| CAMB-78601 | hCoV-19/England/CAMB-78601/2020 | EPI_ISL_433756 | 05/04/2020 |
| CAMB-7862F | hCoV-19/England/CAMB-7862F/2020 | EPI_ISL_433757 | 05/04/2020 |
| CAMB-786A7 | hCoV-19/England/CAMB-786A7/2020 | EPI_ISL_433762 | 05/04/2020 |
| CAMB-78786 | hCoV-19/England/CAMB-78786/2020 | EPI_ISL_433770 | 05/04/2020 |
| CAMB-7B914 | hCoV-19/England/CAMB-7B914/2020 | EPI_ISL_442195 | 05/04/2020 |
| CAMB-7BB87 | hCoV-19/England/CAMB-7BB87/2020 | EPI_ISL_442818 | 05/04/2020 |
| CAMB-7BBA5 | hCoV-19/England/CAMB-7BBA5/2020 | EPI_ISL_442131 | 05/04/2020 |
| CAMB-7BBB4 | hCoV-19/England/CAMB-7BBB4/2020 | EPI_ISL_442075 | 05/04/2020 |
| CAMB-7BBC3 | hCoV-19/England/CAMB-7BBC3/2020 | EPI_ISL_442105 | 05/04/2020 |
| CAMB-7BBE1 | hCoV-19/England/CAMB-7BBE1/2020 | EPI_ISL_442112 | 05/04/2020 |
| CAMB-7BCDF | hCoV-19/England/CAMB-7BCDF/2020 | EPI_ISL_442139 | 05/04/2020 |
| CAMB-7BD54 | hCoV-19/England/CAMB-7BD54/2020 | EPI_ISL_442122 | 05/04/2020 |
| CAMB-7BDEB | hCoV-19/England/CAMB-7BDEB/2020 | EPI_ISL_442134 | 05/04/2020 |
| CAMB-7BE42 | hCoV-19/England/CAMB-7BE42/2020 | EPI_ISL_442109 | 05/04/2020 |
| CAMB-7BEAC | hCoV-19/England/CAMB-7BEAC/2020 | EPI_ISL_470471 | 05/04/2020 |
| CAMB-7BF6D | hCoV-19/England/CAMB-7BF6D/2020 | EPI_ISL_470415 | 05/04/2020 |
| CAMB-7C403 | hCoV-19/England/CAMB-7C403/2020 | EPI_ISL_442325 | 05/04/2020 |
| CAMB-7C46D | hCoV-19/England/CAMB-7C46D/2020 | EPI_ISL_442310 | 05/04/2020 |
| CAMB-7C4D6 | hCoV-19/England/CAMB-7C4D6/2020 | EPI_ISL_442280 | 05/04/2020 |
| CAMB-7C61C | hCoV-19/England/CAMB-7C61C/2020 | EPI_ISL_442288 | 05/04/2020 |
| CAMB-7C649 | hCoV-19/England/CAMB-7C649/2020 | EPI_ISL_441621 | 05/04/2020 |
| CAMB-7C6EF | hCoV-19/England/CAMB-7C6EF/2020 | EPI_ISL_441577 | 05/04/2020 |
| CAMB-7C782 | hCoV-19/England/CAMB-7C782/2020 | EPI_ISL_442103 | 05/04/2020 |
| CAMB-7C7A0 | hCoV-19/England/CAMB-7C7A0/2020 | EPI_ISL_442048 | 05/04/2020 |
| CAMB-7C8DA | hCoV-19/England/CAMB-7C8DA/2020 | EPI_ISL_442124 | 05/04/2020 |
| CAMB-7C922 | hCoV-19/England/CAMB-7C922/2020 | EPI_ISL_442072 | 05/04/2020 |
| CAMB-7CA01 | hCoV-19/England/CAMB-7CA01/2020 | EPI_ISL_442813 | 05/04/2020 |
| CAMB-7CAF2 | hCoV-19/England/CAMB-7CAF2/2020 | EPI_ISL_442088 | 05/04/2020 |
| CAMB-7CB0E | hCoV-19/England/CAMB-7CB0E/2020 | EPI_ISL_442178 | 05/04/2020 |
| CAMB-7CBD1 | hCoV-19/England/CAMB-7CBD1/2020 | EPI_ISL_442047 | 05/04/2020 |
| CAMB-7CC29 | hCoV-19/England/CAMB-7CC29/2020 | EPI_ISL_442313 | 05/04/2020 |
| CAMB-7CC83 | hCoV-19/England/CAMB-7CC83/2020 | EPI_ISL_441558 | 05/04/2020 |
| CAMB-7CD17 | hCoV-19/England/CAMB-7CD17/2020 | EPI_ISL_441578 | 05/04/2020 |
| CAMB-7CD26 | hCoV-19/England/CAMB-7CD26/2020 | EPI_ISL_442318 | 05/04/2020 |
| CAMB-7CDAE | hCoV-19/England/CAMB-7CDAE/2020 | EPI_ISL_441644 | 05/04/2020 |
| CAMB-7CDF9 | hCoV-19/England/CAMB-7CDF9/2020 | EPI_ISL_442078 | 05/04/2020 |
| CAMB-7CE23 | hCoV-19/England/CAMB-7CE23/2020 | EPI_ISL_442164 | 05/04/2020 |
| CAMB-7D059 | hCoV-19/England/CAMB-7D059/2020 | EPI_ISL_442059 | 05/04/2020 |
| CAMB-7D077 | hCoV-19/England/CAMB-7D077/2020 | EPI_ISL_442045 | 05/04/2020 |
| CAMB-7D2F9 | hCoV-19/England/CAMB-7D2F9/2020 | EPI_ISL_442263 | 05/04/2020 |
| CAMB-7D305 | hCoV-19/England/CAMB-7D305/2020 | EPI_ISL_441560 | 05/04/2020 |
| CAMB-7D350 | hCoV-19/England/CAMB-7D350/2020 | EPI_ISL_442260 | 05/04/2020 |
| CAMB-7D420 | hCoV-19/England/CAMB-7D420/2020 | EPI_ISL_442276 | 05/04/2020 |
| CAMB-7D9C7 | hCoV-19/England/CAMB-7D9C7/2020 | EPI_ISL_441735 | 05/04/2020 |
| CAMB-7DBC1 | hCoV-19/England/CAMB-7DBC1/2020 | EPI_ISL_441561 | 05/04/2020 |
| CAMB-78276 | hCoV-19/England/CAMB-78276/2020 | EPI_ISL_433708 | 06/04/2020 |
| CAMB-7849E | hCoV-19/England/CAMB-7849E/2020 | EPI_ISL_433738 | 06/04/2020 |
| CAMB-784BC | hCoV-19/England/CAMB-784BC/2020 | EPI_ISL_433740 | 06/04/2020 |
| CAMB-784DA | hCoV-19/England/CAMB-784DA/2020 | EPI_ISL_433742 | 06/04/2020 |
| CAMB-7859B | hCoV-19/England/CAMB-7859B/2020 | EPI_ISL_433751 | 06/04/2020 |
| CAMB-7874A | hCoV-19/England/CAMB-7874A/2020 | EPI_ISL_433768 | 06/04/2020 |
| CAMB-78847 | hCoV-19/England/CAMB-78847/2020 | EPI_ISL_433780 | 06/04/2020 |
| CAMB-78865 | hCoV-19/England/CAMB-78865/2020 | EPI_ISL_433781 | 06/04/2020 |
| CAMB-78883 | hCoV-19/England/CAMB-78883/2020 | EPI_ISL_433783 | 06/04/2020 |
| CAMB-78944 | hCoV-19/England/CAMB-78944/2020 | EPI_ISL_433794 | 06/04/2020 |
| CAMB-789EA | hCoV-19/England/CAMB-789EA/2020 | EPI_ISL_433804 | 06/04/2020 |
| CAMB-78A32 | hCoV-19/England/CAMB-78A32/2020 | EPI_ISL_433808 | 06/04/2020 |
| CAMB-7ABE2 | hCoV-19/England/CAMB-7ABE2/2020 | EPI_ISL_433850 | 06/04/2020 |
| CAMB-7ACA3 | hCoV-19/England/CAMB-7ACA3/2020 | EPI_ISL_433862 | 06/04/2020 |
| CAMB-7C53D | hCoV-19/England/CAMB-7C53D/2020 | EPI_ISL_441641 | 06/04/2020 |
| CAMB-7C588 | hCoV-19/England/CAMB-7C588/2020 | EPI_ISL_441579 | 06/04/2020 |
| CAMB-7C5C4 | hCoV-19/England/CAMB-7C5C4/2020 | EPI_ISL_442212 | 06/04/2020 |
| CAMB-7C685 | hCoV-19/England/CAMB-7C685/2020 | EPI_ISL_441547 | 06/04/2020 |
| CAMB-7C746 | hCoV-19/England/CAMB-7C746/2020 | EPI_ISL_442177 | 06/04/2020 |
| CAMB-7C7CE | hCoV-19/England/CAMB-7C7CE/2020 | EPI_ISL_442055 | 06/04/2020 |
| CAMB-7C861 | hCoV-19/England/CAMB-7C861/2020 | EPI_ISL_441594 | 06/04/2020 |
| CAMB-7C8F8 | hCoV-19/England/CAMB-7C8F8/2020 | EPI_ISL_442184 | 06/04/2020 |
| CAMB-7CB3B | hCoV-19/England/CAMB-7CB3B/2020 | EPI_ISL_442082 | 06/04/2020 |
| CAMB-7CCA1 | hCoV-19/England/CAMB-7CCA1/2020 | EPI_ISL_441548 | 06/04/2020 |
| CAMB-7CCED | hCoV-19/England/CAMB-7CCED/2020 | EPI_ISL_441611 | 06/04/2020 |
| CAMB-7CD35 | hCoV-19/England/CAMB-7CD35/2020 | EPI_ISL_441638 | 06/04/2020 |
| CAMB-7CD80 | hCoV-19/England/CAMB-7CD80/2020 | EPI_ISL_442255 | 06/04/2020 |
| CAMB-7CE41 | hCoV-19/England/CAMB-7CE41/2020 | EPI_ISL_441567 | 06/04/2020 |
| CAMB-7CEAB | hCoV-19/England/CAMB-7CEAB/2020 | EPI_ISL_441603 | 06/04/2020 |
| CAMB-7CEC9 | hCoV-19/England/CAMB-7CEC9/2020 | EPI_ISL_442230 | 06/04/2020 |
| CAMB-7CF3F | hCoV-19/England/CAMB-7CF3F/2020 | EPI_ISL_442306 | 06/04/2020 |
| CAMB-7CF99 | hCoV-19/England/CAMB-7CF99/2020 | EPI_ISL_442272 | 06/04/2020 |
| CAMB-7CFA8 | hCoV-19/England/CAMB-7CFA8/2020 | EPI_ISL_442264 | 06/04/2020 |
| CAMB-7CFF3 | hCoV-19/England/CAMB-7CFF3/2020 | EPI_ISL_442168 | 06/04/2020 |
| CAMB-7D00E | hCoV-19/England/CAMB-7D00E/2020 | EPI_ISL_442183 | 06/04/2020 |
| CAMB-7D0FF | hCoV-19/England/CAMB-7D0FF/2020 | EPI_ISL_441605 | 06/04/2020 |
| CAMB-7D1A1 | hCoV-19/England/CAMB-7D1A1/2020 | EPI_ISL_442316 | 06/04/2020 |
| CAMB-7D1CF | hCoV-19/England/CAMB-7D1CF/2020 | EPI_ISL_442217 | 06/04/2020 |
| CAMB-7D226 | hCoV-19/England/CAMB-7D226/2020 | EPI_ISL_442090 | 06/04/2020 |
| CAMB-7D323 | hCoV-19/England/CAMB-7D323/2020 | EPI_ISL_441649 | 06/04/2020 |
| CAMB-7D37E | hCoV-19/England/CAMB-7D37E/2020 | EPI_ISL_442243 | 06/04/2020 |
| CAMB-7D3F6 | hCoV-19/England/CAMB-7D3F6/2020 | EPI_ISL_441632 | 06/04/2020 |
| CAMB-7D675 | hCoV-19/England/CAMB-7D675/2020 | EPI_ISL_441720 | 06/04/2020 |
| CAMB-7D684 | hCoV-19/England/CAMB-7D684/2020 | EPI_ISL_441563 | 06/04/2020 |
| CAMB-7D718 | hCoV-19/England/CAMB-7D718/2020 | EPI_ISL_441689 | 06/04/2020 |
| CAMB-7D754 | hCoV-19/England/CAMB-7D754/2020 | EPI_ISL_442326 | 06/04/2020 |
| CAMB-7D763 | hCoV-19/England/CAMB-7D763/2020 | EPI_ISL_441759 | 06/04/2020 |
| CAMB-7D790 | hCoV-19/England/CAMB-7D790/2020 | EPI_ISL_442340 | 06/04/2020 |
| CAMB-7D824 | hCoV-19/England/CAMB-7D824/2020 | EPI_ISL_441684 | 06/04/2020 |
| CAMB-7D851 | hCoV-19/England/CAMB-7D851/2020 | EPI_ISL_441580 | 06/04/2020 |
| CAMB-7D860 | hCoV-19/England/CAMB-7D860/2020 | EPI_ISL_441665 | 06/04/2020 |
| CAMB-7D89D | hCoV-19/England/CAMB-7D89D/2020 | EPI_ISL_442307 | 06/04/2020 |
| CAMB-7D97C | hCoV-19/England/CAMB-7D97C/2020 | EPI_ISL_441773 | 06/04/2020 |
| CAMB-7DA00 | hCoV-19/England/CAMB-7DA00/2020 | EPI_ISL_441778 | 06/04/2020 |
| CAMB-7DA3D | hCoV-19/England/CAMB-7DA3D/2020 | EPI_ISL_441666 | 06/04/2020 |
| CAMB-7DA6A | hCoV-19/England/CAMB-7DA6A/2020 | EPI_ISL_441553 | 06/04/2020 |
| CAMB-7DA97 | hCoV-19/England/CAMB-7DA97/2020 | EPI_ISL_441677 | 06/04/2020 |
| CAMB-7DAD3 | hCoV-19/England/CAMB-7DAD3/2020 | EPI_ISL_441722 | 06/04/2020 |
| CAMB-7DB76 | hCoV-19/England/CAMB-7DB76/2020 | EPI_ISL_441703 | 06/04/2020 |
| CAMB-7DC73 | hCoV-19/England/CAMB-7DC73/2020 | EPI_ISL_441757 | 06/04/2020 |
| CAMB-7DCA0 | hCoV-19/England/CAMB-7DCA0/2020 | EPI_ISL_442258 | 06/04/2020 |
| CAMB-7DD61 | hCoV-19/England/CAMB-7DD61/2020 | EPI_ISL_442110 | 06/04/2020 |
| CAMB-7DE04 | hCoV-19/England/CAMB-7DE04/2020 | EPI_ISL_442092 | 06/04/2020 |
| CAMB-7DE9B | hCoV-19/England/CAMB-7DE9B/2020 | EPI_ISL_470513 | 06/04/2020 |
| CAMB-7E049 | hCoV-19/England/CAMB-7E049/2020 | EPI_ISL_442093 | 06/04/2020 |
| CAMB-7ECDC | hCoV-19/England/CAMB-7ECDC/2020 | EPI_ISL_442881 | 06/04/2020 |
| CAMB-7ED60 | hCoV-19/England/CAMB-7ED60/2020 | EPI_ISL_442893 | 06/04/2020 |
| CAMB-7EFF1 | hCoV-19/England/CAMB-7EFF1/2020 | EPI_ISL_443032 | 06/04/2020 |
| CAMB-787D1 | hCoV-19/England/CAMB-787D1/2020 | EPI_ISL_433774 | 07/04/2020 |
| CAMB-788ED | hCoV-19/England/CAMB-788ED/2020 | EPI_ISL_433789 | 07/04/2020 |
| CAMB-78953 | hCoV-19/England/CAMB-78953/2020 | EPI_ISL_433795 | 07/04/2020 |
| CAMB-78971 | hCoV-19/England/CAMB-78971/2020 | EPI_ISL_433797 | 07/04/2020 |
| CAMB-789DB | hCoV-19/England/CAMB-789DB/2020 | EPI_ISL_433803 | 07/04/2020 |
| CAMB-78A14 | hCoV-19/England/CAMB-78A14/2020 | EPI_ISL_433807 | 07/04/2020 |
| CAMB-78A50 | hCoV-19/England/CAMB-78A50/2020 | EPI_ISL_433809 | 07/04/2020 |
| CAMB-7A8FA | hCoV-19/England/CAMB-7A8FA/2020 | EPI_ISL_433828 | 07/04/2020 |
| CAMB-7AA7C | hCoV-19/England/CAMB-7AA7C/2020 | EPI_ISL_433832 | 07/04/2020 |
| CAMB-7AAC7 | hCoV-19/England/CAMB-7AAC7/2020 | EPI_ISL_433836 | 07/04/2020 |
| CAMB-7AAF4 | hCoV-19/England/CAMB-7AAF4/2020 | EPI_ISL_433839 | 07/04/2020 |
| CAMB-7AB00 | hCoV-19/England/CAMB-7AB00/2020 | EPI_ISL_433840 | 07/04/2020 |
| CAMB-7AB79 | hCoV-19/England/CAMB-7AB79/2020 | EPI_ISL_433844 | 07/04/2020 |
| CAMB-7ABA6 | hCoV-19/England/CAMB-7ABA6/2020 | EPI_ISL_433847 | 07/04/2020 |
| CAMB-7AD19 | hCoV-19/England/CAMB-7AD19/2020 | EPI_ISL_444394 | 07/04/2020 |
| CAMB-7AD46 | hCoV-19/England/CAMB-7AD46/2020 | EPI_ISL_433870 | 07/04/2020 |
| CAMB-7AD55 | hCoV-19/England/CAMB-7AD55/2020 | EPI_ISL_433871 | 07/04/2020 |
| CAMB-7D4A8 | hCoV-19/England/CAMB-7D4A8/2020 | EPI_ISL_441705 | 07/04/2020 |
| CAMB-7D53C | hCoV-19/England/CAMB-7D53C/2020 | EPI_ISL_442311 | 07/04/2020 |
| CAMB-7D54B | hCoV-19/England/CAMB-7D54B/2020 | EPI_ISL_441728 | 07/04/2020 |
| CAMB-7D7BE | hCoV-19/England/CAMB-7D7BE/2020 | EPI_ISL_442216 | 07/04/2020 |
| CAMB-7D833 | hCoV-19/England/CAMB-7D833/2020 | EPI_ISL_441566 | 07/04/2020 |
| CAMB-7DA4C | hCoV-19/England/CAMB-7DA4C/2020 | EPI_ISL_441582 | 07/04/2020 |
| CAMB-7DB49 | hCoV-19/England/CAMB-7DB49/2020 | EPI_ISL_441734 | 07/04/2020 |
| CAMB-7DBEF | hCoV-19/England/CAMB-7DBEF/2020 | EPI_ISL_442221 | 07/04/2020 |
| CAMB-7DCEC | hCoV-19/England/CAMB-7DCEC/2020 | EPI_ISL_442273 | 07/04/2020 |
| CAMB-7DF01 | hCoV-19/England/CAMB-7DF01/2020 | EPI_ISL_442236 | 07/04/2020 |
| CAMB-7DF89 | hCoV-19/England/CAMB-7DF89/2020 | EPI_ISL_441633 | 07/04/2020 |
| CAMB-7DFA7 | hCoV-19/England/CAMB-7DFA7/2020 | EPI_ISL_442294 | 07/04/2020 |
| CAMB-7DFF2 | hCoV-19/England/CAMB-7DFF2/2020 | EPI_ISL_442200 | 07/04/2020 |
| CAMB-7E0C1 | hCoV-19/England/CAMB-7E0C1/2020 | EPI_ISL_442179 | 07/04/2020 |
| CAMB-7E0FE | hCoV-19/England/CAMB-7E0FE/2020 | EPI_ISL_442056 | 07/04/2020 |
| CAMB-7E841 | hCoV-19/England/CAMB-7E841/2020 | EPI_ISL_443081 | 07/04/2020 |
| CAMB-7E96C | hCoV-19/England/CAMB-7E96C/2020 | EPI_ISL_443064 | 07/04/2020 |
| CAMB-7E98A | hCoV-19/England/CAMB-7E98A/2020 | EPI_ISL_442560 | 07/04/2020 |
| CAMB-7E9E4 | hCoV-19/England/CAMB-7E9E4/2020 | EPI_ISL_443053 | 07/04/2020 |
| CAMB-7EA4B | hCoV-19/England/CAMB-7EA4B/2020 | EPI_ISL_442553 | 07/04/2020 |
| CAMB-7EA87 | hCoV-19/England/CAMB-7EA87/2020 | EPI_ISL_442582 | 07/04/2020 |
| CAMB-7EAE1 | hCoV-19/England/CAMB-7EAE1/2020 | EPI_ISL_442849 | 07/04/2020 |
| CAMB-7EAF0 | hCoV-19/England/CAMB-7EAF0/2020 | EPI_ISL_442891 | 07/04/2020 |
| CAMB-7EB39 | hCoV-19/England/CAMB-7EB39/2020 | EPI_ISL_442917 | 07/04/2020 |
| CAMB-7EB66 | hCoV-19/England/CAMB-7EB66/2020 | EPI_ISL_442932 | 07/04/2020 |
| CAMB-7EB75 | hCoV-19/England/CAMB-7EB75/2020 | EPI_ISL_442963 | 07/04/2020 |
| CAMB-7EBC0 | hCoV-19/England/CAMB-7EBC0/2020 | EPI_ISL_443008 | 07/04/2020 |
| CAMB-7EC18 | hCoV-19/England/CAMB-7EC18/2020 | EPI_ISL_442601 | 07/04/2020 |
| CAMB-7EC90 | hCoV-19/England/CAMB-7EC90/2020 | EPI_ISL_442888 | 07/04/2020 |
| CAMB-7EE30 | hCoV-19/England/CAMB-7EE30/2020 | EPI_ISL_442871 | 07/04/2020 |
| CAMB-7EE7C | hCoV-19/England/CAMB-7EE7C/2020 | EPI_ISL_442897 | 07/04/2020 |
| CAMB-7EEC7 | hCoV-19/England/CAMB-7EEC7/2020 | EPI_ISL_442850 | 07/04/2020 |
| CAMB-7F00C | hCoV-19/England/CAMB-7F00C/2020 | EPI_ISL_443020 | 07/04/2020 |
| CAMB-7F048 | hCoV-19/England/CAMB-7F048/2020 | EPI_ISL_591080 | 07/04/2020 |
| CAMB-7F057 | hCoV-19/England/CAMB-7F057/2020 | EPI_ISL_442978 | 07/04/2020 |
| CAMB-7F066 | hCoV-19/England/CAMB-7F066/2020 | EPI_ISL_442840 | 07/04/2020 |
| CAMB-7F0C0 | hCoV-19/England/CAMB-7F0C0/2020 | EPI_ISL_442901 | 07/04/2020 |
| CAMB-7F233 | hCoV-19/England/CAMB-7F233/2020 | EPI_ISL_442884 | 07/04/2020 |
| CAMB-7F2BB | hCoV-19/England/CAMB-7F2BB/2020 | EPI_ISL_442900 | 07/04/2020 |
| CAMB-80A37 | hCoV-19/England/CAMB-80A37/2020 | EPI_ISL_442941 | 07/04/2020 |
| CAMB-80ABF | hCoV-19/England/CAMB-80ABF/2020 | EPI_ISL_470508 | 07/04/2020 |
| CAMB-78838 | hCoV-19/England/CAMB-78838/2020 | EPI_ISL_433779 | 08/04/2020 |
| CAMB-78917 | hCoV-19/England/CAMB-78917/2020 | EPI_ISL_433791 | 08/04/2020 |
| CAMB-7AB1F | hCoV-19/England/CAMB-7AB1F/2020 | EPI_ISL_433841 | 08/04/2020 |
| CAMB-7AC0D | hCoV-19/England/CAMB-7AC0D/2020 | EPI_ISL_433852 | 08/04/2020 |
| CAMB-7AC3A | hCoV-19/England/CAMB-7AC3A/2020 | EPI_ISL_433855 | 08/04/2020 |
| CAMB-7AC49 | hCoV-19/England/CAMB-7AC49/2020 | EPI_ISL_433856 | 08/04/2020 |
| CAMB-7AC76 | hCoV-19/England/CAMB-7AC76/2020 | EPI_ISL_433859 | 08/04/2020 |
| CAMB-7AD28 | hCoV-19/England/CAMB-7AD28/2020 | EPI_ISL_433868 | 08/04/2020 |
| CAMB-7AD37 | hCoV-19/England/CAMB-7AD37/2020 | EPI_ISL_433869 | 08/04/2020 |
| CAMB-7AD64 | hCoV-19/England/CAMB-7AD64/2020 | EPI_ISL_433872 | 08/04/2020 |
| CAMB-7AD91 | hCoV-19/England/CAMB-7AD91/2020 | EPI_ISL_433874 | 08/04/2020 |
| CAMB-7ADA0 | hCoV-19/England/CAMB-7ADA0/2020 | EPI_ISL_433875 | 08/04/2020 |
| CAMB-7ADBF | hCoV-19/England/CAMB-7ADBF/2020 | EPI_ISL_433876 | 08/04/2020 |
| CAMB-7E86F | hCoV-19/England/CAMB-7E86F/2020 | EPI_ISL_443096 | 08/04/2020 |
| CAMB-7E8E7 | hCoV-19/England/CAMB-7E8E7/2020 | EPI_ISL_442546 | 08/04/2020 |
| CAMB-7EE5E | hCoV-19/England/CAMB-7EE5E/2020 | EPI_ISL_443002 | 08/04/2020 |
| CAMB-7EF88 | hCoV-19/England/CAMB-7EF88/2020 | EPI_ISL_442929 | 08/04/2020 |
| CAMB-7EFD3 | hCoV-19/England/CAMB-7EFD3/2020 | EPI_ISL_442974 | 08/04/2020 |
| CAMB-7F01B | hCoV-19/England/CAMB-7F01B/2020 | EPI_ISL_442916 | 08/04/2020 |
| CAMB-7F0B1 | hCoV-19/England/CAMB-7F0B1/2020 | EPI_ISL_442836 | 08/04/2020 |
| CAMB-7F1FA | hCoV-19/England/CAMB-7F1FA/2020 | EPI_ISL_442896 | 08/04/2020 |
| CAMB-7F27F | hCoV-19/England/CAMB-7F27F/2020 | EPI_ISL_443018 | 08/04/2020 |
| CAMB-7F28E | hCoV-19/England/CAMB-7F28E/2020 | EPI_ISL_442868 | 08/04/2020 |
| CAMB-7F29D | hCoV-19/England/CAMB-7F29D/2020 | EPI_ISL_442926 | 08/04/2020 |
| CAMB-7F2E8 | hCoV-19/England/CAMB-7F2E8/2020 | EPI_ISL_442967 | 08/04/2020 |
| CAMB-7F312 | hCoV-19/England/CAMB-7F312/2020 | EPI_ISL_442965 | 08/04/2020 |
| CAMB-7F34F | hCoV-19/England/CAMB-7F34F/2020 | EPI_ISL_442977 | 08/04/2020 |
| CAMB-7F37C | hCoV-19/England/CAMB-7F37C/2020 | EPI_ISL_442843 | 08/04/2020 |
| CAMB-7F400 | hCoV-19/England/CAMB-7F400/2020 | EPI_ISL_443039 | 08/04/2020 |
| CAMB-7F46A | hCoV-19/England/CAMB-7F46A/2020 | EPI_ISL_442990 | 08/04/2020 |
| CAMB-7F4A6 | hCoV-19/England/CAMB-7F4A6/2020 | EPI_ISL_443047 | 08/04/2020 |
| CAMB-7F4F1 | hCoV-19/England/CAMB-7F4F1/2020 | EPI_ISL_442852 | 08/04/2020 |
| CAMB-7F558 | hCoV-19/England/CAMB-7F558/2020 | EPI_ISL_442910 | 08/04/2020 |
| CAMB-7F576 | hCoV-19/England/CAMB-7F576/2020 | EPI_ISL_442946 | 08/04/2020 |
| CAMB-7F619 | hCoV-19/England/CAMB-7F619/2020 | EPI_ISL_442950 | 08/04/2020 |
| CAMB-7F79E | hCoV-19/England/CAMB-7F79E/2020 | EPI_ISL_443019 | 08/04/2020 |
| CAMB-7FA59 | hCoV-19/England/CAMB-7FA59/2020 | EPI_ISL_470486 | 08/04/2020 |
| CAMB-7ABF1 | hCoV-19/England/CAMB-7ABF1/2020 | EPI_ISL_433851 | 09/04/2020 |
| CAMB-7AE07 | hCoV-19/England/CAMB-7AE07/2020 | EPI_ISL_433880 | 09/04/2020 |
| CAMB-7AE52 | hCoV-19/England/CAMB-7AE52/2020 | EPI_ISL_433884 | 09/04/2020 |
| CAMB-7AE61 | hCoV-19/England/CAMB-7AE61/2020 | EPI_ISL_433885 | 09/04/2020 |
| CAMB-7AEAD | hCoV-19/England/CAMB-7AEAD/2020 | EPI_ISL_433888 | 09/04/2020 |
| CAMB-7AEDA | hCoV-19/England/CAMB-7AEDA/2020 | EPI_ISL_433890 | 09/04/2020 |
| CAMB-7AF8C | hCoV-19/England/CAMB-7AF8C/2020 | EPI_ISL_433901 | 09/04/2020 |
| CAMB-7AFAA | hCoV-19/England/CAMB-7AFAA/2020 | EPI_ISL_433902 | 09/04/2020 |
| CAMB-7B2FB | hCoV-19/England/CAMB-7B2FB/2020 | EPI_ISL_444396 | 09/04/2020 |
| CAMB-7F716 | hCoV-19/England/CAMB-7F716/2020 | EPI_ISL_470498 | 09/04/2020 |
| CAMB-7F85F | hCoV-19/England/CAMB-7F85F/2020 | EPI_ISL_470478 | 09/04/2020 |
| CAMB-7F95C | hCoV-19/England/CAMB-7F95C/2020 | EPI_ISL_470445 | 09/04/2020 |
| CAMB-7FA68 | hCoV-19/England/CAMB-7FA68/2020 | EPI_ISL_470476 | 09/04/2020 |
| CAMB-80123 | hCoV-19/England/CAMB-80123/2020 | EPI_ISL_470512 | 09/04/2020 |
| CAMB-8018D | hCoV-19/England/CAMB-8018D/2020 | EPI_ISL_470373 | 09/04/2020 |
| CAMB-80466 | hCoV-19/England/CAMB-80466/2020 | EPI_ISL_470434 | 09/04/2020 |
| CAMB-80527 | hCoV-19/England/CAMB-80527/2020 | EPI_ISL_470474 | 09/04/2020 |
| CAMB-80590 | hCoV-19/England/CAMB-80590/2020 | EPI_ISL_470377 | 09/04/2020 |
| CAMB-805EB | hCoV-19/England/CAMB-805EB/2020 | EPI_ISL_470438 | 09/04/2020 |
| CAMB-8077C | hCoV-19/England/CAMB-8077C/2020 | EPI_ISL_470455 | 09/04/2020 |
| CAMB-80897 | hCoV-19/England/CAMB-80897/2020 | EPI_ISL_442895 | 09/04/2020 |
| CAMB-80976 | hCoV-19/England/CAMB-80976/2020 | EPI_ISL_442867 | 09/04/2020 |
| CAMB-809D0 | hCoV-19/England/CAMB-809D0/2020 | EPI_ISL_442912 | 09/04/2020 |
| CAMB-80CF5 | hCoV-19/England/CAMB-80CF5/2020 | EPI_ISL_443005 | 09/04/2020 |
| CAMB-81438 | hCoV-19/England/CAMB-81438/2020 | EPI_ISL_442593 | 09/04/2020 |
| CAMB-81483 | hCoV-19/England/CAMB-81483/2020 | EPI_ISL_443113 | 09/04/2020 |
| CAMB-7AA6D | hCoV-19/England/CAMB-7AA6D/2020 | EPI_ISL_433831 | 10/04/2020 |
| CAMB-7AF40 | hCoV-19/England/CAMB-7AF40/2020 | EPI_ISL_433897 | 10/04/2020 |
| CAMB-7B0F1 | hCoV-19/England/CAMB-7B0F1/2020 | EPI_ISL_433918 | 10/04/2020 |
| CAMB-7B219 | hCoV-19/England/CAMB-7B219/2020 | EPI_ISL_444395 | 10/04/2020 |
| CAMB-7B2A0 | hCoV-19/England/CAMB-7B2A0/2020 | EPI_ISL_433936 | 10/04/2020 |
| CAMB-80518 | hCoV-19/England/CAMB-80518/2020 | EPI_ISL_470468 | 10/04/2020 |
| CAMB-80536 | hCoV-19/England/CAMB-80536/2020 | EPI_ISL_470457 | 10/04/2020 |
| CAMB-805DC | hCoV-19/England/CAMB-805DC/2020 | EPI_ISL_470430 | 10/04/2020 |
| CAMB-80642 | hCoV-19/England/CAMB-80642/2020 | EPI_ISL_470417 | 10/04/2020 |
| CAMB-806AC | hCoV-19/England/CAMB-806AC/2020 | EPI_ISL_442562 | 10/04/2020 |
| CAMB-80800 | hCoV-19/England/CAMB-80800/2020 | EPI_ISL_442966 | 10/04/2020 |
| CAMB-8082E | hCoV-19/England/CAMB-8082E/2020 | EPI_ISL_442902 | 10/04/2020 |
| CAMB-808F1 | hCoV-19/England/CAMB-808F1/2020 | EPI_ISL_442586 | 10/04/2020 |
| CAMB-8090D | hCoV-19/England/CAMB-8090D/2020 | EPI_ISL_443034 | 10/04/2020 |
| CAMB-80958 | hCoV-19/England/CAMB-80958/2020 | EPI_ISL_442622 | 10/04/2020 |
| CAMB-80994 | hCoV-19/England/CAMB-80994/2020 | EPI_ISL_442873 | 10/04/2020 |
| CAMB-80BAD | hCoV-19/England/CAMB-80BAD/2020 | EPI_ISL_443015 | 10/04/2020 |
| CAMB-80BDA | hCoV-19/England/CAMB-80BDA/2020 | EPI_ISL_442993 | 10/04/2020 |
| CAMB-80C40 | hCoV-19/England/CAMB-80C40/2020 | EPI_ISL_442915 | 10/04/2020 |
| CAMB-80CD7 | hCoV-19/England/CAMB-80CD7/2020 | EPI_ISL_442964 | 10/04/2020 |
| CAMB-80D01 | hCoV-19/England/CAMB-80D01/2020 | EPI_ISL_443013 | 10/04/2020 |
| CAMB-80D89 | hCoV-19/England/CAMB-80D89/2020 | EPI_ISL_442877 | 10/04/2020 |
| CAMB-80DA7 | hCoV-19/England/CAMB-80DA7/2020 | EPI_ISL_442832 | 10/04/2020 |
| CAMB-80DE3 | hCoV-19/England/CAMB-80DE3/2020 | EPI_ISL_442953 | 10/04/2020 |
| CAMB-80DF2 | hCoV-19/England/CAMB-80DF2/2020 | EPI_ISL_442991 | 10/04/2020 |
| CAMB-80E0E | hCoV-19/England/CAMB-80E0E/2020 | EPI_ISL_442979 | 10/04/2020 |
| CAMB-80E2C | hCoV-19/England/CAMB-80E2C/2020 | EPI_ISL_442844 | 10/04/2020 |
| CAMB-8124D | hCoV-19/England/CAMB-8124D/2020 | EPI_ISL_443038 | 10/04/2020 |
| CAMB-81298 | hCoV-19/England/CAMB-81298/2020 | EPI_ISL_443027 | 10/04/2020 |
| CAMB-8130E | hCoV-19/England/CAMB-8130E/2020 | EPI_ISL_443029 | 10/04/2020 |
| CAMB-8132C | hCoV-19/England/CAMB-8132C/2020 | EPI_ISL_442924 | 10/04/2020 |
| CAMB-81368 | hCoV-19/England/CAMB-81368/2020 | EPI_ISL_442857 | 10/04/2020 |
| CAMB-813C2 | hCoV-19/England/CAMB-813C2/2020 | EPI_ISL_442594 | 10/04/2020 |
| CAMB-813FF | hCoV-19/England/CAMB-813FF/2020 | EPI_ISL_443088 | 10/04/2020 |
| CAMB-81474 | hCoV-19/England/CAMB-81474/2020 | EPI_ISL_442573 | 10/04/2020 |
| CAMB-81492 | hCoV-19/England/CAMB-81492/2020 | EPI_ISL_443086 | 10/04/2020 |
| CAMB-814B0 | hCoV-19/England/CAMB-814B0/2020 | EPI_ISL_442527 | 10/04/2020 |
| CAMB-815F9 | hCoV-19/England/CAMB-815F9/2020 | EPI_ISL_442853 | 10/04/2020 |
| CAMB-82FAF | hCoV-19/England/CAMB-82FAF/2020 | EPI_ISL_443373 | 10/04/2020 |
| CAMB-7B06A | hCoV-19/England/CAMB-7B06A/2020 | EPI_ISL_433909 | 11/04/2020 |
| CAMB-7B079 | hCoV-19/England/CAMB-7B079/2020 | EPI_ISL_433910 | 11/04/2020 |
| CAMB-7B0C4 | hCoV-19/England/CAMB-7B0C4/2020 | EPI_ISL_433915 | 11/04/2020 |
| CAMB-7B0D3 | hCoV-19/England/CAMB-7B0D3/2020 | EPI_ISL_433916 | 11/04/2020 |
| CAMB-7B10D | hCoV-19/England/CAMB-7B10D/2020 | EPI_ISL_433919 | 11/04/2020 |
| CAMB-7B1B2 | hCoV-19/England/CAMB-7B1B2/2020 | EPI_ISL_433928 | 11/04/2020 |
| CAMB-7B2EC | hCoV-19/England/CAMB-7B2EC/2020 | EPI_ISL_433939 | 11/04/2020 |
| CAMB-7B316 | hCoV-19/England/CAMB-7B316/2020 | EPI_ISL_433941 | 11/04/2020 |
| CAMB-80B16 | hCoV-19/England/CAMB-80B16/2020 | EPI_ISL_442848 | 11/04/2020 |
| CAMB-80B34 | hCoV-19/England/CAMB-80B34/2020 | EPI_ISL_443006 | 11/04/2020 |
| CAMB-80B43 | hCoV-19/England/CAMB-80B43/2020 | EPI_ISL_442878 | 11/04/2020 |
| CAMB-80B52 | hCoV-19/England/CAMB-80B52/2020 | EPI_ISL_443046 | 11/04/2020 |
| CAMB-80B9E | hCoV-19/England/CAMB-80B9E/2020 | EPI_ISL_442880 | 11/04/2020 |
| CAMB-80DB6 | hCoV-19/England/CAMB-80DB6/2020 | EPI_ISL_442828 | 11/04/2020 |
| CAMB-811F5 | hCoV-19/England/CAMB-811F5/2020 | EPI_ISL_442907 | 11/04/2020 |
| CAMB-812B6 | hCoV-19/England/CAMB-812B6/2020 | EPI_ISL_443016 | 11/04/2020 |
| CAMB-812C5 | hCoV-19/England/CAMB-812C5/2020 | EPI_ISL_442862 | 11/04/2020 |
| CAMB-8131D | hCoV-19/England/CAMB-8131D/2020 | EPI_ISL_443009 | 11/04/2020 |
| CAMB-813A4 | hCoV-19/England/CAMB-813A4/2020 | EPI_ISL_443102 | 11/04/2020 |
| CAMB-813E0 | hCoV-19/England/CAMB-813E0/2020 | EPI_ISL_442580 | 11/04/2020 |
| CAMB-814CF | hCoV-19/England/CAMB-814CF/2020 | EPI_ISL_442581 | 11/04/2020 |
| CAMB-815CC | hCoV-19/England/CAMB-815CC/2020 | EPI_ISL_443033 | 11/04/2020 |
| CAMB-81623 | hCoV-19/England/CAMB-81623/2020 | EPI_ISL_442988 | 11/04/2020 |
| CAMB-816E7 | hCoV-19/England/CAMB-816E7/2020 | EPI_ISL_443110 | 11/04/2020 |
| CAMB-816F6 | hCoV-19/England/CAMB-816F6/2020 | EPI_ISL_442563 | 11/04/2020 |
| CAMB-81711 | hCoV-19/England/CAMB-81711/2020 | EPI_ISL_443073 | 11/04/2020 |
| CAMB-8173F | hCoV-19/England/CAMB-8173F/2020 | EPI_ISL_442613 | 11/04/2020 |
| CAMB-8175D | hCoV-19/England/CAMB-8175D/2020 | EPI_ISL_443099 | 11/04/2020 |
| CAMB-81799 | hCoV-19/England/CAMB-81799/2020 | EPI_ISL_442564 | 11/04/2020 |
| CAMB-817A8 | hCoV-19/England/CAMB-817A8/2020 | EPI_ISL_442596 | 11/04/2020 |
| CAMB-817C6 | hCoV-19/England/CAMB-817C6/2020 | EPI_ISL_442535 | 11/04/2020 |
| CAMB-817E4 | hCoV-19/England/CAMB-817E4/2020 | EPI_ISL_443112 | 11/04/2020 |
| CAMB-8182D | hCoV-19/England/CAMB-8182D/2020 | EPI_ISL_443105 | 11/04/2020 |
| CAMB-819A2 | hCoV-19/England/CAMB-819A2/2020 | EPI_ISL_443079 | 11/04/2020 |
| CAMB-819EE | hCoV-19/England/CAMB-819EE/2020 | EPI_ISL_442532 | 11/04/2020 |
| CAMB-81A81 | hCoV-19/England/CAMB-81A81/2020 | EPI_ISL_443084 | 11/04/2020 |
| CAMB-81AAF | hCoV-19/England/CAMB-81AAF/2020 | EPI_ISL_443087 | 11/04/2020 |
| CAMB-81ACD | hCoV-19/England/CAMB-81ACD/2020 | EPI_ISL_443109 | 11/04/2020 |
| CAMB-81AEB | hCoV-19/England/CAMB-81AEB/2020 | EPI_ISL_443093 | 11/04/2020 |
| CAMB-81AFA | hCoV-19/England/CAMB-81AFA/2020 | EPI_ISL_442555 | 11/04/2020 |
| CAMB-81B9D | hCoV-19/England/CAMB-81B9D/2020 | EPI_ISL_442561 | 11/04/2020 |
| CAMB-83366 | hCoV-19/England/CAMB-83366/2020 | EPI_ISL_443440 | 11/04/2020 |
| CAMB-7B1EF | hCoV-19/England/CAMB-7B1EF/2020 | EPI_ISL_433931 | 12/04/2020 |
| CAMB-7B2BF | hCoV-19/England/CAMB-7B2BF/2020 | EPI_ISL_433937 | 12/04/2020 |
| CAMB-7B307 | hCoV-19/England/CAMB-7B307/2020 | EPI_ISL_433940 | 12/04/2020 |
| CAMB-7B334 | hCoV-19/England/CAMB-7B334/2020 | EPI_ISL_433943 | 12/04/2020 |
| CAMB-7B3DA | hCoV-19/England/CAMB-7B3DA/2020 | EPI_ISL_433951 | 12/04/2020 |
| CAMB-7B3E9 | hCoV-19/England/CAMB-7B3E9/2020 | EPI_ISL_433952 | 12/04/2020 |
| CAMB-7B4C8 | hCoV-19/England/CAMB-7B4C8/2020 | EPI_ISL_433965 | 12/04/2020 |
| CAMB-7C00F | hCoV-19/England/CAMB-7C00F/2020 | EPI_ISL_433979 | 12/04/2020 |
| CAMB-7C087 | hCoV-19/England/CAMB-7C087/2020 | EPI_ISL_433987 | 12/04/2020 |
| CAMB-81526 | hCoV-19/England/CAMB-81526/2020 | EPI_ISL_442952 | 12/04/2020 |
| CAMB-81580 | hCoV-19/England/CAMB-81580/2020 | EPI_ISL_442961 | 12/04/2020 |
| CAMB-8168D | hCoV-19/England/CAMB-8168D/2020 | EPI_ISL_443062 | 12/04/2020 |
| CAMB-8174E | hCoV-19/England/CAMB-8174E/2020 | EPI_ISL_442548 | 12/04/2020 |
| CAMB-8178A | hCoV-19/England/CAMB-8178A/2020 | EPI_ISL_443082 | 12/04/2020 |
| CAMB-8181E | hCoV-19/England/CAMB-8181E/2020 | EPI_ISL_443057 | 12/04/2020 |
| CAMB-8185A | hCoV-19/England/CAMB-8185A/2020 | EPI_ISL_443111 | 12/04/2020 |
| CAMB-81896 | hCoV-19/England/CAMB-81896/2020 | EPI_ISL_443094 | 12/04/2020 |
| CAMB-818A5 | hCoV-19/England/CAMB-818A5/2020 | EPI_ISL_443078 | 12/04/2020 |
| CAMB-818E1 | hCoV-19/England/CAMB-818E1/2020 | EPI_ISL_442529 | 12/04/2020 |
| CAMB-81966 | hCoV-19/England/CAMB-81966/2020 | EPI_ISL_443116 | 12/04/2020 |
| CAMB-81984 | hCoV-19/England/CAMB-81984/2020 | EPI_ISL_591092 | 12/04/2020 |
| CAMB-81BAC | hCoV-19/England/CAMB-81BAC/2020 | EPI_ISL_442591 | 12/04/2020 |
| CAMB-81C21 | hCoV-19/England/CAMB-81C21/2020 | EPI_ISL_442575 | 12/04/2020 |
| CAMB-81C30 | hCoV-19/England/CAMB-81C30/2020 | EPI_ISL_443060 | 12/04/2020 |
| CAMB-81C6D | hCoV-19/England/CAMB-81C6D/2020 | EPI_ISL_442551 | 12/04/2020 |
| CAMB-81C8B | hCoV-19/England/CAMB-81C8B/2020 | EPI_ISL_443100 | 12/04/2020 |
| CAMB-8225B | hCoV-19/England/CAMB-8225B/2020 | EPI_ISL_443428 | 12/04/2020 |
| CAMB-8233A | hCoV-19/England/CAMB-8233A/2020 | EPI_ISL_443461 | 12/04/2020 |
| CAMB-825F8 | hCoV-19/England/CAMB-825F8/2020 | EPI_ISL_443673 | 12/04/2020 |
| CAMB-8267D | hCoV-19/England/CAMB-8267D/2020 | EPI_ISL_443504 | 12/04/2020 |
| CAMB-82E93 | hCoV-19/England/CAMB-82E93/2020 | EPI_ISL_443356 | 12/04/2020 |
| CAMB-82F72 | hCoV-19/England/CAMB-82F72/2020 | EPI_ISL_443425 | 12/04/2020 |
| CAMB-7B413 | hCoV-19/England/CAMB-7B413/2020 | EPI_ISL_433955 | 13/04/2020 |
| CAMB-7B422 | hCoV-19/England/CAMB-7B422/2020 | EPI_ISL_433956 | 13/04/2020 |
| CAMB-7B440 | hCoV-19/England/CAMB-7B440/2020 | EPI_ISL_433958 | 13/04/2020 |
| CAMB-7B55C | hCoV-19/England/CAMB-7B55C/2020 | EPI_ISL_433974 | 13/04/2020 |
| CAMB-7BFD6 | hCoV-19/England/CAMB-7BFD6/2020 | EPI_ISL_433976 | 13/04/2020 |
| CAMB-7BFE5 | hCoV-19/England/CAMB-7BFE5/2020 | EPI_ISL_433977 | 13/04/2020 |
| CAMB-7C02D | hCoV-19/England/CAMB-7C02D/2020 | EPI_ISL_433981 | 13/04/2020 |
| CAMB-7C03C | hCoV-19/England/CAMB-7C03C/2020 | EPI_ISL_433982 | 13/04/2020 |
| CAMB-7C0A5 | hCoV-19/England/CAMB-7C0A5/2020 | EPI_ISL_433989 | 13/04/2020 |
| CAMB-7C10C | hCoV-19/England/CAMB-7C10C/2020 | EPI_ISL_433995 | 13/04/2020 |
| CAMB-7C11B | hCoV-19/England/CAMB-7C11B/2020 | EPI_ISL_433996 | 13/04/2020 |
| CAMB-7C166 | hCoV-19/England/CAMB-7C166/2020 | EPI_ISL_434001 | 13/04/2020 |
| CAMB-7C1A2 | hCoV-19/England/CAMB-7C1A2/2020 | EPI_ISL_434005 | 13/04/2020 |
| CAMB-81948 | hCoV-19/England/CAMB-81948/2020 | EPI_ISL_443101 | 13/04/2020 |
| CAMB-81975 | hCoV-19/England/CAMB-81975/2020 | EPI_ISL_443117 | 13/04/2020 |
| CAMB-81B42 | hCoV-19/England/CAMB-81B42/2020 | EPI_ISL_443055 | 13/04/2020 |
| CAMB-81B60 | hCoV-19/England/CAMB-81B60/2020 | EPI_ISL_442526 | 13/04/2020 |
| CAMB-81C03 | hCoV-19/England/CAMB-81C03/2020 | EPI_ISL_442602 | 13/04/2020 |
| CAMB-81D3D | hCoV-19/England/CAMB-81D3D/2020 | EPI_ISL_443106 | 13/04/2020 |
| CAMB-81D6A | hCoV-19/England/CAMB-81D6A/2020 | EPI_ISL_442567 | 13/04/2020 |
| CAMB-81DC4 | hCoV-19/England/CAMB-81DC4/2020 | EPI_ISL_443075 | 13/04/2020 |
| CAMB-8226A | hCoV-19/England/CAMB-8226A/2020 | EPI_ISL_443503 | 13/04/2020 |
| CAMB-823D0 | hCoV-19/England/CAMB-823D0/2020 | EPI_ISL_591079 | 13/04/2020 |
| CAMB-82419 | hCoV-19/England/CAMB-82419/2020 | EPI_ISL_443439 | 13/04/2020 |
| CAMB-82437 | hCoV-19/England/CAMB-82437/2020 | EPI_ISL_443533 | 13/04/2020 |
| CAMB-82455 | hCoV-19/England/CAMB-82455/2020 | EPI_ISL_591077 | 13/04/2020 |
| CAMB-824DD | hCoV-19/England/CAMB-824DD/2020 | EPI_ISL_443653 | 13/04/2020 |
| CAMB-82570 | hCoV-19/England/CAMB-82570/2020 | EPI_ISL_443562 | 13/04/2020 |
| CAMB-82640 | hCoV-19/England/CAMB-82640/2020 | EPI_ISL_443531 | 13/04/2020 |
| CAMB-8266E | hCoV-19/England/CAMB-8266E/2020 | EPI_ISL_443583 | 13/04/2020 |
| CAMB-8268C | hCoV-19/England/CAMB-8268C/2020 | EPI_ISL_443674 | 13/04/2020 |
| CAMB-826B9 | hCoV-19/England/CAMB-826B9/2020 | EPI_ISL_443475 | 13/04/2020 |
| CAMB-826F5 | hCoV-19/England/CAMB-826F5/2020 | EPI_ISL_443524 | 13/04/2020 |
| CAMB-8275C | hCoV-19/England/CAMB-8275C/2020 | EPI_ISL_443684 | 13/04/2020 |
| CAMB-827D4 | hCoV-19/England/CAMB-827D4/2020 | EPI_ISL_443581 | 13/04/2020 |
| CAMB-8281D | hCoV-19/England/CAMB-8281D/2020 | EPI_ISL_443486 | 13/04/2020 |
| CAMB-8283B | hCoV-19/England/CAMB-8283B/2020 | EPI_ISL_443407 | 13/04/2020 |
| CAMB-82DE1 | hCoV-19/England/CAMB-82DE1/2020 | EPI_ISL_443666 | 13/04/2020 |
| CAMB-82EA2 | hCoV-19/England/CAMB-82EA2/2020 | EPI_ISL_443321 | 13/04/2020 |
| CAMB-8306F | hCoV-19/England/CAMB-8306F/2020 | EPI_ISL_443634 | 13/04/2020 |
| CAMB-8313F | hCoV-19/England/CAMB-8313F/2020 | EPI_ISL_443561 | 13/04/2020 |
| CAMB-7C069 | hCoV-19/England/CAMB-7C069/2020 | EPI_ISL_433985 | 14/04/2020 |
| CAMB-7C0B4 | hCoV-19/England/CAMB-7C0B4/2020 | EPI_ISL_433990 | 14/04/2020 |
| CAMB-7C0C3 | hCoV-19/England/CAMB-7C0C3/2020 | EPI_ISL_433991 | 14/04/2020 |
| CAMB-7C0D2 | hCoV-19/England/CAMB-7C0D2/2020 | EPI_ISL_433992 | 14/04/2020 |
| CAMB-7C139 | hCoV-19/England/CAMB-7C139/2020 | EPI_ISL_433998 | 14/04/2020 |
| CAMB-7C1EE | hCoV-19/England/CAMB-7C1EE/2020 | EPI_ISL_434009 | 14/04/2020 |
| CAMB-7C209 | hCoV-19/England/CAMB-7C209/2020 | EPI_ISL_434011 | 14/04/2020 |
| CAMB-7C218 | hCoV-19/England/CAMB-7C218/2020 | EPI_ISL_434012 | 14/04/2020 |
| CAMB-7C2AF | hCoV-19/England/CAMB-7C2AF/2020 | EPI_ISL_434021 | 14/04/2020 |
| CAMB-7C306 | hCoV-19/England/CAMB-7C306/2020 | EPI_ISL_434026 | 14/04/2020 |
| CAMB-7C315 | hCoV-19/England/CAMB-7C315/2020 | EPI_ISL_434027 | 14/04/2020 |
| CAMB-7C333 | hCoV-19/England/CAMB-7C333/2020 | EPI_ISL_434029 | 14/04/2020 |
| CAMB-7C37F | hCoV-19/England/CAMB-7C37F/2020 | EPI_ISL_434033 | 14/04/2020 |
| CAMB-7C38E | hCoV-19/England/CAMB-7C38E/2020 | EPI_ISL_434034 | 14/04/2020 |
| CAMB-7C3AC | hCoV-19/England/CAMB-7C3AC/2020 | EPI_ISL_434036 | 14/04/2020 |
| CAMB-7C3CA | hCoV-19/England/CAMB-7C3CA/2020 | EPI_ISL_434038 | 14/04/2020 |
| CAMB-8224C | hCoV-19/England/CAMB-8224C/2020 | EPI_ISL_443452 | 14/04/2020 |
| CAMB-82358 | hCoV-19/England/CAMB-82358/2020 | EPI_ISL_443645 | 14/04/2020 |
| CAMB-82482 | hCoV-19/England/CAMB-82482/2020 | EPI_ISL_443450 | 14/04/2020 |
| CAMB-82491 | hCoV-19/England/CAMB-82491/2020 | EPI_ISL_443458 | 14/04/2020 |
| CAMB-824A0 | hCoV-19/England/CAMB-824A0/2020 | EPI_ISL_443438 | 14/04/2020 |
| CAMB-824BF | hCoV-19/England/CAMB-824BF/2020 | EPI_ISL_443555 | 14/04/2020 |
| CAMB-82507 | hCoV-19/England/CAMB-82507/2020 | EPI_ISL_443459 | 14/04/2020 |
| CAMB-826C8 | hCoV-19/England/CAMB-826C8/2020 | EPI_ISL_443456 | 14/04/2020 |
| CAMB-826E6 | hCoV-19/England/CAMB-826E6/2020 | EPI_ISL_443579 | 14/04/2020 |
| CAMB-82789 | hCoV-19/England/CAMB-82789/2020 | EPI_ISL_443496 | 14/04/2020 |
| CAMB-827A7 | hCoV-19/England/CAMB-827A7/2020 | EPI_ISL_443464 | 14/04/2020 |
| CAMB-82859 | hCoV-19/England/CAMB-82859/2020 | EPI_ISL_443576 | 14/04/2020 |
| CAMB-828D1 | hCoV-19/England/CAMB-828D1/2020 | EPI_ISL_443417 | 14/04/2020 |
| CAMB-82974 | hCoV-19/England/CAMB-82974/2020 | EPI_ISL_443535 | 14/04/2020 |
| CAMB-82D5A | hCoV-19/England/CAMB-82D5A/2020 | EPI_ISL_443542 | 14/04/2020 |
| CAMB-82DC3 | hCoV-19/England/CAMB-82DC3/2020 | EPI_ISL_443560 | 14/04/2020 |
| CAMB-82DD2 | hCoV-19/England/CAMB-82DD2/2020 | EPI_ISL_443536 | 14/04/2020 |
| CAMB-82DF0 | hCoV-19/England/CAMB-82DF0/2020 | EPI_ISL_443567 | 14/04/2020 |
| CAMB-8307E | hCoV-19/England/CAMB-8307E/2020 | EPI_ISL_443522 | 14/04/2020 |
| CAMB-830AB | hCoV-19/England/CAMB-830AB/2020 | EPI_ISL_443402 | 14/04/2020 |
| CAMB-830BA | hCoV-19/England/CAMB-830BA/2020 | EPI_ISL_443654 | 14/04/2020 |
| CAMB-830F6 | hCoV-19/England/CAMB-830F6/2020 | EPI_ISL_443433 | 14/04/2020 |
| CAMB-831A8 | hCoV-19/England/CAMB-831A8/2020 | EPI_ISL_443619 | 14/04/2020 |
| CAMB-831E4 | hCoV-19/England/CAMB-831E4/2020 | EPI_ISL_443429 | 14/04/2020 |
| CAMB-8322D | hCoV-19/England/CAMB-8322D/2020 | EPI_ISL_443485 | 14/04/2020 |
| CAMB-83296 | hCoV-19/England/CAMB-83296/2020 | EPI_ISL_443506 | 14/04/2020 |
| CAMB-832A5 | hCoV-19/England/CAMB-832A5/2020 | EPI_ISL_443685 | 14/04/2020 |
| CAMB-832B4 | hCoV-19/England/CAMB-832B4/2020 | EPI_ISL_443618 | 14/04/2020 |
| CAMB-8331B | hCoV-19/England/CAMB-8331B/2020 | EPI_ISL_591078 | 14/04/2020 |
| CAMB-833DF | hCoV-19/England/CAMB-833DF/2020 | EPI_ISL_443415 | 14/04/2020 |
| CAMB-833EE | hCoV-19/England/CAMB-833EE/2020 | EPI_ISL_443664 | 14/04/2020 |
| CAMB-83418 | hCoV-19/England/CAMB-83418/2020 | EPI_ISL_443527 | 14/04/2020 |
| CAMB-83560 | hCoV-19/England/CAMB-83560/2020 | EPI_ISL_443444 | 14/04/2020 |
| CAMB-8358E | hCoV-19/England/CAMB-8358E/2020 | EPI_ISL_443463 | 14/04/2020 |
| CAMB-835CA | hCoV-19/England/CAMB-835CA/2020 | EPI_ISL_443394 | 14/04/2020 |
| CAMB-8369A | hCoV-19/England/CAMB-8369A/2020 | EPI_ISL_443636 | 14/04/2020 |
| CAMB-7C263 | hCoV-19/England/CAMB-7C263/2020 | EPI_ISL_434017 | 15/04/2020 |
| CAMB-7E164 | hCoV-19/England/CAMB-7E164/2020 | EPI_ISL_434046 | 15/04/2020 |
| CAMB-7E207 | hCoV-19/England/CAMB-7E207/2020 | EPI_ISL_434052 | 15/04/2020 |
| CAMB-7E243 | hCoV-19/England/CAMB-7E243/2020 | EPI_ISL_434055 | 15/04/2020 |
| CAMB-7FC80 | hCoV-19/England/CAMB-7FC80/2020 | EPI_ISL_444399 | 15/04/2020 |
| CAMB-7FC9F | hCoV-19/England/CAMB-7FC9F/2020 | EPI_ISL_444400 | 15/04/2020 |
| CAMB-7FCBD | hCoV-19/England/CAMB-7FCBD/2020 | EPI_ISL_444401 | 15/04/2020 |
| CAMB-80F29 | hCoV-19/England/CAMB-80F29/2020 | EPI_ISL_444448 | 15/04/2020 |
| CAMB-80F38 | hCoV-19/England/CAMB-80F38/2020 | EPI_ISL_444449 | 15/04/2020 |
| CAMB-80FB0 | hCoV-19/England/CAMB-80FB0/2020 | EPI_ISL_438551 | 15/04/2020 |
| CAMB-83032 | hCoV-19/England/CAMB-83032/2020 | EPI_ISL_443652 | 15/04/2020 |
| CAMB-832C3 | hCoV-19/England/CAMB-832C3/2020 | EPI_ISL_443396 | 15/04/2020 |
| CAMB-83445 | hCoV-19/England/CAMB-83445/2020 | EPI_ISL_443638 | 15/04/2020 |
| CAMB-834CD | hCoV-19/England/CAMB-834CD/2020 | EPI_ISL_443395 | 15/04/2020 |
| CAMB-834FA | hCoV-19/England/CAMB-834FA/2020 | EPI_ISL_443608 | 15/04/2020 |
| CAMB-8357F | hCoV-19/England/CAMB-8357F/2020 | EPI_ISL_443422 | 15/04/2020 |
| CAMB-836D6 | hCoV-19/England/CAMB-836D6/2020 | EPI_ISL_443435 | 15/04/2020 |
| CAMB-83779 | hCoV-19/England/CAMB-83779/2020 | EPI_ISL_443643 | 15/04/2020 |
| CAMB-83797 | hCoV-19/England/CAMB-83797/2020 | EPI_ISL_443446 | 15/04/2020 |
| CAMB-837B5 | hCoV-19/England/CAMB-837B5/2020 | EPI_ISL_443572 | 15/04/2020 |
| CAMB-8383A | hCoV-19/England/CAMB-8383A/2020 | EPI_ISL_443683 | 15/04/2020 |
| CAMB-839CE | hCoV-19/England/CAMB-839CE/2020 | EPI_ISL_443337 | 15/04/2020 |
| CAMB-83A34 | hCoV-19/England/CAMB-83A34/2020 | EPI_ISL_443593 | 15/04/2020 |
| CAMB-83AAD | hCoV-19/England/CAMB-83AAD/2020 | EPI_ISL_443338 | 15/04/2020 |
| CAMB-83ADA | hCoV-19/England/CAMB-83ADA/2020 | EPI_ISL_443340 | 15/04/2020 |
| CAMB-83B13 | hCoV-19/England/CAMB-83B13/2020 | EPI_ISL_591091 | 15/04/2020 |
| CAMB-83B6E | hCoV-19/England/CAMB-83B6E/2020 | EPI_ISL_443336 | 15/04/2020 |
| CAMB-83BC8 | hCoV-19/England/CAMB-83BC8/2020 | EPI_ISL_443349 | 15/04/2020 |
| CAMB-83C98 | hCoV-19/England/CAMB-83C98/2020 | EPI_ISL_443350 | 15/04/2020 |
| CAMB-8422C | hCoV-19/England/CAMB-8422C/2020 | EPI_ISL_443454 | 15/04/2020 |
| CAMB-84611 | hCoV-19/England/CAMB-84611/2020 | EPI_ISL_591076 | 15/04/2020 |
| CAMB-1AA2D4 | hCoV-19/England/CAMB-1AA2D4/2020 | EPI_ISL_443408 | 16/04/2020 |
| CAMB-1AA30E | hCoV-19/England/CAMB-1AA30E/2020 | EPI_ISL_443656 | 16/04/2020 |
| CAMB-1AA359 | hCoV-19/England/CAMB-1AA359/2020 | EPI_ISL_443517 | 16/04/2020 |
| CAMB-1AA3E0 | hCoV-19/England/CAMB-1AA3E0/2020 | EPI_ISL_443448 | 16/04/2020 |
| CAMB-7E137 | hCoV-19/England/CAMB-7E137/2020 | EPI_ISL_434043 | 16/04/2020 |
| CAMB-7E146 | hCoV-19/England/CAMB-7E146/2020 | EPI_ISL_434044 | 16/04/2020 |
| CAMB-7E155 | hCoV-19/England/CAMB-7E155/2020 | EPI_ISL_434045 | 16/04/2020 |
| CAMB-7E1BF | hCoV-19/England/CAMB-7E1BF/2020 | EPI_ISL_434047 | 16/04/2020 |
| CAMB-7E1DD | hCoV-19/England/CAMB-7E1DD/2020 | EPI_ISL_434049 | 16/04/2020 |
| CAMB-7FBB0 | hCoV-19/England/CAMB-7FBB0/2020 | EPI_ISL_433487 | 16/04/2020 |
| CAMB-7FBCF | hCoV-19/England/CAMB-7FBCF/2020 | EPI_ISL_433488 | 16/04/2020 |
| CAMB-7FBDE | hCoV-19/England/CAMB-7FBDE/2020 | EPI_ISL_433489 | 16/04/2020 |
| CAMB-7FBED | hCoV-19/England/CAMB-7FBED/2020 | EPI_ISL_444397 | 16/04/2020 |
| CAMB-7FBFC | hCoV-19/England/CAMB-7FBFC/2020 | EPI_ISL_433490 | 16/04/2020 |
| CAMB-7FC08 | hCoV-19/England/CAMB-7FC08/2020 | EPI_ISL_433491 | 16/04/2020 |
| CAMB-7FC17 | hCoV-19/England/CAMB-7FC17/2020 | EPI_ISL_433492 | 16/04/2020 |
| CAMB-7FC26 | hCoV-19/England/CAMB-7FC26/2020 | EPI_ISL_433493 | 16/04/2020 |
| CAMB-7FC35 | hCoV-19/England/CAMB-7FC35/2020 | EPI_ISL_433494 | 16/04/2020 |
| CAMB-7FC44 | hCoV-19/England/CAMB-7FC44/2020 | EPI_ISL_433495 | 16/04/2020 |
| CAMB-7FC53 | hCoV-19/England/CAMB-7FC53/2020 | EPI_ISL_433496 | 16/04/2020 |
| CAMB-7FC62 | hCoV-19/England/CAMB-7FC62/2020 | EPI_ISL_433497 | 16/04/2020 |
| CAMB-80E95 | hCoV-19/England/CAMB-80E95/2020 | EPI_ISL_444439 | 16/04/2020 |
| CAMB-81131 | hCoV-19/England/CAMB-81131/2020 | EPI_ISL_438571 | 16/04/2020 |
| CAMB-83885 | hCoV-19/England/CAMB-83885/2020 | EPI_ISL_443598 | 16/04/2020 |
| CAMB-83991 | hCoV-19/England/CAMB-83991/2020 | EPI_ISL_443378 | 16/04/2020 |
| CAMB-83A52 | hCoV-19/England/CAMB-83A52/2020 | EPI_ISL_443354 | 16/04/2020 |
| CAMB-83BD7 | hCoV-19/England/CAMB-83BD7/2020 | EPI_ISL_443368 | 16/04/2020 |
| CAMB-83BE6 | hCoV-19/England/CAMB-83BE6/2020 | EPI_ISL_443343 | 16/04/2020 |
| CAMB-83C10 | hCoV-19/England/CAMB-83C10/2020 | EPI_ISL_443375 | 16/04/2020 |
| CAMB-83C3E | hCoV-19/England/CAMB-83C3E/2020 | EPI_ISL_443376 | 16/04/2020 |
| CAMB-83CA7 | hCoV-19/England/CAMB-83CA7/2020 | EPI_ISL_443352 | 16/04/2020 |
| CAMB-83CC5 | hCoV-19/England/CAMB-83CC5/2020 | EPI_ISL_443371 | 16/04/2020 |
| CAMB-841E3 | hCoV-19/England/CAMB-841E3/2020 | EPI_ISL_443412 | 16/04/2020 |
| CAMB-841F2 | hCoV-19/England/CAMB-841F2/2020 | EPI_ISL_443565 | 16/04/2020 |
| CAMB-8423B | hCoV-19/England/CAMB-8423B/2020 | EPI_ISL_443633 | 16/04/2020 |
| CAMB-8424A | hCoV-19/England/CAMB-8424A/2020 | EPI_ISL_443564 | 16/04/2020 |
| CAMB-84277 | hCoV-19/England/CAMB-84277/2020 | EPI_ISL_443451 | 16/04/2020 |
| CAMB-842B3 | hCoV-19/England/CAMB-842B3/2020 | EPI_ISL_443621 | 16/04/2020 |
| CAMB-842C2 | hCoV-19/England/CAMB-842C2/2020 | EPI_ISL_443484 | 16/04/2020 |
| CAMB-84329 | hCoV-19/England/CAMB-84329/2020 | EPI_ISL_443635 | 16/04/2020 |
| CAMB-84356 | hCoV-19/England/CAMB-84356/2020 | EPI_ISL_443614 | 16/04/2020 |
| CAMB-84365 | hCoV-19/England/CAMB-84365/2020 | EPI_ISL_443418 | 16/04/2020 |
| CAMB-843B0 | hCoV-19/England/CAMB-843B0/2020 | EPI_ISL_443574 | 16/04/2020 |
| CAMB-843DE | hCoV-19/England/CAMB-843DE/2020 | EPI_ISL_443403 | 16/04/2020 |
| CAMB-843FC | hCoV-19/England/CAMB-843FC/2020 | EPI_ISL_443566 | 16/04/2020 |
| CAMB-84435 | hCoV-19/England/CAMB-84435/2020 | EPI_ISL_443632 | 16/04/2020 |
| CAMB-84453 | hCoV-19/England/CAMB-84453/2020 | EPI_ISL_443510 | 16/04/2020 |
| CAMB-8456F | hCoV-19/England/CAMB-8456F/2020 | EPI_ISL_443660 | 16/04/2020 |
| CAMB-8458D | hCoV-19/England/CAMB-8458D/2020 | EPI_ISL_443525 | 16/04/2020 |
| CAMB-845C9 | hCoV-19/England/CAMB-845C9/2020 | EPI_ISL_443668 | 16/04/2020 |
| CAMB-1AA2C5 | hCoV-19/England/CAMB-1AA2C5/2020 | EPI_ISL_443414 | 17/04/2020 |
| CAMB-1AA2E3 | hCoV-19/England/CAMB-1AA2E3/2020 | EPI_ISL_443676 | 17/04/2020 |
| CAMB-1AA368 | hCoV-19/England/CAMB-1AA368/2020 | EPI_ISL_443663 | 17/04/2020 |
| CAMB-1AA438 | hCoV-19/England/CAMB-1AA438/2020 | EPI_ISL_443413 | 17/04/2020 |
| CAMB-1AA4DE | hCoV-19/England/CAMB-1AA4DE/2020 | EPI_ISL_443514 | 17/04/2020 |
| CAMB-1AA5EA | hCoV-19/England/CAMB-1AA5EA/2020 | EPI_ISL_470162 | 17/04/2020 |
| CAMB-1AA614 | hCoV-19/England/CAMB-1AA614/2020 | EPI_ISL_470279 | 17/04/2020 |
| CAMB-1AA623 | hCoV-19/England/CAMB-1AA623/2020 | EPI_ISL_470165 | 17/04/2020 |
| CAMB-1AA66F | hCoV-19/England/CAMB-1AA66F/2020 | EPI_ISL_470112 | 17/04/2020 |
| CAMB-1AA69C | hCoV-19/England/CAMB-1AA69C/2020 | EPI_ISL_470200 | 17/04/2020 |
| CAMB-1AA7C6 | hCoV-19/England/CAMB-1AA7C6/2020 | EPI_ISL_470209 | 17/04/2020 |
| CAMB-1AA83C | hCoV-19/England/CAMB-1AA83C/2020 | EPI_ISL_470153 | 17/04/2020 |
| CAMB-1AAA36 | hCoV-19/England/CAMB-1AAA36/2020 | EPI_ISL_470129 | 17/04/2020 |
| CAMB-1AAE49 | hCoV-19/England/CAMB-1AAE49/2020 | EPI_ISL_444318 | 17/04/2020 |
| CAMB-1AAE58 | hCoV-19/England/CAMB-1AAE58/2020 | EPI_ISL_444319 | 17/04/2020 |
| CAMB-1AAE94 | hCoV-19/England/CAMB-1AAE94/2020 | EPI_ISL_444321 | 17/04/2020 |
| CAMB-1AAEB2 | hCoV-19/England/CAMB-1AAEB2/2020 | EPI_ISL_444322 | 17/04/2020 |
| CAMB-7E2CB | hCoV-19/England/CAMB-7E2CB/2020 | EPI_ISL_434062 | 17/04/2020 |
| CAMB-7E2F8 | hCoV-19/England/CAMB-7E2F8/2020 | EPI_ISL_433468 | 17/04/2020 |
| CAMB-7E304 | hCoV-19/England/CAMB-7E304/2020 | EPI_ISL_433469 | 17/04/2020 |
| CAMB-7E322 | hCoV-19/England/CAMB-7E322/2020 | EPI_ISL_433471 | 17/04/2020 |
| CAMB-7FB29 | hCoV-19/England/CAMB-7FB29/2020 | EPI_ISL_433479 | 17/04/2020 |
| CAMB-7FB47 | hCoV-19/England/CAMB-7FB47/2020 | EPI_ISL_433480 | 17/04/2020 |
| CAMB-7FB56 | hCoV-19/England/CAMB-7FB56/2020 | EPI_ISL_433481 | 17/04/2020 |
| CAMB-7FB65 | hCoV-19/England/CAMB-7FB65/2020 | EPI_ISL_433482 | 17/04/2020 |
| CAMB-7FB74 | hCoV-19/England/CAMB-7FB74/2020 | EPI_ISL_433483 | 17/04/2020 |
| CAMB-7FB83 | hCoV-19/England/CAMB-7FB83/2020 | EPI_ISL_433484 | 17/04/2020 |
| CAMB-7FB92 | hCoV-19/England/CAMB-7FB92/2020 | EPI_ISL_433485 | 17/04/2020 |
| CAMB-7FBA1 | hCoV-19/England/CAMB-7FBA1/2020 | EPI_ISL_433486 | 17/04/2020 |
| CAMB-80E86 | hCoV-19/England/CAMB-80E86/2020 | EPI_ISL_444438 | 17/04/2020 |
| CAMB-80EB3 | hCoV-19/England/CAMB-80EB3/2020 | EPI_ISL_444441 | 17/04/2020 |
| CAMB-80EC2 | hCoV-19/England/CAMB-80EC2/2020 | EPI_ISL_444442 | 17/04/2020 |
| CAMB-80EE0 | hCoV-19/England/CAMB-80EE0/2020 | EPI_ISL_444444 | 17/04/2020 |
| CAMB-841C5 | hCoV-19/England/CAMB-841C5/2020 | EPI_ISL_443480 | 17/04/2020 |
| CAMB-844EA | hCoV-19/England/CAMB-844EA/2020 | EPI_ISL_443620 | 17/04/2020 |
| CAMB-845AB | hCoV-19/England/CAMB-845AB/2020 | EPI_ISL_443658 | 17/04/2020 |
| CAMB-1AA5BD | hCoV-19/England/CAMB-1AA5BD/2020 | EPI_ISL_470092 | 18/04/2020 |
| CAMB-1AA92A | hCoV-19/England/CAMB-1AA92A/2020 | EPI_ISL_470346 | 18/04/2020 |
| CAMB-1AA948 | hCoV-19/England/CAMB-1AA948/2020 | EPI_ISL_470243 | 18/04/2020 |
| CAMB-1AAA54 | hCoV-19/England/CAMB-1AAA54/2020 | EPI_ISL_470340 | 18/04/2020 |
| CAMB-1AAA90 | hCoV-19/England/CAMB-1AAA90/2020 | EPI_ISL_470235 | 18/04/2020 |
| CAMB-1AAB06 | hCoV-19/England/CAMB-1AAB06/2020 | EPI_ISL_470191 | 18/04/2020 |
| CAMB-1AAB24 | hCoV-19/England/CAMB-1AAB24/2020 | EPI_ISL_470144 | 18/04/2020 |
| CAMB-1AABAC | hCoV-19/England/CAMB-1AABAC/2020 | EPI_ISL_470155 | 18/04/2020 |
| CAMB-1AABD9 | hCoV-19/England/CAMB-1AABD9/2020 | EPI_ISL_470190 | 18/04/2020 |
| CAMB-1AABE8 | hCoV-19/England/CAMB-1AABE8/2020 | EPI_ISL_470104 | 18/04/2020 |
| CAMB-1AACF4 | hCoV-19/England/CAMB-1AACF4/2020 | EPI_ISL_470250 | 18/04/2020 |
| CAMB-1AAD79 | hCoV-19/England/CAMB-1AAD79/2020 | EPI_ISL_470282 | 18/04/2020 |
| CAMB-1AAE1C | hCoV-19/England/CAMB-1AAE1C/2020 | EPI_ISL_470260 | 18/04/2020 |
| CAMB-1AB21F | hCoV-19/England/CAMB-1AB21F/2020 | EPI_ISL_470241 | 18/04/2020 |
| CAMB-7F88C | hCoV-19/England/CAMB-7F88C/2020 | EPI_ISL_433477 | 18/04/2020 |
| CAMB-7FD23 | hCoV-19/England/CAMB-7FD23/2020 | EPI_ISL_444406 | 18/04/2020 |
| CAMB-80E59 | hCoV-19/England/CAMB-80E59/2020 | EPI_ISL_444435 | 18/04/2020 |
| CAMB-1AA8E1 | hCoV-19/England/CAMB-1AA8E1/2020 | EPI_ISL_470272 | 19/04/2020 |
| CAMB-1AAE0D | hCoV-19/England/CAMB-1AAE0D/2020 | EPI_ISL_470326 | 19/04/2020 |
| CAMB-1AB112 | hCoV-19/England/CAMB-1AB112/2020 | EPI_ISL_470273 | 19/04/2020 |
| CAMB-1AB18B | hCoV-19/England/CAMB-1AB18B/2020 | EPI_ISL_470177 | 19/04/2020 |
| CAMB-1AB19A | hCoV-19/England/CAMB-1AB19A/2020 | EPI_ISL_470334 | 19/04/2020 |
| CAMB-1AB1A9 | hCoV-19/England/CAMB-1AB1A9/2020 | EPI_ISL_470305 | 19/04/2020 |
| CAMB-1AB4FB | hCoV-19/England/CAMB-1AB4FB/2020 | EPI_ISL_470263 | 19/04/2020 |
| CAMB-1ABB6F | hCoV-19/England/CAMB-1ABB6F/2020 | EPI_ISL_470284 | 19/04/2020 |
| CAMB-1ABC6C | hCoV-19/England/CAMB-1ABC6C/2020 | EPI_ISL_470261 | 19/04/2020 |
| CAMB-7FD05 | hCoV-19/England/CAMB-7FD05/2020 | EPI_ISL_444405 | 19/04/2020 |
| CAMB-7FE20 | hCoV-19/England/CAMB-7FE20/2020 | EPI_ISL_444421 | 19/04/2020 |
| CAMB-7FE3F | hCoV-19/England/CAMB-7FE3F/2020 | EPI_ISL_444422 | 19/04/2020 |
| CAMB-7FEE4 | hCoV-19/England/CAMB-7FEE4/2020 | EPI_ISL_444431 | 19/04/2020 |
| CAMB-81061 | hCoV-19/England/CAMB-81061/2020 | EPI_ISL_438560 | 19/04/2020 |
| CAMB-81140 | hCoV-19/England/CAMB-81140/2020 | EPI_ISL_438572 | 19/04/2020 |
| CAMB-1AAE67 | hCoV-19/England/CAMB-1AAE67/2020 | EPI_ISL_444320 | 20/04/2020 |
| CAMB-1AB1F4 | hCoV-19/England/CAMB-1AB1F4/2020 | EPI_ISL_470265 | 20/04/2020 |
| CAMB-1AB23D | hCoV-19/England/CAMB-1AB23D/2020 | EPI_ISL_470276 | 20/04/2020 |
| CAMB-1AB2B5 | hCoV-19/England/CAMB-1AB2B5/2020 | EPI_ISL_470297 | 20/04/2020 |
| CAMB-1AB3C1 | hCoV-19/England/CAMB-1AB3C1/2020 | EPI_ISL_470204 | 20/04/2020 |
| CAMB-1AB4DD | hCoV-19/England/CAMB-1AB4DD/2020 | EPI_ISL_470245 | 20/04/2020 |
| CAMB-1AB895 | hCoV-19/England/CAMB-1AB895/2020 | EPI_ISL_470318 | 20/04/2020 |
| CAMB-1ABAAE | hCoV-19/England/CAMB-1ABAAE/2020 | EPI_ISL_470142 | 20/04/2020 |
| CAMB-1ABB32 | hCoV-19/England/CAMB-1ABB32/2020 | EPI_ISL_470151 | 20/04/2020 |
| CAMB-1ABB41 | hCoV-19/England/CAMB-1ABB41/2020 | EPI_ISL_470157 | 20/04/2020 |
| CAMB-1ABBAB | hCoV-19/England/CAMB-1ABBAB/2020 | EPI_ISL_470266 | 20/04/2020 |
| CAMB-1ABBF6 | hCoV-19/England/CAMB-1ABBF6/2020 | EPI_ISL_470186 | 20/04/2020 |
| CAMB-1ABC3F | hCoV-19/England/CAMB-1ABC3F/2020 | EPI_ISL_470271 | 20/04/2020 |
| CAMB-1ABC5D | hCoV-19/England/CAMB-1ABC5D/2020 | EPI_ISL_470323 | 20/04/2020 |
| CAMB-1ABCC6 | hCoV-19/England/CAMB-1ABCC6/2020 | EPI_ISL_470291 | 20/04/2020 |
| CAMB-1ABD5A | hCoV-19/England/CAMB-1ABD5A/2020 | EPI_ISL_470320 | 20/04/2020 |
| CAMB-79943 | hCoV-19/England/CAMB-79943/2020 | EPI_ISL_444393 | 20/04/2020 |
| CAMB-7FD32 | hCoV-19/England/CAMB-7FD32/2020 | EPI_ISL_444407 | 20/04/2020 |
| CAMB-7FD6F | hCoV-19/England/CAMB-7FD6F/2020 | EPI_ISL_444409 | 20/04/2020 |
| CAMB-80AFB | hCoV-19/England/CAMB-80AFB/2020 | EPI_ISL_444432 | 20/04/2020 |
| CAMB-80FDE | hCoV-19/England/CAMB-80FDE/2020 | EPI_ISL_438553 | 20/04/2020 |
| CAMB-80FFC | hCoV-19/England/CAMB-80FFC/2020 | EPI_ISL_438554 | 20/04/2020 |
| CAMB-81007 | hCoV-19/England/CAMB-81007/2020 | EPI_ISL_438555 | 20/04/2020 |
| CAMB-81016 | hCoV-19/England/CAMB-81016/2020 | EPI_ISL_438556 | 20/04/2020 |
| CAMB-81025 | hCoV-19/England/CAMB-81025/2020 | EPI_ISL_438557 | 20/04/2020 |
| CAMB-81043 | hCoV-19/England/CAMB-81043/2020 | EPI_ISL_438559 | 20/04/2020 |
| CAMB-8109E | hCoV-19/England/CAMB-8109E/2020 | EPI_ISL_438563 | 20/04/2020 |
| CAMB-810AD | hCoV-19/England/CAMB-810AD/2020 | EPI_ISL_438564 | 20/04/2020 |
| CAMB-81122 | hCoV-19/England/CAMB-81122/2020 | EPI_ISL_438570 | 20/04/2020 |
| CAMB-8117D | hCoV-19/England/CAMB-8117D/2020 | EPI_ISL_438574 | 20/04/2020 |
| CAMB-8118C | hCoV-19/England/CAMB-8118C/2020 | EPI_ISL_438575 | 20/04/2020 |
| CAMB-81FA0 | hCoV-19/England/CAMB-81FA0/2020 | EPI_ISL_438602 | 20/04/2020 |
| CAMB-81FCE | hCoV-19/England/CAMB-81FCE/2020 | EPI_ISL_438604 | 20/04/2020 |
| CAMB-82006 | hCoV-19/England/CAMB-82006/2020 | EPI_ISL_438608 | 20/04/2020 |
| CAMB-821A9 | hCoV-19/England/CAMB-821A9/2020 | EPI_ISL_438628 | 20/04/2020 |
| CAMB-1AB40A | hCoV-19/England/CAMB-1AB40A/2020 | EPI_ISL_470306 | 21/04/2020 |
| CAMB-1AB428 | hCoV-19/England/CAMB-1AB428/2020 | EPI_ISL_470164 | 21/04/2020 |
| CAMB-1AB491 | hCoV-19/England/CAMB-1AB491/2020 | EPI_ISL_470240 | 21/04/2020 |
| CAMB-1AB552 | hCoV-19/England/CAMB-1AB552/2020 | EPI_ISL_470208 | 21/04/2020 |
| CAMB-1AB59E | hCoV-19/England/CAMB-1AB59E/2020 | EPI_ISL_470195 | 21/04/2020 |
| CAMB-1AB5BC | hCoV-19/England/CAMB-1AB5BC/2020 | EPI_ISL_470213 | 21/04/2020 |
| CAMB-1AB5DA | hCoV-19/England/CAMB-1AB5DA/2020 | EPI_ISL_470120 | 21/04/2020 |
| CAMB-1AB631 | hCoV-19/England/CAMB-1AB631/2020 | EPI_ISL_470170 | 21/04/2020 |
| CAMB-1AB65F | hCoV-19/England/CAMB-1AB65F/2020 | EPI_ISL_470332 | 21/04/2020 |
| CAMB-1AB69B | hCoV-19/England/CAMB-1AB69B/2020 | EPI_ISL_470342 | 21/04/2020 |
| CAMB-1AB6AA | hCoV-19/England/CAMB-1AB6AA/2020 | EPI_ISL_470349 | 21/04/2020 |
| CAMB-1AB6B9 | hCoV-19/England/CAMB-1AB6B9/2020 | EPI_ISL_470324 | 21/04/2020 |
| CAMB-1AB6C8 | hCoV-19/England/CAMB-1AB6C8/2020 | EPI_ISL_470173 | 21/04/2020 |
| CAMB-1AB9ED | hCoV-19/England/CAMB-1AB9ED/2020 | EPI_ISL_470146 | 21/04/2020 |
| CAMB-1ABA26 | hCoV-19/England/CAMB-1ABA26/2020 | EPI_ISL_470296 | 21/04/2020 |
| CAMB-1ABD69 | hCoV-19/England/CAMB-1ABD69/2020 | EPI_ISL_470335 | 21/04/2020 |
| CAMB-1ABD78 | hCoV-19/England/CAMB-1ABD78/2020 | EPI_ISL_470258 | 21/04/2020 |
| CAMB-1ABD87 | hCoV-19/England/CAMB-1ABD87/2020 | EPI_ISL_470227 | 21/04/2020 |
| CAMB-1ABE2A | hCoV-19/England/CAMB-1ABE2A/2020 | EPI_ISL_470270 | 21/04/2020 |
| CAMB-81E58 | hCoV-19/England/CAMB-81E58/2020 | EPI_ISL_438585 | 21/04/2020 |
| CAMB-81F46 | hCoV-19/England/CAMB-81F46/2020 | EPI_ISL_438597 | 21/04/2020 |
| CAMB-81F64 | hCoV-19/England/CAMB-81F64/2020 | EPI_ISL_438598 | 21/04/2020 |
| CAMB-81FBF | hCoV-19/England/CAMB-81FBF/2020 | EPI_ISL_438603 | 21/04/2020 |
| CAMB-81FFB | hCoV-19/England/CAMB-81FFB/2020 | EPI_ISL_438607 | 21/04/2020 |
| CAMB-82015 | hCoV-19/England/CAMB-82015/2020 | EPI_ISL_438609 | 21/04/2020 |
| CAMB-82060 | hCoV-19/England/CAMB-82060/2020 | EPI_ISL_438614 | 21/04/2020 |
| CAMB-820AC | hCoV-19/England/CAMB-820AC/2020 | EPI_ISL_438618 | 21/04/2020 |
| CAMB-82103 | hCoV-19/England/CAMB-82103/2020 | EPI_ISL_438620 | 21/04/2020 |
| CAMB-82121 | hCoV-19/England/CAMB-82121/2020 | EPI_ISL_438621 | 21/04/2020 |
| CAMB-8214F | hCoV-19/England/CAMB-8214F/2020 | EPI_ISL_438623 | 21/04/2020 |
| CAMB-8217C | hCoV-19/England/CAMB-8217C/2020 | EPI_ISL_438625 | 21/04/2020 |
| CAMB-821C7 | hCoV-19/England/CAMB-821C7/2020 | EPI_ISL_438630 | 21/04/2020 |
| CAMB-8222E | hCoV-19/England/CAMB-8222E/2020 | EPI_ISL_438634 | 21/04/2020 |
| CAMB-82947 | hCoV-19/England/CAMB-82947/2020 | EPI_ISL_438640 | 21/04/2020 |
| CAMB-829A1 | hCoV-19/England/CAMB-829A1/2020 | EPI_ISL_438642 | 21/04/2020 |
| CAMB-1AB640 | hCoV-19/England/CAMB-1AB640/2020 | EPI_ISL_470220 | 22/04/2020 |
| CAMB-1AB66E | hCoV-19/England/CAMB-1AB66E/2020 | EPI_ISL_470099 | 22/04/2020 |
| CAMB-1ABCD5 | hCoV-19/England/CAMB-1ABCD5/2020 | EPI_ISL_470285 | 22/04/2020 |
| CAMB-1AE373 | hCoV-19/England/CAMB-1AE373/2020 | EPI_ISL_459498 | 22/04/2020 |
| CAMB-79934 | hCoV-19/England/CAMB-79934/2020 | EPI_ISL_444392 | 22/04/2020 |
| CAMB-81EC1 | hCoV-19/England/CAMB-81EC1/2020 | EPI_ISL_438591 | 22/04/2020 |
| CAMB-82A80 | hCoV-19/England/CAMB-82A80/2020 | EPI_ISL_438653 | 22/04/2020 |
| CAMB-82B23 | hCoV-19/England/CAMB-82B23/2020 | EPI_ISL_438661 | 22/04/2020 |
| CAMB-82B50 | hCoV-19/England/CAMB-82B50/2020 | EPI_ISL_438663 | 22/04/2020 |
| CAMB-82B6F | hCoV-19/England/CAMB-82B6F/2020 | EPI_ISL_438664 | 22/04/2020 |
| CAMB-83D0E | hCoV-19/England/CAMB-83D0E/2020 | EPI_ISL_438683 | 22/04/2020 |
| CAMB-83FBD | hCoV-19/England/CAMB-83FBD/2020 | EPI_ISL_438718 | 22/04/2020 |
| CAMB-829FC | hCoV-19/England/CAMB-829FC/2020 | EPI_ISL_438645 | 23/04/2020 |
| CAMB-82A9F | hCoV-19/England/CAMB-82A9F/2020 | EPI_ISL_438654 | 23/04/2020 |
| CAMB-82AAE | hCoV-19/England/CAMB-82AAE/2020 | EPI_ISL_438655 | 23/04/2020 |
| CAMB-82B32 | hCoV-19/England/CAMB-82B32/2020 | EPI_ISL_438662 | 23/04/2020 |
| CAMB-83CF2 | hCoV-19/England/CAMB-83CF2/2020 | EPI_ISL_438682 | 23/04/2020 |
| CAMB-83D59 | hCoV-19/England/CAMB-83D59/2020 | EPI_ISL_438686 | 23/04/2020 |
| CAMB-83D86 | hCoV-19/England/CAMB-83D86/2020 | EPI_ISL_438689 | 23/04/2020 |
| CAMB-1ABF72 | hCoV-19/England/CAMB-1ABF72/2020 | EPI_ISL_444352 | 24/04/2020 |
| CAMB-82CA8 | hCoV-19/England/CAMB-82CA8/2020 | EPI_ISL_438678 | 24/04/2020 |
| CAMB-83463 | hCoV-19/England/CAMB-83463/2020 | EPI_ISL_438680 | 24/04/2020 |
| CAMB-83D77 | hCoV-19/England/CAMB-83D77/2020 | EPI_ISL_438688 | 24/04/2020 |
| CAMB-83D95 | hCoV-19/England/CAMB-83D95/2020 | EPI_ISL_438690 | 24/04/2020 |
| CAMB-83DA4 | hCoV-19/England/CAMB-83DA4/2020 | EPI_ISL_438691 | 24/04/2020 |
| CAMB-83DB3 | hCoV-19/England/CAMB-83DB3/2020 | EPI_ISL_438692 | 24/04/2020 |
| CAMB-8413E | hCoV-19/England/CAMB-8413E/2020 | EPI_ISL_438740 | 24/04/2020 |
| CAMB-8415C | hCoV-19/England/CAMB-8415C/2020 | EPI_ISL_438741 | 24/04/2020 |
| CAMB-1ADD1C | hCoV-19/England/CAMB-1ADD1C/2020 | EPI_ISL_459501 | 25/04/2020 |
| CAMB-82BAB | hCoV-19/England/CAMB-82BAB/2020 | EPI_ISL_438666 | 25/04/2020 |
| CAMB-83481 | hCoV-19/England/CAMB-83481/2020 | EPI_ISL_438681 | 25/04/2020 |
| CAMB-83DD1 | hCoV-19/England/CAMB-83DD1/2020 | EPI_ISL_438694 | 25/04/2020 |
| CAMB-83E29 | hCoV-19/England/CAMB-83E29/2020 | EPI_ISL_438698 | 25/04/2020 |
| CAMB-83F17 | hCoV-19/England/CAMB-83F17/2020 | EPI_ISL_438710 | 25/04/2020 |
| CAMB-83F53 | hCoV-19/England/CAMB-83F53/2020 | EPI_ISL_438713 | 25/04/2020 |
| CAMB-83FF9 | hCoV-19/England/CAMB-83FF9/2020 | EPI_ISL_438722 | 25/04/2020 |
| CAMB-1AB7C5 | hCoV-19/England/CAMB-1AB7C5/2020 | EPI_ISL_444332 | 26/04/2020 |
| CAMB-1AB7E3 | hCoV-19/England/CAMB-1AB7E3/2020 | EPI_ISL_444333 | 26/04/2020 |
| CAMB-1ABF27 | hCoV-19/England/CAMB-1ABF27/2020 | EPI_ISL_444348 | 26/04/2020 |
| CAMB-1ADD67 | hCoV-19/England/CAMB-1ADD67/2020 | EPI_ISL_459471 | 26/04/2020 |
| CAMB-1ADE28 | hCoV-19/England/CAMB-1ADE28/2020 | EPI_ISL_459425 | 26/04/2020 |
| CAMB-1ADE55 | hCoV-19/England/CAMB-1ADE55/2020 | EPI_ISL_459496 | 26/04/2020 |
| CAMB-1ADF16 | hCoV-19/England/CAMB-1ADF16/2020 | EPI_ISL_459423 | 26/04/2020 |
| CAMB-1ADF34 | hCoV-19/England/CAMB-1ADF34/2020 | EPI_ISL_459457 | 26/04/2020 |
| CAMB-1ADF61 | hCoV-19/England/CAMB-1ADF61/2020 | EPI_ISL_459475 | 26/04/2020 |
| CAMB-1ADFBC | hCoV-19/England/CAMB-1ADFBC/2020 | EPI_ISL_459414 | 26/04/2020 |
| CAMB-1AE030 | hCoV-19/England/CAMB-1AE030/2020 | EPI_ISL_459437 | 26/04/2020 |
| CAMB-1AE04F | hCoV-19/England/CAMB-1AE04F/2020 | EPI_ISL_459462 | 26/04/2020 |
| CAMB-1AE06D | hCoV-19/England/CAMB-1AE06D/2020 | EPI_ISL_459503 | 26/04/2020 |
| CAMB-1AE2FE | hCoV-19/England/CAMB-1AE2FE/2020 | EPI_ISL_459480 | 26/04/2020 |
| CAMB-83E1A | hCoV-19/England/CAMB-83E1A/2020 | EPI_ISL_438697 | 26/04/2020 |
| CAMB-83EED | hCoV-19/England/CAMB-83EED/2020 | EPI_ISL_438708 | 26/04/2020 |
| CAMB-83F44 | hCoV-19/England/CAMB-83F44/2020 | EPI_ISL_438712 | 26/04/2020 |
| CAMB-84013 | hCoV-19/England/CAMB-84013/2020 | EPI_ISL_438724 | 26/04/2020 |
| CAMB-84040 | hCoV-19/England/CAMB-84040/2020 | EPI_ISL_438727 | 26/04/2020 |
| CAMB-8406E | hCoV-19/England/CAMB-8406E/2020 | EPI_ISL_438729 | 26/04/2020 |
| CAMB-840AA | hCoV-19/England/CAMB-840AA/2020 | EPI_ISL_438732 | 26/04/2020 |
| CAMB-840D7 | hCoV-19/England/CAMB-840D7/2020 | EPI_ISL_438735 | 26/04/2020 |
| CAMB-1AA429 | hCoV-19/England/CAMB-1AA429/2020 | EPI_ISL_444313 | 27/04/2020 |
| CAMB-1AA447 | hCoV-19/England/CAMB-1AA447/2020 | EPI_ISL_444314 | 27/04/2020 |
| CAMB-1AA465 | hCoV-19/England/CAMB-1AA465/2020 | EPI_ISL_444315 | 27/04/2020 |
| CAMB-1AAD2E | hCoV-19/England/CAMB-1AAD2E/2020 | EPI_ISL_444317 | 27/04/2020 |
| CAMB-1AB6E6 | hCoV-19/England/CAMB-1AB6E6/2020 | EPI_ISL_444327 | 27/04/2020 |
| CAMB-1ABEA2 | hCoV-19/England/CAMB-1ABEA2/2020 | EPI_ISL_444340 | 27/04/2020 |
| CAMB-1ABEC0 | hCoV-19/England/CAMB-1ABEC0/2020 | EPI_ISL_444342 | 27/04/2020 |
| CAMB-1ABF54 | hCoV-19/England/CAMB-1ABF54/2020 | EPI_ISL_444350 | 27/04/2020 |
| CAMB-1ABF81 | hCoV-19/England/CAMB-1ABF81/2020 | EPI_ISL_444353 | 27/04/2020 |
| CAMB-1ABF90 | hCoV-19/England/CAMB-1ABF90/2020 | EPI_ISL_444354 | 27/04/2020 |
| CAMB-1AC050 | hCoV-19/England/CAMB-1AC050/2020 | EPI_ISL_444364 | 27/04/2020 |
| CAMB-1AC08D | hCoV-19/England/CAMB-1AC08D/2020 | EPI_ISL_444367 | 27/04/2020 |
| CAMB-1AC199 | hCoV-19/England/CAMB-1AC199/2020 | EPI_ISL_444380 | 27/04/2020 |
| CAMB-1AC25A | hCoV-19/England/CAMB-1AC25A/2020 | EPI_ISL_444385 | 27/04/2020 |
| CAMB-1ACFDB | hCoV-19/England/CAMB-1ACFDB/2020 | EPI_ISL_447953 | 27/04/2020 |
| CAMB-1AD12F | hCoV-19/England/CAMB-1AD12F/2020 | EPI_ISL_447968 | 27/04/2020 |
| CAMB-1AD13E | hCoV-19/England/CAMB-1AD13E/2020 | EPI_ISL_447969 | 27/04/2020 |
| CAMB-1ADEDD | hCoV-19/England/CAMB-1ADEDD/2020 | EPI_ISL_459434 | 27/04/2020 |
| CAMB-1ADFCB | hCoV-19/England/CAMB-1ADFCB/2020 | EPI_ISL_459485 | 27/04/2020 |
| CAMB-1AE003 | hCoV-19/England/CAMB-1AE003/2020 | EPI_ISL_459479 | 27/04/2020 |
| CAMB-1AE05E | hCoV-19/England/CAMB-1AE05E/2020 | EPI_ISL_459466 | 27/04/2020 |
| CAMB-1AE07C | hCoV-19/England/CAMB-1AE07C/2020 | EPI_ISL_459483 | 27/04/2020 |
| CAMB-1AE0A9 | hCoV-19/England/CAMB-1AE0A9/2020 | EPI_ISL_459488 | 27/04/2020 |
| CAMB-1AE0E5 | hCoV-19/England/CAMB-1AE0E5/2020 | EPI_ISL_459454 | 27/04/2020 |
| CAMB-1AE13D | hCoV-19/England/CAMB-1AE13D/2020 | EPI_ISL_459451 | 27/04/2020 |
| CAMB-1AE197 | hCoV-19/England/CAMB-1AE197/2020 | EPI_ISL_459461 | 27/04/2020 |
| CAMB-1AE1A6 | hCoV-19/England/CAMB-1AE1A6/2020 | EPI_ISL_459500 | 27/04/2020 |
| CAMB-1AE1B5 | hCoV-19/England/CAMB-1AE1B5/2020 | EPI_ISL_459415 | 27/04/2020 |
| CAMB-1AE1D3 | hCoV-19/England/CAMB-1AE1D3/2020 | EPI_ISL_459493 | 27/04/2020 |
| CAMB-1AE1F1 | hCoV-19/England/CAMB-1AE1F1/2020 | EPI_ISL_459430 | 27/04/2020 |
| CAMB-1AE294 | hCoV-19/England/CAMB-1AE294/2020 | EPI_ISL_459436 | 27/04/2020 |
| CAMB-1AE2B2 | hCoV-19/England/CAMB-1AE2B2/2020 | EPI_ISL_459424 | 27/04/2020 |
| CAMB-1AE2D0 | hCoV-19/England/CAMB-1AE2D0/2020 | EPI_ISL_459426 | 27/04/2020 |
| CAMB-1AE522 | hCoV-19/England/CAMB-1AE522/2020 | EPI_ISL_459417 | 27/04/2020 |
| CAMB-1AE610 | hCoV-19/England/CAMB-1AE610/2020 | EPI_ISL_459419 | 27/04/2020 |
| CAMB-1AE63E | hCoV-19/England/CAMB-1AE63E/2020 | EPI_ISL_459447 | 27/04/2020 |
| CAMB-1AE65C | hCoV-19/England/CAMB-1AE65C/2020 | EPI_ISL_459449 | 27/04/2020 |
| CAMB-1AE67A | hCoV-19/England/CAMB-1AE67A/2020 | EPI_ISL_459491 | 27/04/2020 |
| CAMB-1AE698 | hCoV-19/England/CAMB-1AE698/2020 | EPI_ISL_459438 | 27/04/2020 |
| CAMB-1AE6B6 | hCoV-19/England/CAMB-1AE6B6/2020 | EPI_ISL_459499 | 27/04/2020 |
| CAMB-83F35 | hCoV-19/England/CAMB-83F35/2020 | EPI_ISL_438711 | 27/04/2020 |
| CAMB-84110 | hCoV-19/England/CAMB-84110/2020 | EPI_ISL_438739 | 27/04/2020 |
| CAMB-8463F | hCoV-19/England/CAMB-8463F/2020 | EPI_ISL_438743 | 27/04/2020 |
| CAMB-8467B | hCoV-19/England/CAMB-8467B/2020 | EPI_ISL_438747 | 27/04/2020 |
| CAMB-1AB76B | hCoV-19/England/CAMB-1AB76B/2020 | EPI_ISL_444330 | 28/04/2020 |
| CAMB-1ABEDF | hCoV-19/England/CAMB-1ABEDF/2020 | EPI_ISL_444343 | 28/04/2020 |
| CAMB-1ABFBE | hCoV-19/England/CAMB-1ABFBE/2020 | EPI_ISL_444355 | 28/04/2020 |
| CAMB-1AC005 | hCoV-19/England/CAMB-1AC005/2020 | EPI_ISL_444359 | 28/04/2020 |
| CAMB-1AC06F | hCoV-19/England/CAMB-1AC06F/2020 | EPI_ISL_444365 | 28/04/2020 |
| CAMB-1AC0F6 | hCoV-19/England/CAMB-1AC0F6/2020 | EPI_ISL_444372 | 28/04/2020 |
| CAMB-1AC111 | hCoV-19/England/CAMB-1AC111/2020 | EPI_ISL_477783 | 28/04/2020 |
| CAMB-1AC1D5 | hCoV-19/England/CAMB-1AC1D5/2020 | EPI_ISL_444382 | 28/04/2020 |
| CAMB-1AC296 | hCoV-19/England/CAMB-1AC296/2020 | EPI_ISL_444387 | 28/04/2020 |
| CAMB-1AC2B4 | hCoV-19/England/CAMB-1AC2B4/2020 | EPI_ISL_444388 | 28/04/2020 |
| CAMB-1AC2D2 | hCoV-19/England/CAMB-1AC2D2/2020 | EPI_ISL_444389 | 28/04/2020 |
| CAMB-1AD17A | hCoV-19/England/CAMB-1AD17A/2020 | EPI_ISL_447972 | 28/04/2020 |
| CAMB-1AE0C7 | hCoV-19/England/CAMB-1AE0C7/2020 | EPI_ISL_459433 | 28/04/2020 |
| CAMB-1AE382 | hCoV-19/England/CAMB-1AE382/2020 | EPI_ISL_459470 | 28/04/2020 |
| CAMB-1AE4CB | hCoV-19/England/CAMB-1AE4CB/2020 | EPI_ISL_459476 | 28/04/2020 |
| CAMB-1AE689 | hCoV-19/England/CAMB-1AE689/2020 | EPI_ISL_459472 | 28/04/2020 |
| CAMB-1AEF51 | hCoV-19/England/CAMB-1AEF51/2020 | EPI_ISL_459274 | 28/04/2020 |
| CAMB-1AC014 | hCoV-19/England/CAMB-1AC014/2020 | EPI_ISL_444360 | 29/04/2020 |
| CAMB-1AC023 | hCoV-19/England/CAMB-1AC023/2020 | EPI_ISL_444361 | 29/04/2020 |
| CAMB-1AC041 | hCoV-19/England/CAMB-1AC041/2020 | EPI_ISL_444363 | 29/04/2020 |
| CAMB-1AC1F3 | hCoV-19/England/CAMB-1AC1F3/2020 | EPI_ISL_444383 | 29/04/2020 |
| CAMB-1AC3C0 | hCoV-19/England/CAMB-1AC3C0/2020 | EPI_ISL_444391 | 29/04/2020 |
| CAMB-1ACFEA | hCoV-19/England/CAMB-1ACFEA/2020 | EPI_ISL_447954 | 29/04/2020 |
| CAMB-1AD040 | hCoV-19/England/CAMB-1AD040/2020 | EPI_ISL_447959 | 29/04/2020 |
| CAMB-1AD0AA | hCoV-19/England/CAMB-1AD0AA/2020 | EPI_ISL_447963 | 29/04/2020 |
| CAMB-1AD0B9 | hCoV-19/England/CAMB-1AD0B9/2020 | EPI_ISL_447964 | 29/04/2020 |
| CAMB-1AD0E6 | hCoV-19/England/CAMB-1AD0E6/2020 | EPI_ISL_447966 | 29/04/2020 |
| CAMB-1AD268 | hCoV-19/England/CAMB-1AD268/2020 | EPI_ISL_447983 | 29/04/2020 |
| CAMB-1AE71D | hCoV-19/England/CAMB-1AE71D/2020 | EPI_ISL_459452 | 29/04/2020 |
| CAMB-1AE847 | hCoV-19/England/CAMB-1AE847/2020 | EPI_ISL_459439 | 29/04/2020 |
| CAMB-1AE8B0 | hCoV-19/England/CAMB-1AE8B0/2020 | EPI_ISL_459266 | 29/04/2020 |
| CAMB-1AEF8E | hCoV-19/England/CAMB-1AEF8E/2020 | EPI_ISL_459252 | 29/04/2020 |
| CAMB-1AEFCA | hCoV-19/England/CAMB-1AEFCA/2020 | EPI_ISL_459265 | 29/04/2020 |
| CAMB-1AF07B | hCoV-19/England/CAMB-1AF07B/2020 | EPI_ISL_459288 | 29/04/2020 |
| CAMB-1AF15A | hCoV-19/England/CAMB-1AF15A/2020 | EPI_ISL_459319 | 29/04/2020 |
| CAMB-1AF1B4 | hCoV-19/England/CAMB-1AF1B4/2020 | EPI_ISL_459248 | 29/04/2020 |
| CAMB-1AD004 | hCoV-19/England/CAMB-1AD004/2020 | EPI_ISL_447956 | 30/04/2020 |
| CAMB-1AD259 | hCoV-19/England/CAMB-1AD259/2020 | EPI_ISL_447982 | 30/04/2020 |
| CAMB-1AD2C2 | hCoV-19/England/CAMB-1AD2C2/2020 | EPI_ISL_447987 | 30/04/2020 |
| CAMB-1AD7B4 | hCoV-19/England/CAMB-1AD7B4/2020 | EPI_ISL_448008 | 30/04/2020 |
| CAMB-1AD893 | hCoV-19/England/CAMB-1AD893/2020 | EPI_ISL_448018 | 30/04/2020 |
| CAMB-1AE601 | hCoV-19/England/CAMB-1AE601/2020 | EPI_ISL_459418 | 30/04/2020 |
| CAMB-1AE72C | hCoV-19/England/CAMB-1AE72C/2020 | EPI_ISL_459490 | 30/04/2020 |
| CAMB-1AE7C2 | hCoV-19/England/CAMB-1AE7C2/2020 | EPI_ISL_459422 | 30/04/2020 |
| CAMB-1AF03F | hCoV-19/England/CAMB-1AF03F/2020 | EPI_ISL_459246 | 30/04/2020 |
| CAMB-1AF05D | hCoV-19/England/CAMB-1AF05D/2020 | EPI_ISL_459296 | 30/04/2020 |
| CAMB-1AF08A | hCoV-19/England/CAMB-1AF08A/2020 | EPI_ISL_459245 | 30/04/2020 |
| CAMB-1AF1F0 | hCoV-19/England/CAMB-1AF1F0/2020 | EPI_ISL_459313 | 30/04/2020 |
| CAMB-1AF293 | hCoV-19/England/CAMB-1AF293/2020 | EPI_ISL_459247 | 30/04/2020 |
| CAMB-1B0235 | hCoV-19/England/CAMB-1B0235/2020 | EPI_ISL_459256 | 30/04/2020 |
| CAMB-1B0244 | hCoV-19/England/CAMB-1B0244/2020 | EPI_ISL_459259 | 30/04/2020 |
| CAMB-1B02DB | hCoV-19/England/CAMB-1B02DB/2020 | EPI_ISL_459263 | 30/04/2020 |
| CAMB-1AC357 | hCoV-19/England/CAMB-1AC357/2020 | EPI_ISL_447951 | 01/05/2020 |
| CAMB-1AD6A8 | hCoV-19/England/CAMB-1AD6A8/2020 | EPI_ISL_447994 | 01/05/2020 |
| CAMB-1AD6B7 | hCoV-19/England/CAMB-1AD6B7/2020 | EPI_ISL_447995 | 01/05/2020 |
| CAMB-1AD70F | hCoV-19/England/CAMB-1AD70F/2020 | EPI_ISL_447999 | 01/05/2020 |
| CAMB-1AD796 | hCoV-19/England/CAMB-1AD796/2020 | EPI_ISL_448006 | 01/05/2020 |
| CAMB-1AD7A5 | hCoV-19/England/CAMB-1AD7A5/2020 | EPI_ISL_448007 | 01/05/2020 |
| CAMB-1AD82A | hCoV-19/England/CAMB-1AD82A/2020 | EPI_ISL_448015 | 01/05/2020 |
| CAMB-1AD8DF | hCoV-19/England/CAMB-1AD8DF/2020 | EPI_ISL_448021 | 01/05/2020 |
| CAMB-1AD909 | hCoV-19/England/CAMB-1AD909/2020 | EPI_ISL_448023 | 01/05/2020 |
| CAMB-1AD963 | hCoV-19/England/CAMB-1AD963/2020 | EPI_ISL_448028 | 01/05/2020 |
| CAMB-1ADAD9 | hCoV-19/England/CAMB-1ADAD9/2020 | EPI_ISL_448032 | 01/05/2020 |
| CAMB-1ADAF7 | hCoV-19/England/CAMB-1ADAF7/2020 | EPI_ISL_448033 | 01/05/2020 |
| CAMB-1ADB12 | hCoV-19/England/CAMB-1ADB12/2020 | EPI_ISL_448034 | 01/05/2020 |
| CAMB-1ADB21 | hCoV-19/England/CAMB-1ADB21/2020 | EPI_ISL_448035 | 01/05/2020 |
| CAMB-1AE9BD | hCoV-19/England/CAMB-1AE9BD/2020 | EPI_ISL_459280 | 01/05/2020 |
| CAMB-1AEBB7 | hCoV-19/England/CAMB-1AEBB7/2020 | EPI_ISL_448056 | 01/05/2020 |
| CAMB-1AF61F | hCoV-19/England/CAMB-1AF61F/2020 | EPI_ISL_459241 | 01/05/2020 |
| CAMB-1AF63D | hCoV-19/England/CAMB-1AF63D/2020 | EPI_ISL_459317 | 01/05/2020 |
| CAMB-1AF65B | hCoV-19/England/CAMB-1AF65B/2020 | EPI_ISL_459269 | 01/05/2020 |
| CAMB-1AF679 | hCoV-19/England/CAMB-1AF679/2020 | EPI_ISL_459287 | 01/05/2020 |
| CAMB-1AF846 | hCoV-19/England/CAMB-1AF846/2020 | EPI_ISL_459272 | 01/05/2020 |
| CAMB-1AF864 | hCoV-19/England/CAMB-1AF864/2020 | EPI_ISL_459249 | 01/05/2020 |
| CAMB-1AF891 | hCoV-19/England/CAMB-1AF891/2020 | EPI_ISL_459307 | 01/05/2020 |
| CAMB-1AF925 | hCoV-19/England/CAMB-1AF925/2020 | EPI_ISL_459251 | 01/05/2020 |
| CAMB-1AF961 | hCoV-19/England/CAMB-1AF961/2020 | EPI_ISL_459277 | 01/05/2020 |
| CAMB-1AFA7D | hCoV-19/England/CAMB-1AFA7D/2020 | EPI_ISL_459278 | 01/05/2020 |
| CAMB-1AFBC5 | hCoV-19/England/CAMB-1AFBC5/2020 | EPI_ISL_459219 | 01/05/2020 |
| CAMB-1AFC2C | hCoV-19/England/CAMB-1AFC2C/2020 | EPI_ISL_459211 | 01/05/2020 |
| CAMB-1AFC4A | hCoV-19/England/CAMB-1AFC4A/2020 | EPI_ISL_459168 | 01/05/2020 |
| CAMB-1AFDA1 | hCoV-19/England/CAMB-1AFDA1/2020 | EPI_ISL_459233 | 01/05/2020 |
| CAMB-1B02CC | hCoV-19/England/CAMB-1B02CC/2020 | EPI_ISL_459270 | 01/05/2020 |
| CAMB-1B03C9 | hCoV-19/England/CAMB-1B03C9/2020 | EPI_ISL_459284 | 01/05/2020 |
| CAMB-1B03D8 | hCoV-19/England/CAMB-1B03D8/2020 | EPI_ISL_459243 | 01/05/2020 |
| CAMB-1B0402 | hCoV-19/England/CAMB-1B0402/2020 | EPI_ISL_459318 | 01/05/2020 |
| CAMB-1B0411 | hCoV-19/England/CAMB-1B0411/2020 | EPI_ISL_459281 | 01/05/2020 |
| CAMB-1B047B | hCoV-19/England/CAMB-1B047B/2020 | EPI_ISL_459250 | 01/05/2020 |
| CAMB-1B048A | hCoV-19/England/CAMB-1B048A/2020 | EPI_ISL_459275 | 01/05/2020 |
| CAMB-1AD7E1 | hCoV-19/England/CAMB-1AD7E1/2020 | EPI_ISL_448011 | 02/05/2020 |
| CAMB-1AD80C | hCoV-19/England/CAMB-1AD80C/2020 | EPI_ISL_448013 | 02/05/2020 |
| CAMB-1AD936 | hCoV-19/England/CAMB-1AD936/2020 | EPI_ISL_448026 | 02/05/2020 |
| CAMB-1AD972 | hCoV-19/England/CAMB-1AD972/2020 | EPI_ISL_448029 | 02/05/2020 |
| CAMB-1ADAE8 | hCoV-19/England/CAMB-1ADAE8/2020 | not_in_GISAID | 02/05/2020 |
| CAMB-1ADB30 | hCoV-19/England/CAMB-1ADB30/2020 | EPI_ISL_448036 | 02/05/2020 |
| CAMB-1AEBC6 | hCoV-19/England/CAMB-1AEBC6/2020 | EPI_ISL_448057 | 02/05/2020 |
| CAMB-1AFA5F | hCoV-19/England/CAMB-1AFA5F/2020 | EPI_ISL_459293 | 02/05/2020 |
| CAMB-1AFAE6 | hCoV-19/England/CAMB-1AFAE6/2020 | EPI_ISL_459312 | 02/05/2020 |
| CAMB-1AFB2F | hCoV-19/England/CAMB-1AFB2F/2020 | EPI_ISL_459184 | 02/05/2020 |
| CAMB-1AFB7A | hCoV-19/England/CAMB-1AFB7A/2020 | EPI_ISL_459194 | 02/05/2020 |
| CAMB-1AFB98 | hCoV-19/England/CAMB-1AFB98/2020 | EPI_ISL_459172 | 02/05/2020 |
| CAMB-1B01DE | hCoV-19/England/CAMB-1B01DE/2020 | EPI_ISL_459282 | 02/05/2020 |
| CAMB-1B0217 | hCoV-19/England/CAMB-1B0217/2020 | EPI_ISL_459315 | 02/05/2020 |
| CAMB-1B02F9 | hCoV-19/England/CAMB-1B02F9/2020 | EPI_ISL_459254 | 02/05/2020 |
| CAMB-1B0341 | hCoV-19/England/CAMB-1B0341/2020 | EPI_ISL_459311 | 02/05/2020 |
| CAMB-1AD981 | hCoV-19/England/CAMB-1AD981/2020 | EPI_ISL_448030 | 03/05/2020 |
| CAMB-1ADB5E | hCoV-19/England/CAMB-1ADB5E/2020 | EPI_ISL_448037 | 03/05/2020 |
| CAMB-1AFB01 | hCoV-19/England/CAMB-1AFB01/2020 | EPI_ISL_459294 | 03/05/2020 |
| CAMB-1AFB10 | hCoV-19/England/CAMB-1AFB10/2020 | EPI_ISL_459186 | 03/05/2020 |
| CAMB-1AD8B1 | hCoV-19/England/CAMB-1AD8B1/2020 | EPI_ISL_448020 | 04/05/2020 |
| CAMB-1ADBB8 | hCoV-19/England/CAMB-1ADBB8/2020 | EPI_ISL_448043 | 04/05/2020 |
| CAMB-1ADBC7 | hCoV-19/England/CAMB-1ADBC7/2020 | EPI_ISL_448044 | 04/05/2020 |
| CAMB-1ADBE5 | hCoV-19/England/CAMB-1ADBE5/2020 | EPI_ISL_448046 | 04/05/2020 |
| CAMB-1AE391 | hCoV-19/England/CAMB-1AE391/2020 | EPI_ISL_448051 | 04/05/2020 |
| CAMB-1AEBD5 | hCoV-19/England/CAMB-1AEBD5/2020 | EPI_ISL_448058 | 04/05/2020 |
| CAMB-1AEBE4 | hCoV-19/England/CAMB-1AEBE4/2020 | EPI_ISL_448059 | 04/05/2020 |
| CAMB-1AFB6B | hCoV-19/England/CAMB-1AFB6B/2020 | EPI_ISL_459179 | 04/05/2020 |
| CAMB-1AFCE0 | hCoV-19/England/CAMB-1AFCE0/2020 | EPI_ISL_459225 | 04/05/2020 |
| CAMB-1AFD0B | hCoV-19/England/CAMB-1AFD0B/2020 | EPI_ISL_459221 | 04/05/2020 |
| CAMB-1AFD29 | hCoV-19/England/CAMB-1AFD29/2020 | EPI_ISL_459203 | 04/05/2020 |
| CAMB-1AFD47 | hCoV-19/England/CAMB-1AFD47/2020 | EPI_ISL_459212 | 04/05/2020 |
| CAMB-1AFD92 | hCoV-19/England/CAMB-1AFD92/2020 | EPI_ISL_459198 | 04/05/2020 |
| CAMB-1AFDB0 | hCoV-19/England/CAMB-1AFDB0/2020 | EPI_ISL_459170 | 04/05/2020 |
| CAMB-1AFDDE | hCoV-19/England/CAMB-1AFDDE/2020 | EPI_ISL_459189 | 04/05/2020 |
| CAMB-1AFDFC | hCoV-19/England/CAMB-1AFDFC/2020 | EPI_ISL_459239 | 04/05/2020 |
| CAMB-1AFE17 | hCoV-19/England/CAMB-1AFE17/2020 | EPI_ISL_459230 | 04/05/2020 |
| CAMB-1B0077 | hCoV-19/England/CAMB-1B0077/2020 | EPI_ISL_448104 | 04/05/2020 |
| CAMB-1B0851 | hCoV-19/England/CAMB-1B0851/2020 | EPI_ISL_459190 | 04/05/2020 |
| CAMB-1B087F | hCoV-19/England/CAMB-1B087F/2020 | EPI_ISL_459169 | 04/05/2020 |
| CAMB-1B08AC | hCoV-19/England/CAMB-1B08AC/2020 | EPI_ISL_459183 | 04/05/2020 |
| CAMB-1B08E8 | hCoV-19/England/CAMB-1B08E8/2020 | EPI_ISL_459167 | 04/05/2020 |
| CAMB-1B1119 | hCoV-19/England/CAMB-1B1119/2020 | EPI_ISL_459227 | 04/05/2020 |
| CAMB-1ADC00 | hCoV-19/England/CAMB-1ADC00/2020 | EPI_ISL_448048 | 05/05/2020 |
| CAMB-1AE3CE | hCoV-19/England/CAMB-1AE3CE/2020 | EPI_ISL_448053 | 05/05/2020 |
| CAMB-1AEB6C | hCoV-19/England/CAMB-1AEB6C/2020 | not_in_GISAID | 05/05/2020 |
| CAMB-1AEC1E | hCoV-19/England/CAMB-1AEC1E/2020 | EPI_ISL_448062 | 05/05/2020 |
| CAMB-1B099A | hCoV-19/England/CAMB-1B099A/2020 | EPI_ISL_459206 | 05/05/2020 |
| CAMB-1B102B | hCoV-19/England/CAMB-1B102B/2020 | EPI_ISL_459229 | 05/05/2020 |
| CAMB-1B1049 | hCoV-19/England/CAMB-1B1049/2020 | EPI_ISL_459175 | 05/05/2020 |
| CAMB-1B10C1 | hCoV-19/England/CAMB-1B10C1/2020 | EPI_ISL_459217 | 05/05/2020 |
| CAMB-1B10EF | hCoV-19/England/CAMB-1B10EF/2020 | EPI_ISL_459182 | 05/05/2020 |
| CAMB-1B1146 | hCoV-19/England/CAMB-1B1146/2020 | EPI_ISL_459193 | 05/05/2020 |
| CAMB-1AECD2 | hCoV-19/England/CAMB-1AECD2/2020 | EPI_ISL_448071 | 06/05/2020 |
| CAMB-1AED0C | hCoV-19/England/CAMB-1AED0C/2020 | EPI_ISL_448074 | 06/05/2020 |
| CAMB-1AED1B | hCoV-19/England/CAMB-1AED1B/2020 | EPI_ISL_448075 | 06/05/2020 |
| CAMB-1AF66A | hCoV-19/England/CAMB-1AF66A/2020 | EPI_ISL_448089 | 06/05/2020 |
| CAMB-1B000E | hCoV-19/England/CAMB-1B000E/2020 | EPI_ISL_448097 | 06/05/2020 |
| CAMB-1B001D | hCoV-19/England/CAMB-1B001D/2020 | EPI_ISL_448098 | 06/05/2020 |
| CAMB-1B011A | hCoV-19/England/CAMB-1B011A/2020 | EPI_ISL_448111 | 06/05/2020 |
| CAMB-1B0183 | hCoV-19/England/CAMB-1B0183/2020 | EPI_ISL_452884 | 06/05/2020 |
| CAMB-1B01A1 | hCoV-19/England/CAMB-1B01A1/2020 | EPI_ISL_448112 | 06/05/2020 |
| CAMB-1B05D2 | hCoV-19/England/CAMB-1B05D2/2020 | EPI_ISL_448114 | 06/05/2020 |
| CAMB-1B05F0 | hCoV-19/England/CAMB-1B05F0/2020 | EPI_ISL_448115 | 06/05/2020 |
| CAMB-1B11BF | hCoV-19/England/CAMB-1B11BF/2020 | EPI_ISL_459209 | 06/05/2020 |
| CAMB-1B1261 | hCoV-19/England/CAMB-1B1261/2020 | EPI_ISL_459178 | 06/05/2020 |
| CAMB-1B12AD | hCoV-19/England/CAMB-1B12AD/2020 | EPI_ISL_459185 | 06/05/2020 |
| CAMB-1B12E9 | hCoV-19/England/CAMB-1B12E9/2020 | EPI_ISL_459215 | 06/05/2020 |
| CAMB-1B1313 | hCoV-19/England/CAMB-1B1313/2020 | EPI_ISL_458570 | 06/05/2020 |
| CAMB-1B144D | hCoV-19/England/CAMB-1B144D/2020 | EPI_ISL_458530 | 06/05/2020 |
| CAMB-1B1489 | hCoV-19/England/CAMB-1B1489/2020 | EPI_ISL_458538 | 06/05/2020 |
| CAMB-1B14D4 | hCoV-19/England/CAMB-1B14D4/2020 | EPI_ISL_458572 | 06/05/2020 |
| CAMB-1B151D | hCoV-19/England/CAMB-1B151D/2020 | EPI_ISL_458575 | 06/05/2020 |
| CAMB-1B1586 | hCoV-19/England/CAMB-1B1586/2020 | EPI_ISL_458566 | 06/05/2020 |
| CAMB-1B15C2 | hCoV-19/England/CAMB-1B15C2/2020 | EPI_ISL_458517 | 06/05/2020 |
| CAMB-1B15E0 | hCoV-19/England/CAMB-1B15E0/2020 | EPI_ISL_458559 | 06/05/2020 |
| CAMB-1B228E | hCoV-19/England/CAMB-1B228E/2020 | EPI_ISL_469906 | 06/05/2020 |
| CAMB-1AED39 | hCoV-19/England/CAMB-1AED39/2020 | EPI_ISL_448077 | 07/05/2020 |
| CAMB-1AED57 | hCoV-19/England/CAMB-1AED57/2020 | EPI_ISL_452857 | 07/05/2020 |
| CAMB-1AED75 | hCoV-19/England/CAMB-1AED75/2020 | EPI_ISL_452859 | 07/05/2020 |
| CAMB-1AEF15 | hCoV-19/England/CAMB-1AEF15/2020 | EPI_ISL_452871 | 07/05/2020 |
| CAMB-1AEF24 | hCoV-19/England/CAMB-1AEF24/2020 | EPI_ISL_448079 | 07/05/2020 |
| CAMB-1AF56D | hCoV-19/England/CAMB-1AF56D/2020 | EPI_ISL_448081 | 07/05/2020 |
| CAMB-1AFF9C | hCoV-19/England/CAMB-1AFF9C/2020 | EPI_ISL_448091 | 07/05/2020 |
| CAMB-1AFFC9 | hCoV-19/England/CAMB-1AFFC9/2020 | EPI_ISL_448094 | 07/05/2020 |
| CAMB-1B004A | hCoV-19/England/CAMB-1B004A/2020 | EPI_ISL_448101 | 07/05/2020 |
| CAMB-1B0059 | hCoV-19/England/CAMB-1B0059/2020 | EPI_ISL_448102 | 07/05/2020 |
| CAMB-1B0693 | hCoV-19/England/CAMB-1B0693/2020 | EPI_ISL_452906 | 07/05/2020 |
| CAMB-1B0709 | hCoV-19/England/CAMB-1B0709/2020 | EPI_ISL_452913 | 07/05/2020 |
| CAMB-1B0718 | hCoV-19/England/CAMB-1B0718/2020 | EPI_ISL_452914 | 07/05/2020 |
| CAMB-1B0727 | hCoV-19/England/CAMB-1B0727/2020 | EPI_ISL_452915 | 07/05/2020 |
| CAMB-1B0745 | hCoV-19/England/CAMB-1B0745/2020 | EPI_ISL_452917 | 07/05/2020 |
| CAMB-1B0754 | hCoV-19/England/CAMB-1B0754/2020 | EPI_ISL_452918 | 07/05/2020 |
| CAMB-1B0763 | hCoV-19/England/CAMB-1B0763/2020 | EPI_ISL_452919 | 07/05/2020 |
| CAMB-1B07BE | hCoV-19/England/CAMB-1B07BE/2020 | EPI_ISL_452921 | 07/05/2020 |
| CAMB-1B07FA | hCoV-19/England/CAMB-1B07FA/2020 | EPI_ISL_452924 | 07/05/2020 |
| CAMB-1B0824 | hCoV-19/England/CAMB-1B0824/2020 | EPI_ISL_452925 | 07/05/2020 |
| CAMB-1B1155 | hCoV-19/England/CAMB-1B1155/2020 | EPI_ISL_459204 | 07/05/2020 |
| CAMB-1B1164 | hCoV-19/England/CAMB-1B1164/2020 | EPI_ISL_459176 | 07/05/2020 |
| CAMB-1B1182 | hCoV-19/England/CAMB-1B1182/2020 | EPI_ISL_459166 | 07/05/2020 |
| CAMB-1B1243 | hCoV-19/England/CAMB-1B1243/2020 | EPI_ISL_459197 | 07/05/2020 |
| CAMB-1B12F8 | hCoV-19/England/CAMB-1B12F8/2020 | EPI_ISL_459207 | 07/05/2020 |
| CAMB-1B1304 | hCoV-19/England/CAMB-1B1304/2020 | EPI_ISL_459191 | 07/05/2020 |
| CAMB-1B1331 | hCoV-19/England/CAMB-1B1331/2020 | EPI_ISL_458557 | 07/05/2020 |
| CAMB-1B13AA | hCoV-19/England/CAMB-1B13AA/2020 | EPI_ISL_458561 | 07/05/2020 |
| CAMB-1B150E | hCoV-19/England/CAMB-1B150E/2020 | EPI_ISL_458521 | 07/05/2020 |
| CAMB-1B1559 | hCoV-19/England/CAMB-1B1559/2020 | EPI_ISL_458563 | 07/05/2020 |
| CAMB-1B1717 | hCoV-19/England/CAMB-1B1717/2020 | EPI_ISL_458519 | 07/05/2020 |
| CAMB-1B1814 | hCoV-19/England/CAMB-1B1814/2020 | EPI_ISL_458532 | 07/05/2020 |
| CAMB-1B1823 | hCoV-19/England/CAMB-1B1823/2020 | EPI_ISL_458534 | 07/05/2020 |
| CAMB-1B188D | hCoV-19/England/CAMB-1B188D/2020 | EPI_ISL_458565 | 07/05/2020 |
| CAMB-1B18E7 | hCoV-19/England/CAMB-1B18E7/2020 | EPI_ISL_458547 | 07/05/2020 |
| CAMB-1B1BC0 | hCoV-19/England/CAMB-1B1BC0/2020 | EPI_ISL_458548 | 07/05/2020 |
| CAMB-1B1C27 | hCoV-19/England/CAMB-1B1C27/2020 | EPI_ISL_458562 | 07/05/2020 |
| CAMB-1B1C45 | hCoV-19/England/CAMB-1B1C45/2020 | EPI_ISL_458522 | 07/05/2020 |
| CAMB-1B1DE8 | hCoV-19/England/CAMB-1B1DE8/2020 | EPI_ISL_458567 | 07/05/2020 |
| CAMB-1AF55E | hCoV-19/England/CAMB-1AF55E/2020 | EPI_ISL_448080 | 08/05/2020 |
| CAMB-1AF5E5 | hCoV-19/England/CAMB-1AF5E5/2020 | EPI_ISL_448085 | 08/05/2020 |
| CAMB-1AF64C | hCoV-19/England/CAMB-1AF64C/2020 | EPI_ISL_448088 | 08/05/2020 |
| CAMB-1AFFBA | hCoV-19/England/CAMB-1AFFBA/2020 | EPI_ISL_448093 | 08/05/2020 |
| CAMB-1AFFE7 | hCoV-19/England/CAMB-1AFFE7/2020 | EPI_ISL_448095 | 08/05/2020 |
| CAMB-1B003B | hCoV-19/England/CAMB-1B003B/2020 | EPI_ISL_448100 | 08/05/2020 |
| CAMB-1B0648 | hCoV-19/England/CAMB-1B0648/2020 | EPI_ISL_452901 | 08/05/2020 |
| CAMB-1B0666 | hCoV-19/England/CAMB-1B0666/2020 | EPI_ISL_452903 | 08/05/2020 |
| CAMB-1B13C8 | hCoV-19/England/CAMB-1B13C8/2020 | EPI_ISL_458576 | 08/05/2020 |
| CAMB-1B1410 | hCoV-19/England/CAMB-1B1410/2020 | EPI_ISL_458520 | 08/05/2020 |
| CAMB-1B142F | hCoV-19/England/CAMB-1B142F/2020 | EPI_ISL_458549 | 08/05/2020 |
| CAMB-1B143E | hCoV-19/England/CAMB-1B143E/2020 | EPI_ISL_458533 | 08/05/2020 |
| CAMB-1B145C | hCoV-19/England/CAMB-1B145C/2020 | EPI_ISL_458523 | 08/05/2020 |
| CAMB-1B1568 | hCoV-19/England/CAMB-1B1568/2020 | EPI_ISL_458560 | 08/05/2020 |
| CAMB-1B1780 | hCoV-19/England/CAMB-1B1780/2020 | EPI_ISL_458539 | 08/05/2020 |
| CAMB-1AED84 | hCoV-19/England/CAMB-1AED84/2020 | EPI_ISL_452860 | 09/05/2020 |
| CAMB-1AED93 | hCoV-19/England/CAMB-1AED93/2020 | EPI_ISL_452861 | 09/05/2020 |
| CAMB-1AEDA2 | hCoV-19/England/CAMB-1AEDA2/2020 | EPI_ISL_452862 | 09/05/2020 |
| CAMB-1B002C | hCoV-19/England/CAMB-1B002C/2020 | EPI_ISL_448099 | 09/05/2020 |
| CAMB-1B0639 | hCoV-19/England/CAMB-1B0639/2020 | EPI_ISL_452900 | 09/05/2020 |
| CAMB-1B1629 | hCoV-19/England/CAMB-1B1629/2020 | EPI_ISL_458558 | 09/05/2020 |
| CAMB-1B1D8E | hCoV-19/England/CAMB-1B1D8E/2020 | EPI_ISL_458569 | 09/05/2020 |
| CAMB-1B1DAC | hCoV-19/England/CAMB-1B1DAC/2020 | EPI_ISL_458516 | 09/05/2020 |
| CAMB-1B1DCA | hCoV-19/England/CAMB-1B1DCA/2020 | EPI_ISL_458552 | 09/05/2020 |
| CAMB-1B1E03 | hCoV-19/England/CAMB-1B1E03/2020 | EPI_ISL_458546 | 09/05/2020 |
| CAMB-1B1E21 | hCoV-19/England/CAMB-1B1E21/2020 | EPI_ISL_458524 | 09/05/2020 |
| CAMB-1B1E6D | hCoV-19/England/CAMB-1B1E6D/2020 | EPI_ISL_458528 | 09/05/2020 |
| CAMB-1B06A2 | hCoV-19/England/CAMB-1B06A2/2020 | EPI_ISL_452907 | 10/05/2020 |
| CAMB-1B0AF1 | hCoV-19/England/CAMB-1B0AF1/2020 | EPI_ISL_452938 | 10/05/2020 |
| CAMB-1B17CC | hCoV-19/England/CAMB-1B17CC/2020 | EPI_ISL_458553 | 10/05/2020 |
| CAMB-1B17EA | hCoV-19/England/CAMB-1B17EA/2020 | EPI_ISL_458573 | 10/05/2020 |
| CAMB-1B1841 | hCoV-19/England/CAMB-1B1841/2020 | EPI_ISL_458554 | 10/05/2020 |
| CAMB-1B1C90 | hCoV-19/England/CAMB-1B1C90/2020 | EPI_ISL_458536 | 10/05/2020 |

### COG-UK authors

**Funding acquisition, leadership, supervision, metadata curation, project administration, samples, logistics, Sequencing, analysis, and Software and analysis tools:**
Dr Thomas R Connor PhD^33, 34^	, and Professor Nicholas J Loman PhD^15^.

**Leadership, supervision, sequencing, analysis, funding acquisition, metadata curation, project administration, samples, logistics, and visualisation:**

Dr Samuel C Robson Ph.D ^68^.

**Leadership, supervision, project administration, visualisation, samples, logistics, metadata curation and software and analysis tools:**

Dr Tanya Golubchik PhD ^27^.

**Leadership, supervision, metadata curation, project administration, samples, logistics sequencing and analysis:**

Dr M. Estee Torok FRCP ^8, 10^.

**Project administration, metadata curation, samples, logistics, sequencing, analysis, and software and analysis tools:**

Dr William L Hamilton PhD ^8, 10^.

**Leadership, supervision, samples logistics, project administration, funding acquisition sequencing and analysis:**

Dr David Bonsall PhD	^27^.

**Leadership and supervision, sequencing, analysis, funding acquisition, visualisation and software and analysis tools:**

Dr Ali R Awan	PhD ^74^.

**Leadership and supervision, funding acquisition, sequencing, analysis, metadata curation, samples and logistics:**

Dr Sally Corden PhD ^33^ .

**Leadership supervision, sequencing analysis, samples, logistics, and metadata curation:** Professor Ian Goodfellow PhD ^11^.

**Leadership, supervision, sequencing, analysis, samples, logistics, and Project administration:**

Professor Darren L Smith PhD ^60, 61^.

**Project administration, metadata curation, samples, logistics, sequencing and analysis:**
Dr Martin D Curran PhD ^14^, and Dr Surendra Parmar PhD ^14^**.**

**Samples, logistics, metadata curation, project administration sequencing and analysis:**
Dr James G Shepherd MBChB MRCP	^21^.

**Sequencing, analysis,	project administration, metadata curation and software and analysis tools:**

Dr Matthew D Parker PhD ^38^.

**Leadership, supervision, funding acquisition, samples, logistics, and metadata curation:**
Dr Catherine Moore^33^	.

**Leadership, supervision, metadata curation, samples, logistics, sequencing and analysis:**
Dr Derek J Fairley PhD ^6, 88^, Professor Matthew W Loose PhD ^54^, and Joanne Watkins MSc ^33^.

**Metadata curation, sequencing, analysis, leadership, supervision and software and analysis tools:**

Dr Matthew Bull PhD^33^ , and Dr Sam Nicholls PhD ^15^	.

**Leadership, supervision, visualisation, sequencing, analysis and software and analysis tools:**

Professor David M Aanensen PhD ^1, 30^.

**Sequencing, analysis, samples, logistics, metadata curation, and visualisation:**
Dr Sharon Glaysher ^70^	.

**Metadata curation, sequencing, analysis, visualisation, software and analysis tools:**
Dr Matthew Bashton PhD ^60^, and Dr Nicole Pacchiarini PhD ^33^.

**Sequencing, analysis, visualisation, metadata curation, and software and analysis tools**:
Dr Anthony P Underwood PhD ^1, 30^.

**Funding acquisition, leadership, supervision and project administration:**
Dr Thushan I de Silva PhD ^38^, and Dr Dennis Wang PhD ^38^**.**

**Project administration, samples, logistics, leadership and supervision**:

Dr Monique Andersson PhD ^28^ , Professor Anoop J Chauhan ^70^, Dr Mariateresa de Cesare PhD^26^, Dr Catherine Ludden ^1,3^ , and Dr Tabitha W Mahungu FRCPath ^91^.

**Sequencing, analysis, project administration and metadata curation:**
Dr Rebecca Dewar PhD ^20^, and Martin P McHugh MSc^20^.

**Samples, logistics, metadata curation and project administration:**
Dr Natasha G Jesudason MBChB MRCP FRCPath ^21^, Dr Kathy K Li MBBCh FRCPath	^21^, Dr Rajiv N Shah	BMBS MRCP MSc ^21^, and Dr Yusri Taha MD, PhD ^66^.

**Leadership, supervision, funding acquisition and metadata curation:**
Dr Kate E Templeton PhD ^20^**.**

**Leadership, supervision, funding acquisition, sequencing and analysis:**
Dr Simon Cottrell PhD ^33^, Dr Justin O’Grady PhD ^51^, Professor Andrew Rambaut DPhil ^19^, and Professor Colin P Smith PhD^93^.

**Leadership, supervision, metadata curation	, sequencing and analysis:**
Professor Matthew T.G. Holden PhD	^87^, and Professor Emma C Thomson PhD/FRCP ^21^.

**Leadership, supervision, samples, logistics and metadata curation**:
Dr Samuel Moses MD ^81, 82^.

**Sequencing, analysis, leadership, supervision, samples and logistics:**
Dr Meera Chand ^7^, Dr Chrystala Constantinidou PhD ^71^, Professor Alistair C Darby PhD ^46^, Professor Julian A Hiscox PhD ^46^, Professor Steve Paterson PhD ^46^, and Dr Meera Unnikrishnan PhD ^71^**.**

**Sequencing, analysis, leadership and supervision and software and analysis tools:**
Dr Andrew J Page PhD ^51^, and Dr Erik M Volz PhD^96^.

**Samples, logistics, sequencing, analysis and metadata curation:**
Dr Charlotte J Houldcroft PhD ^8^, Dr Aminu S Jahun PhD ^11^, Dr James P McKenna PhD ^88^, Dr Luke W Meredith PhD ^11^, Dr Andrew Nelson PhD ^61^, Sarojini Pandey MSc ^72^, and Dr Gregory R Young PhD ^60^.

**Sequencing, analysis, metadata curation, and software and analysis tools:**
Dr Anna Price PhD^34^, Dr Sara Rey PhD ^33^, Dr Sunando Roy PhD ^41^, Dr Ben Temperton Ph.D ^49^, and Matthew Wyles ^38^.

**Sequencing, analysis, metadata curation and visualisation:**

Stefan	Rooke	MSc^19^,	and Dr Sharif Shaaban PhD^87^.

**Visualisation, sequencing, analysis and software and analysis tools:**
Dr Helen Adams PhD ^35^, Dr Yann Bourgeois Ph.D ^69^, Dr Katie F Loveson Ph.D ^68^, Áine O'Toole MSc^19^, and Richard Stark MSc ^71^.

**Project administration, leadership and supervision:**

Dr Ewan M Harrison PhD ^1, 3^, David Heyburn ^33^, and Professor Sharon J Peacock ^2, 3^

**Project administration and funding acquisition:**

Dr David Buck PhD^26^ , and Michaela John BSc Hons ^36^

**Sequencing, analysis and project administration:**

Dorota	Jamrozy ^1^, and Dr Joshua Quick PhD ^15^

**Samples, logistics, and project administration:**

Dr Rahul Batra MD^78^, Katherine L Bellis BSc (Hons) ^1, 3^, Beth Blane BSc ^3^ , Sophia T Girgis MSc ^3^, Dr Angie Green PhD ^26^, Anita Justice MSc ^28^ , Dr Mark Kristiansen PhD ^41^ , and Dr Rachel J Williams PhD ^41^.

**Project administration, software and analysis tools:**

Radoslaw Poplawski BSc ^15^.

**Project administration and visualisation:**

Dr Garry P Scarlett Ph.D^69^.

**Leadership, supervision, and funding acquisition:**

Professor John A Todd PhD ^26^	, Dr Christophe Fraser	PhD ^27^,	Professor Judith Breuer MD ^40,41^, Professor Sergi Castellano PhD ^41^, Dr Stephen L Michell PhD ^49^, Professor Dimitris Gramatopoulos PhD, FRCPath^73^, and Dr Jonathan Edgeworth PhD, FRCPath ^78^.

**Leadership, supervision and metadata curation:**
Dr Gemma L Kay PhD ^51^.

**Leadership, supervision, sequencing and analysis:**
Dr Ana da Silva Filipe PhD ^21^ , Dr Aaron R Jeffries PhD ^49^, Dr Sascha Ott PhD ^71^, Professor Oliver Pybus ^24^ , Professor David L Robertson PhD ^21^, Dr David A Simpson PhD ^6^ , and Dr Chris Williams MB BS^33^.

**Samples, logistics, leadership and supervision:**

Dr Cressida Auckland FRCPath ^50^, Dr John Boyes MBChB^83^, Dr Samir Dervisevic FRCPath^52^ , Professor Sian Ellard FRCPath^49, 50^ , Dr Sonia Goncalves^1^, Dr Emma J Meader FRCPath ^51^, Dr Peter Muir PhD^2^, Dr Husam Osman PhD ^95^, Reenesh Prakash MPH^52^, Dr Venkat Sivaprakasam PhD^18^, and Dr Ian B Vipond PhD^2^.

**Leadership, supervision and visualisation**

Dr Jane AH Masoli MBChB ^49, 50^.

**Sequencing, analysis and metadata curation**

Dr Nabil-Fareed Alikhan PhD ^51^, Matthew Carlile BSc ^54^, Dr Noel Craine DPhil ^33^, Dr Sam T Haldenby PhD ^46^, Dr Nadine Holmes PhD ^54^, Professor Ronan A Lyons MD ^37^, Dr Christopher Moore PhD ^54^, Malorie Perry MSc ^33^ , Dr Ben Warne	MRCP^80^, and Dr Thomas Williams MD ^19^.

**Samples, logistics and metadata curation:**

Dr Lisa Berry PhD ^72^, Dr Andrew Bosworth PhD ^95^ ,Dr Julianne Rose Brown PhD^40^, Sharon Campbell MSc	^67^, Dr Anna Casey PhD	^17^, Dr Gemma Clark PhD ^56^, Jennifer Collins BSc ^66^, Dr Alison Cox PhD ^43,^ ^44^ , Thomas Davis MSc ^84^, Gary Eltringham BSc ^66^, Dr Cariad Evans ^38, 39^ , Dr Clive Graham MD ^64^, Dr Fenella Halstead PhD ^18^, Dr Kathryn Ann Harris PhD ^40^, Dr Christopher Holmes PhD ^58^, Stephanie Hutchings ^2^ , Professor Miren Iturriza-Gomara PhD ^46^ , Dr Kate Johnson ^38, 39^, Katie Jones MSc ^72^, Dr Alexander J Keeley MRCP ^38^, Dr Bridget A Knight PhD ^49, 50^ , Cherian Koshy MSc, CSci, FIBMS ^90^, Steven Liggett ^63^ , Hannah Lowe MSc ^81^ , Dr Anita O Lucaci PhD ^46^ , Dr Jessica Lynch PhD MBChB ^25, 29^ , Dr Patrick C McClure PhD ^55^ , Dr Nathan Moore MBChB ^31^ , Matilde Mori BSc ^25, 29, 32^ , Dr David G Partridge FRCP, FRCPath ^38, 39^ , Pinglawathee Madona ^43, 44^ , Hannah M Pymont MSc ^2^ , Dr Paul Anthony Randell MBBCh ^43, 44^ , Dr Mohammad Raza ^38, 39^ , Felicity Ryan MSc ^81^ , Dr Robert Shaw FRCPath ^28^, Dr Tim J Sloan PhD ^57^ , and Emma Swindells BSc ^65^ .

**Sequencing, analysis, Samples and logistics:**

Alexander Adams BSc	^33^, Dr Hibo Asad PhD ^33^, Alec Birchley MSc ^33^ , Tony Thomas Brooks BSc (Hons) ^41^, Dr Giselda Bucca PhD ^93^, Ethan Butcher ^70^, Dr Sarah L Caddy PhD ^13^, Dr Laura G Caller PhD ^2, 3, 12^ , Yasmin Chaudhry BSc ^11^, Jason Coombes BSc (HONS) ^33^, Michelle Cronin ^33^, Patricia L Dyal MPhil ^41^, Johnathan M Evans MSc ^33^,Laia Fina ^33^, Bree Gatica-Wilcox MPhil ^33^, Dr Iliana Georgana PhD ^11^, Lauren Gilbert A-Levels ^33^ , Lee Graham BSc ^33^, Danielle C Groves BA ^38^, Grant Hall BSc ^11^, Ember Hilvers MPH	^33^ , Dr Myra Hosmillo PhD ^11^, Hannah Jones ^33^, Sophie Jones MSc ^33^, Fahad	A Khokhar BSc	^13^ , Sara Kumziene-Summerhayes MSc ^33^, George MacIntyre-Cockett BSc ^26^, Dr Rocio T Martinez Nunez PhD ^94^ , Dr Caoimhe McKerr PhD ^33^ , Dr Claire McMurray PhD ^15^,	Dr Richard Myers ^7^, Yasmin Nicole Panchbhaya BSc ^41^ , Malte L Pinckert MPhil ^11^ , Amy Plimmer ^33^ , Dr Joanne Stockton PhD ^15^ , Sarah Taylor ^33^ , Dr Alicia Thornton ^7^ , Amy Trebes MSc ^26^ , Alexander J Trotter MRes ^51^ ,Helena Jane Tutill BSc ^41^ ,Charlotte A Williams BSc ^41^ , Anna Yakovleva BSc ^11^ and Dr Wen C Yew PhD ^62^.

**Sequencing, analysis and software and analysis tools:**
Dr Mohammad T Alam PhD ^71^	, Dr Laura Baxter PhD ^71^, Olivia Boyd MSc ^96^ , Dr Fabricia F. Nascimento PhD ^96^, Timothy M Freeman MPhil ^38^, Lily Geidelberg MSc ^96^, Dr Joseph Hughes PhD ^21^, David Jorgensen MSc ^96^, Dr Benjamin B Lindsey MRCP ^38^, Dr Richard J Orton PhD ^21^ , Dr Manon Ragonnet-Cronin PhD ^96^ Joel Southgate MSc ^33, 34,^ and Dr Sreenu Vattipally PhD ^21^.

**Samples, logistics and software and analysis tools:**

Dr Igor	Starinskij MSc MRCP ^23^.

**Visualisation and software and analysis tools:**
Dr Joshua B Singer PhD ^21^ , Dr Khalil Abudahab PhD ^1, 30^, Leonardo de Oliveira Martins PhD^51^ , Dr Thanh Le-Viet PhD	^51^ ,Mirko Menegazzo ^30^ ,Ben EW Taylor Meng ^1, 30^, and Dr Corin A Yeats PhD ^30^.

**Project Administration:**

Sophie Palmer	^3^, Carol M Churcher ^3^ , Dr Alisha Davies ^33^, Elen De Lacy MSc ^33^, Fatima Downing ^33^, Sue Edwards ^33^ , Dr Nikki Smith	PhD ^38^ , and Dr Frances Bolt PhD ^44, 45^	.

**Leadership and supervision:**

Dr. Alex Alderton ^1^, Dr Matt Berriman ^1^, Ian G Charles ^51^, Dr Nicholas Cortes MBChB ^31^ ,Dr Tanya Curran PhD ^88^ , Prof John Danesh ^1^, Dr Sahar Eldirdiri MBBS, MSC FRCPath ^84^, Dr Ngozi Elumogo FRCPath ^52^, Prof Andrew Hattersley FRS ^49, 50^, Professor Alison Holmes MD ^44, 45^, Dr Robin Howe ^33^, Dr Rachel Jones ^33^ , Anita Kenyon MSc ^84^, Prof Robert A Kingsley PhD ^51^, Professor Dominic Kwiatkowski ^1, 9^, Dr Cordelia Langford^1^, Dr Jenifer Mason MBBS ^48^, Dr Alison E Mather PhD ^51^, Lizzie Meadows MA ^51^, Dr Sian Morgan FRCPath ^36^, Dr James Price PhD ^44, 45^, Trevor I Robinson MSc ^48^ , Dr Giri Shankar ^33^ , John Wain ^51^, and Dr Mark A Webber PhD^51^	.

**Metadata curation:**

Dr Declan T Bradley PhD ^5, 6^ ,Dr Michael R Chapman PhD ^1, 3, 4^ , Dr Derrick Crooke ^28^ , Dr David Eyre PhD ^28^, Professor Martyn Guest PhD^34^ , Huw Gulliver ^34^ , Dr Sarah Hoosdally ^28^ , Dr Christine Kitchen PhD ^34^ , Dr Ian Merrick PhD ^34^, Siddharth Mookerjee MPH ^44, 45^ , Robert Munn	BSc ^34^ , Professor Timothy Peto PhD^28^, Will Potter ^52^, Dr Dheeraj K Sethi MBBS ^52^, Wendy Smith ^56^ ,Dr Luke B Snell MB BS ^75, 94^ , Dr Rachael Stanley PhD ^52^ , Claire Stuart ^52^ and Dr Elizabeth Wastenge MD ^20^.

**Sequencing and analysis:**

Dr Erwan Acheson PhD ^6^ , Safiah Afifi BSc ^36^ , Dr Elias Allara	MD PhD ^2, 3^ , Dr Roberto Amato ^1^, Dr Adrienn Angyal PhD^38^, Dr Elihu Aranday-Cortes PhD/DVM^21^ , Cristina Ariani ^1^, Jordan Ashworth ^19^,	Dr Stephen Attwood ^24^, Alp Aydin MSci ^51^ , David J Baker BEng ^51^ , Dr Carlos E Balcazar PhD ^19^, Angela Beckett MSc	^68^ Robert Beer BSc ^36^ ,	Dr Gilberto Betancor	PhD^76^, Emma	Betteridge ^1^ , Dr David Bibby ^7^ , Dr Daniel Bradshaw	^7^ , Catherine Bresner Bsc(Hons) ^34^, Dr Hannah E Bridgewater PhD ^71^ , Alice Broos BSc (Hons) ^21^ , Dr Rebecca Brown PhD ^38^ , Dr Paul E Brown PhD ^71^, Dr Kirstyn Brunker PhD ^22^ , Dr Stephen N Carmichael PhD ^21^ , Jeffrey K. J. Cheng MSc ^71^, Dr Rachel Colquhoun DPhil ^19^ , Dr Gavin Dabrera ^7^ , Dr Johnny Debebe PhD ^54^, Eleanor Drury ^1^, Dr Louis du Plessis ^24^ , Richard Eccles MSc ^46^, Dr Nicholas Ellaby ^7^, Audrey Farbos MSc ^49^, Ben Farr ^1^ ,Dr Jacqueline Findlay PhD ^41^ , Chloe L Fisher MSc ^74^, Leysa Marie Forrest MSc ^41^, Dr Sarah Francois ^24^, Lucy R. Frost BSc ^71^, William Fuller BSc ^34^ , Dr Eileen Gallagher ^7^ , Dr Michael D Gallagher PhD^19^ , Matthew Gemmell MSc ^46^, Dr Rachel AJ Gilroy PhD ^51^, Scott Goodwin ^1^, Dr Luke R Green PhD ^38^ ,Dr Richard Gregory PhD ^46^ ,Dr Natalie Groves ^7^ ,Dr James W Harrison PhD ^49^, Hassan Hartman ^7^ , Dr Andrew R Hesketh PhD^93^,Verity Hill ^19^, Dr Jonathan Hubb ^7^ ,Dr Margaret Hughes PhD^46^ ,Dr David K Jackson ^1^ ,Dr Ben Jackson PhD ^19^ ,Dr Keith James ^1^ ,Natasha Johnson BSc (Hons)^21^ ,Ian Johnston ^1^, Jon-Paul Keatley ^1^, Dr Moritz Kraemer ^24^, Dr Angie Lackenby ^7^, Dr Mara Lawniczak ^1^ , Dr David Lee ^7^, Rich Livett ^1^, Stephanie Lo ^1^, Daniel Mair BSc (Hons) ^21^, Joshua Maksimovic FD sport science ^36^, Nikos Manesis ^7^ ,Dr Robin Manley Ph.D ^49^, Dr Carmen Manso ^7^ ,Dr Angela Marchbank BSc ^34^ ,Dr Inigo Martincorena ^1^ ,Dr Tamyo Mbisa ^7^, Kathryn McCluggage MSC ^36^,Dr JT McCrone PhD ^19^, Shahjahan Miah ^7^ , Michelle L Michelsen BSc ^49^ , Dr Mari Morgan PhD ^33^, Dr Gaia Nebbia PhD, FRCPath ^78^,Charlotte Nelson MSc ^46^ ,Jenna Nichols BSc (Hons) ^21^ ,Dr Paola Niola PhD ^41^ ,Dr Kyriaki Nomikou PhD^21^ ,Steve Palmer ^1^ , Dr. Naomi Park ^1^, Dr Yasmin A Parr PhD^21^ ,Dr Paul J Parsons PhD ^38^ , Vineet Patel ^7^ ,Dr. Minal Patel ^1^ ,Clare Pearson MSc ^2, 1^ ,Dr Steven Platt ^7^ ,Christoph Puethe ^1^, Dr. Mike Quail ^1^,Dr JaynaRaghwani ^24^ , Dr Lucille Rainbow PhD ^46^ ,Shavanthi Rajatileka ^1^ ,Dr Mary Ramsay ^7^ ,Dr Paola C Resende Silva PhD ^41, 42^, Steven Rudder 51, Dr Chris Ruis ^3^ ,Dr Christine M Sambles PhD ^49^ ,Dr Fei Sang PhD ^54^ ,Dr Ulf Schaefer^7^ ,Dr Emily Scher PhD ^19^ ,Dr. Carol Scott ^1^ ,Lesley Shirley ^1^, Adrian W Signell BSc ^76^ ,John Sillitoe ^1^ ,Christen Smith ^1^ ,Dr Katherine L Smollett PhD ^21^ ,Karla Spellman FD ^36^ ,Thomas D Stanton BSc ^19^ ,Dr David J Studholme PhD ^49^ ,Ms Grace Taylor-Joyce BSc ^71^ ,Dr Ana P Tedim PhD ^51^ ,Dr Thomas Thompson PhD^6^ ,Dr Nicholas M Thomson PhD ^51^ ,Scott Thurston^1^ ,Lily Tong PhD ^21^ ,Gerry Tonkin-Hill ^1^, Rachel M Tucker MSc ^38^ , Dr Edith E Vamos PhD ^4^,Dr Tetyana Vasylyeva^24^ , Joanna Warwick-Dugdale BSc ^49^ , Danni Weldon ^1^ ,Dr Mark Whitehead PhD ^46^ ,Dr David Williams ^7^,Dr Kathleen A Williamson PhD^19^,Harry D Wilson BSc ^76^,Trudy Workman HNC ^34^ ,Dr Muhammad Yasir PhD ^51^, Dr Xiaoyu Yu PhD ^19^, and Dr Alex Zarebski ^24^.

**Samples and logistics:**

Dr Evelien M Adriaenssens PhD ^51^, Dr Shazaad S Y Ahmad MSc ^2, 47^ , Adela Alcolea-Medina MPharm ^59, 77^ ,Dr John Allan PhD^60^, Dr Patawee Asamaphan PhD^21^, Laura Atkinson MSc ^40^, Paul Baker MD ^63^, Professor Jonathan Ball PhD ^55^, Dr Edward Barton MD^64^, Dr. Mathew A Beale^1^, Dr. Charlotte Beaver^1^ , Dr Andrew Beggs PhD^16^, Dr Andrew Bell PhD^51^, Duncan J Berger ^1^, Dr Louise Berry. ^56^, Claire M Bewshea MSc ^49^, Kelly Bicknell ^70^, Paul Bird ^58^, Dr Chloe Bishop ^7^ , Dr Tim Boswell ^56^, Cassie Breen BSc^48^, Dr Sarah K Buddenborg^1^, Dr Shirelle Burton-Fanning MD ^66,^ Dr Vicki Chalker ^7^, Dr Joseph G Chappell PhD ^55^, Themoula Charalampous MSc ^78, 94^, Claire Cormie^3^, Dr Nick Cortes PhD^29, 25^, Dr Lindsay J Coupland PhD ^52^, Angela Cowell MSc^48^ , Dr Rose K Davidson PhD ^53 ,^ Joana Dias MSc^3^ , Dr Maria Diaz PhD^51^ , Thomas Dibling^1^, Matthew J Dorman^1^, Dr Nichola Duckworth^57^, Scott Elliott^70^, Sarah Essex^63^, Karlie Fallon ^58^ , Theresa Feltwell ^8^ , Dr Vicki M Fleming PhD ^56^, Sally Forrest BSc ^3^, Luke Foulser^1^, Maria V Garcia-Casado^1^, Dr Artemis Gavriil PhD ^41^, Dr Ryan P George PhD^47^, Laura Gifford MSc ^33^, Harmeet K Gill PhD^3^, Jane Greenaway MSc^65^, Luke Griffith Bsc^53^, Ana Victoria Gutierrez^51^, Dr Antony D Hale MBBS^85^, Dr Tanzina Haque FRCPath, PhD^91^, Katherine L Harper MBiol^85^, Dr Ian Harrison ^7^ , Dr Judith Heaney PhD^89^, Thomas Helmer ^58^, Ellen E Higginson PhD ^3^ , Richard Hopes ^2^, Dr Hannah C Howson-Wells PhD ^56^, Dr Adam D Hunter ^1^, Robert Impey ^70^, Dr Dianne Irish-Tavares FRCPath ^91^, David A Jackson^1^ , Kathryn A Jackson MSc ^46^ , Dr Amelia Joseph ^56^, Leanne Kane ^1^, Sally Kay ^1^, Leanne M Kermack MSc ^3^, Manjinder Khakh ^56^, Dr Stephen P Kidd PhD^29, 25,31^, , Dr Anastasia Kolyva PhD ^51^, Jack CD Lee BSc ^40^, Laura Letchford ^1^ , Nick Levene MSc^79^, Dr LisaJ Levett PhD ^89^, Dr Michelle M Lister PhD ^56^, Allyson Lloyd ^70^ , Dr Joshua Loh PhD^60^ , Dr Louissa R Macfarlane-Smith PhD^85^, Dr Nicholas W Machin MSc ^2 , 47^, Mailis Maes M.phil^3^, Dr Samantha McGuigan ^1^, Liz McMinn ^1^, Dr Lamia Mestek-Boukhibar D.Phil ^41^, Dr Zoltan Molnar PhD ^6^, Lynn Monaghan ^79^, Dr Catrin Moore ^27^, Plamena Naydenova BSc ^3^, Alexandra S Neaverson ^1^, Dr. Rachel Nelson PhD ^1^, Marc O Niebel MSc^21^ , Elaine O'Toole BSc ^48^ , Debra Padgett BSc ^64^, Gaurang Patel ^1^ , Dr Brendan AI Payne MD ^66^, Liam Prestwood ^1^, Dr Veena Raviprakash MD^67^, Nicola Reynolds PhD^86^ Dr Alex Richter PhD ^16^, Dr Esther Robinson PhD^95^, Dr Hazel A Rogers^1^, Dr Aileen Rowan PhD ^96^, Garren Scott BSc ^64^, Dr Divya Shah PhD^40^, Nicola Sheriff BSc ^67^, Dr Graciela Sluga MD - MSc^92^ , Emily Souster^1^, Dr. Michael Spencer-Chapman^1^, Sushmita Sridhar BSc^1, 3^, Tracey Swingler ^53^, Dr Julian Tang^58^, Professor Graham P Taylor DSc^96^, Dr Theocharis Tsoleridis PhD^55^, Dr Lance Turtle PhD MRCP^46^, Dr Sarah Walsh ^57^, Dr Michelle Wantoch PhD ^86^, Joanne Watts BSc^48^ , Dr Sheila Waugh MD^66^, Sam Weeks^41^, Dr Rebecca Williams BMBS ^31^, Dr Iona Willingham^56^, Dr Emma L Wise PhD ^25, 29, 31^, Victoria Wright BSc ^54^, Dr Sarah Wyllie ^70^ , and Jamie Young BSc ^3^.

**Software and analysis tools**

Amy Gaskin MSc^33^, Dr Will Rowe PhD ^15^, and Dr Igor Siveroni PhD^96^.

**Visualisation:**

Dr Robert Johnson PhD ^96^.

**1** Wellcome Sanger Institute, **2** Public Health England, **3** University of Cambridge, **4** Health Data Research UK, Cambridge, **5** Public Health Agency, Northern Ireland ,**6** Queen's University Belfast **7** Public Health England Colindale, **8** Department of Medicine, University of Cambridge, **9** University of Oxford, **10** Departments of Infectious Diseases and Microbiology, Cambridge University Hospitals NHS Foundation Trust; Cambridge, UK, **11** Division of Virology, Department of Pathology, University of Cambridge, **12** The Francis Crick Institute, **13** Cambridge Institute for Therapeutic Immunology and Infectious Disease, Department of Medicine, **14** Public Health England, Clinical Microbiology and Public Health Laboratory, Cambridge, UK, **15** Institute of Microbiology and Infection, University of Birmingham, **16** University of Birmingham, **17** Queen Elizabeth Hospital, **18** Heartlands Hospital, **19** University of Edinburgh, **20** NHS Lothian, **21** MRC-University of Glasgow Centre for Virus Research, **22** Institute of Biodiversity, Animal Health & Comparative Medicine, University of Glasgow, **23** West of Scotland Specialist Virology Centre, **24** Dept Zoology, University of Oxford, **25** University of Surrey, **26** Wellcome Centre for Human Genetics, Nuffield Department of Medicine, University of Oxford, **27** Big Data Institute, Nuffield Department of Medicine, University of Oxford, **28** Oxford University Hospitals NHS Foundation Trust, **29** Basingstoke Hospital, **30** Centre for Genomic Pathogen Surveillance, University of Oxford, **31** Hampshire Hospitals NHS Foundation Trust, **32** University of Southampton, **33** Public Health Wales NHS Trust, **34** Cardiff University, **35** Betsi Cadwaladr University Health Board, **36** Cardiff and Vale University Health Board, **37** Swansea University, **38** University of Sheffield, **39** Sheffield Teaching Hospitals, **40** Great Ormond Street NHS Foundation Trust, **41** University College London, **42** Oswaldo Cruz Institute, Rio de Janeiro **43** North West London Pathology, **44** Imperial College Healthcare NHS Trust, **45** NIHR Health Protection Research Unit in HCAI and AMR, Imperial College London, **46** University of Liverpool, **47** Manchester University NHS Foundation Trust, **48** Liverpool Clinical Laboratories, **49** University of Exeter, **50** Royal Devon and Exeter NHS Foundation Trust, **51** Quadram Institute Bioscience, University of East Anglia, **52** Norfolk and Norwich University Hospital, **53** University of East Anglia, **54** Deep Seq, School of Life Sciences, Queens Medical Centre, University of Nottingham, **55** Virology, School of Life Sciences, Queens Medical Centre, University of Nottingham, **56** Clinical Microbiology Department, Queens Medical Centre, **57** PathLinks, Northern Lincolnshire & Goole NHS Foundation Trust, **58** Clinical Microbiology, University Hospitals of Leicester NHS Trust, **59** Viapath, **60** Hub for Biotechnology in the Built Environment, Northumbria University, **61** NU-OMICS Northumbria University, **62** Northumbria University, **63** South Tees Hospitals NHS Foundation Trust, **64** North Cumbria Integrated Care NHS Foundation Trust, **65** North Tees and Hartlepool NHS Foundation Trust, **66** Newcastle Hospitals NHS Foundation Trust, **67** County Durham and Darlington NHS Foundation Trust, **68** Centre for Enzyme Innovation, University of Portsmouth, **69** School of Biological Sciences, University of Portsmouth, **70** Portsmouth Hospitals NHS Trust, **71** University of Warwick, **72** University Hospitals Coventry and Warwickshire, **73** Warwick Medical School and Institute of Precision Diagnostics, Pathology, UHCW NHS Trust, **74** Genomics Innovation Unit, Guy's and St. Thomas' NHS Foundation Trust, **75** Centre for Clinical Infection & Diagnostics Research, St. Thomas' Hospital and Kings College London, **76** Department of Infectious Diseases, King's College London, **77** Guy's and St. Thomas’ Hospitals NHS Foundation Trust, **78** Centre for Clinical Infection and Diagnostics Research, Department of Infectious Diseases, Guy's and St Thomas' NHS Foundation Trust, **79** Princess Alexandra Hospital Microbiology Dept. , **80** Cambridge University Hospitals NHS Foundation Trust, **81** East Kent Hospitals University NHS Foundation Trust, **82** University of Kent, **83** Gloucestershire Hospitals NHS Foundation Trust, **84** Department of Microbiology, Kettering General Hospital, **85** National Infection Service, PHE and Leeds Teaching Hospitals Trust, **86** Cambridge Stem Cell Institute, University of Cambridge, **87** Public Health Scotland, 88 Belfast Health & Social Care Trust, **89** Health Services Laboratories, **90** Barking, Havering and Redbridge University Hospitals NHS Trust, **91** Royal Free NHS Trust, **92** Maidstone and Tunbridge Wells NHS Trust, **93** University of Brighton, **94** Kings College London, **95** PHE Heartlands, **96** Imperial College London.
