## Supplementary figures and images for "Genomic epidemiology of COVID-19 in care homes in the East of England"

### Fig 1

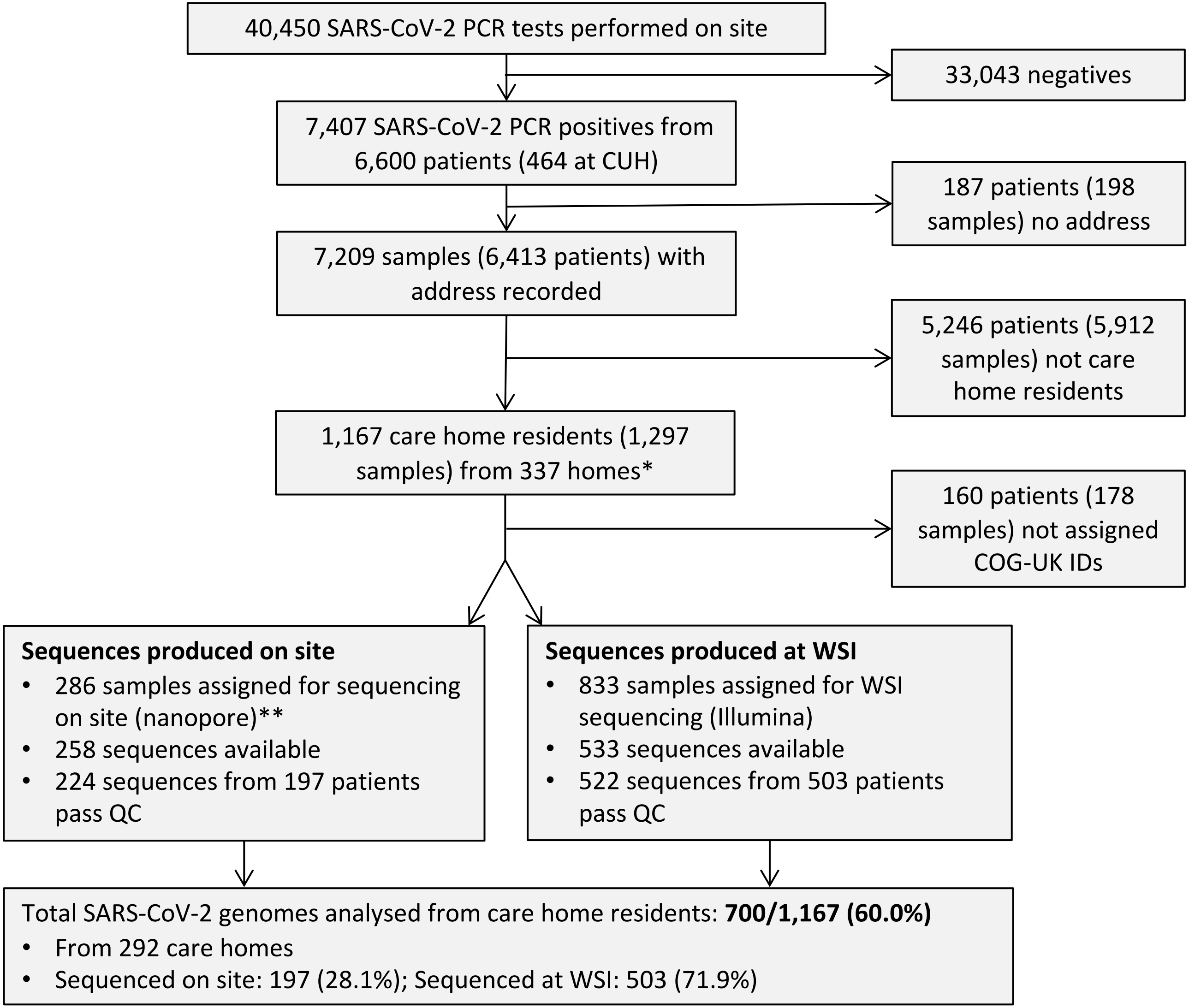

### Fig 1 Supp 2

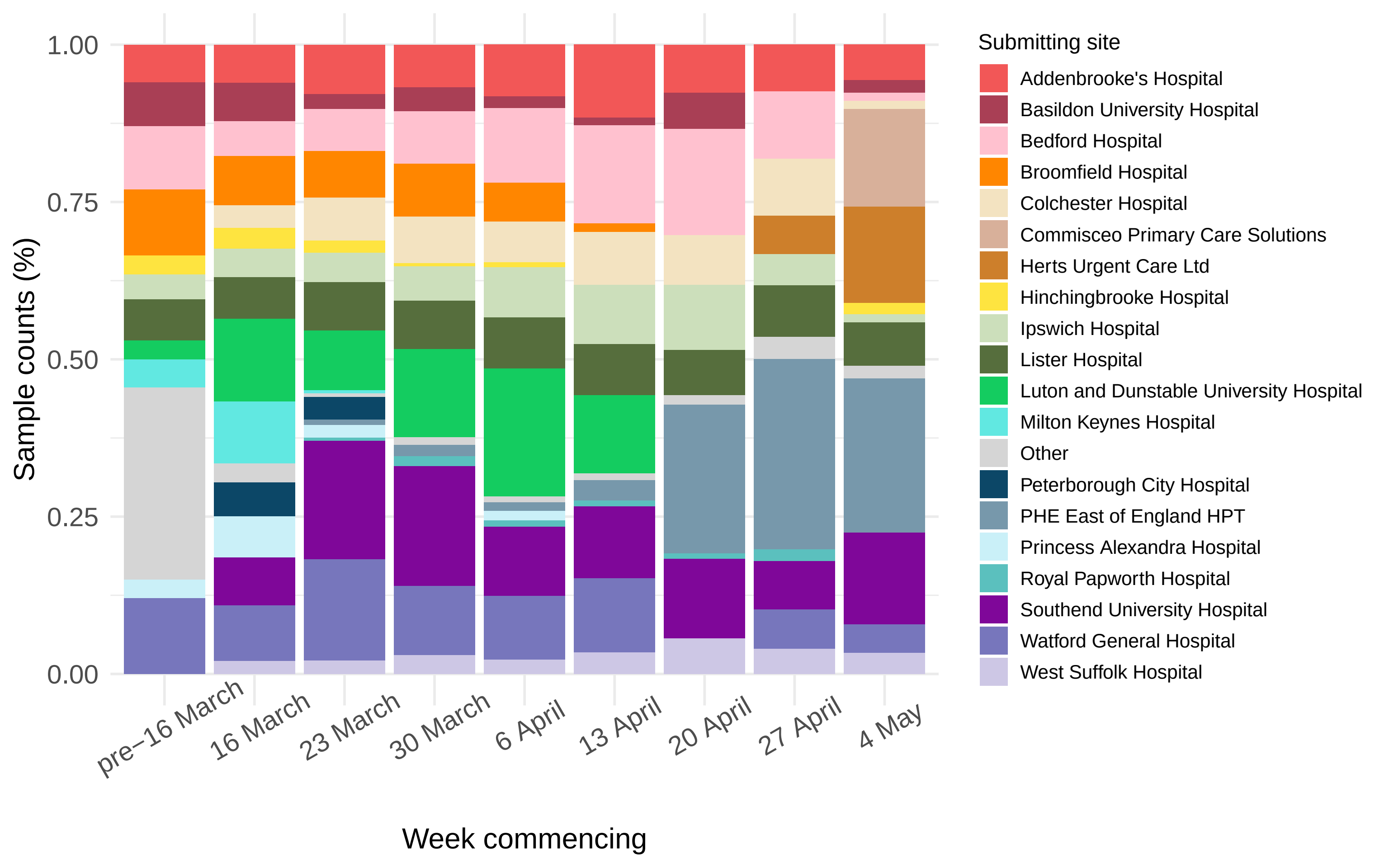

### Fig 1 Supp 3

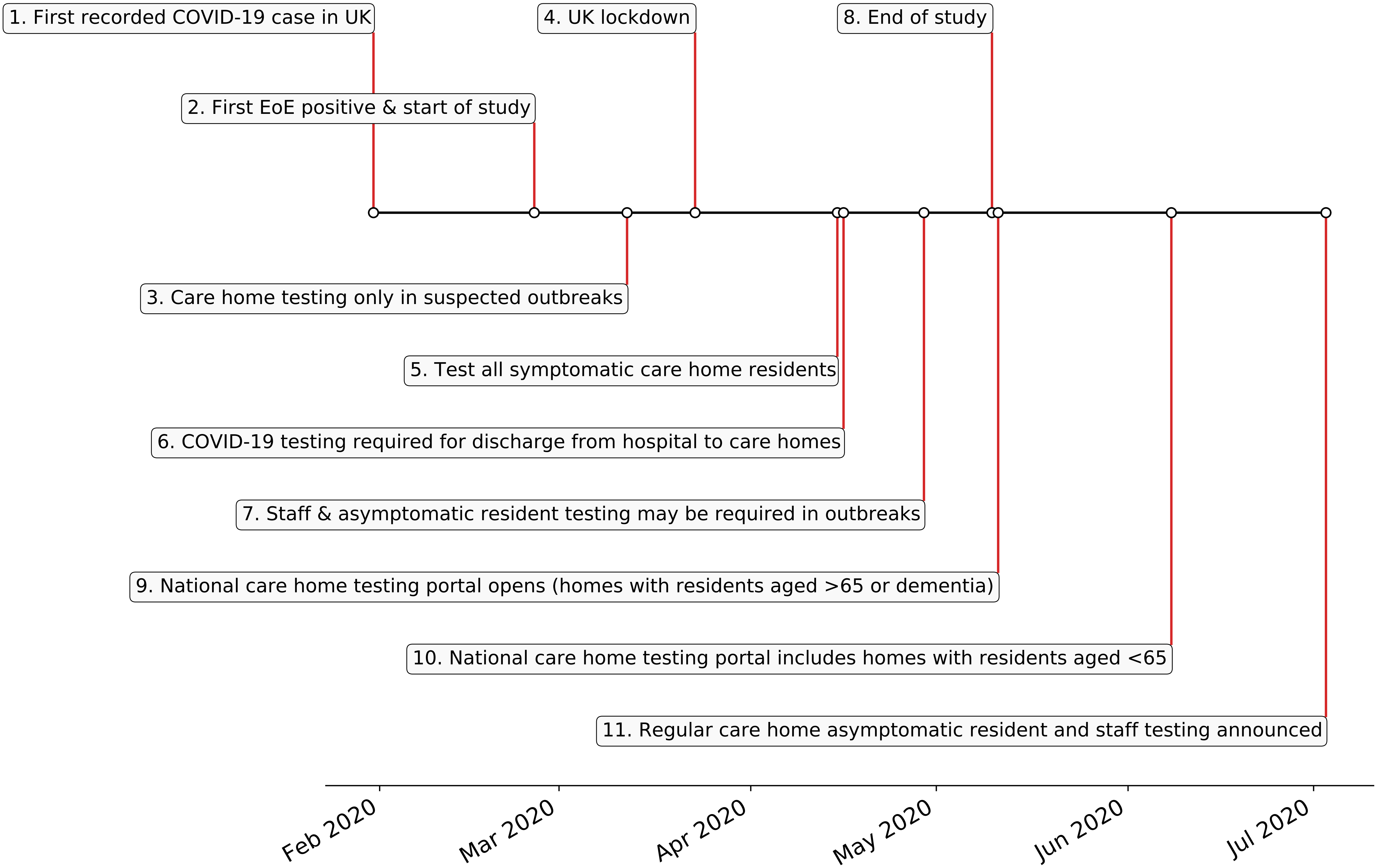

### Fig 1 suppl 1

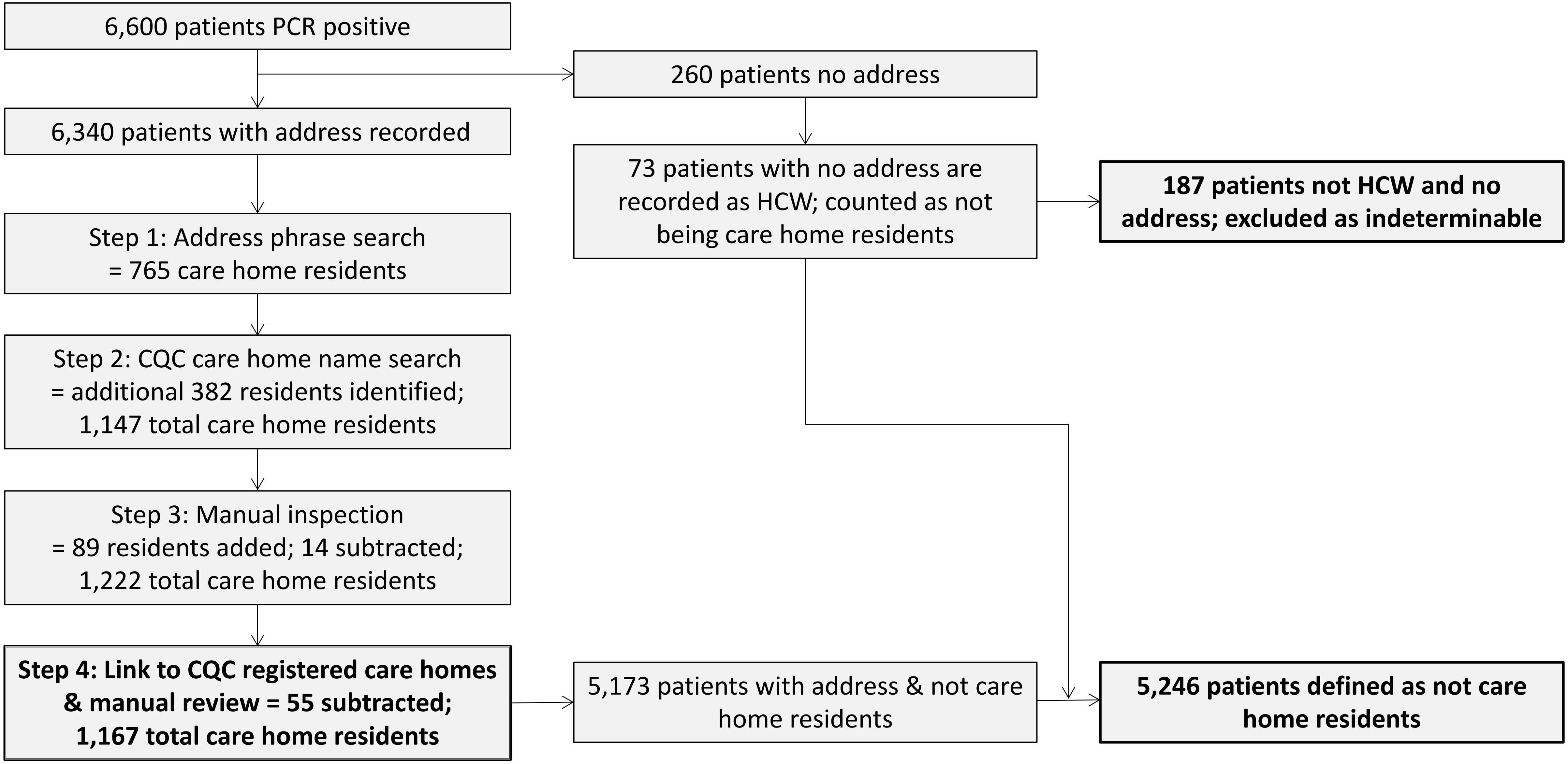

### Fig 2

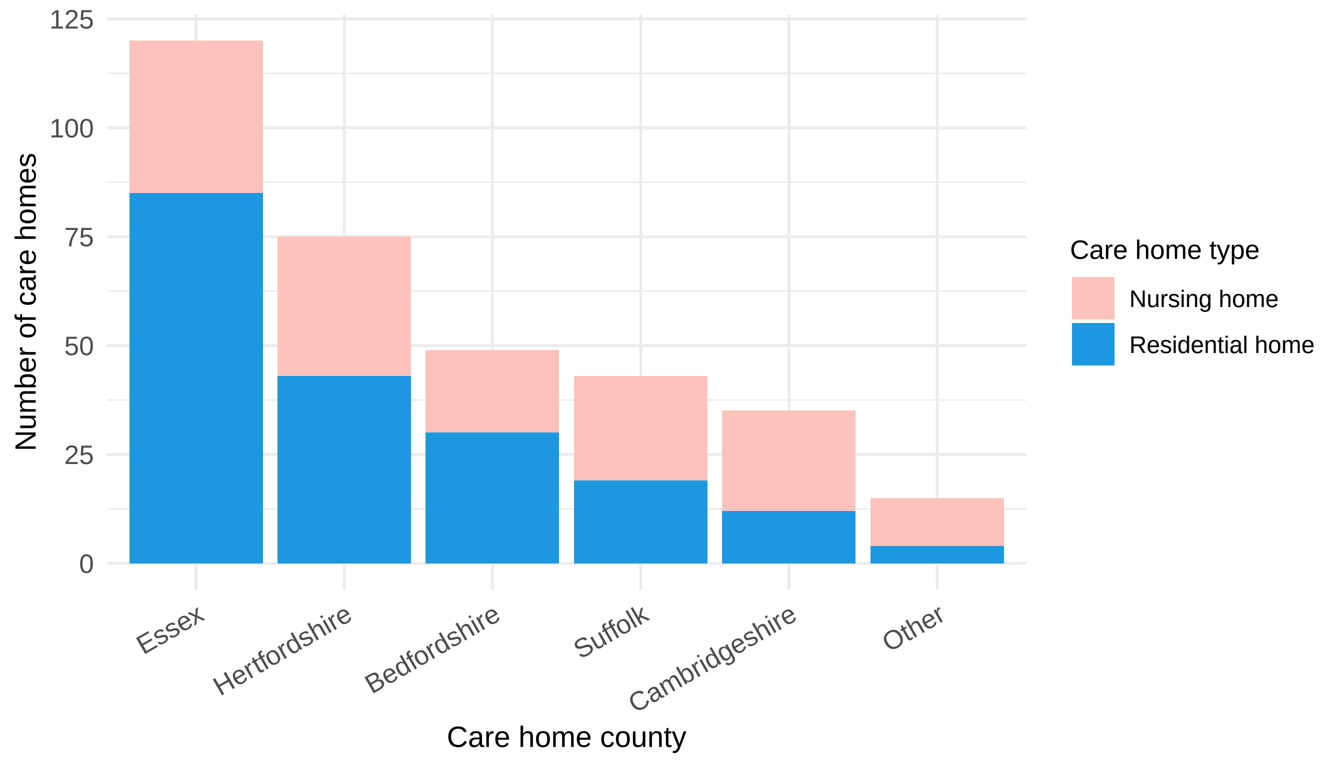

### Fig 2 Supp 1

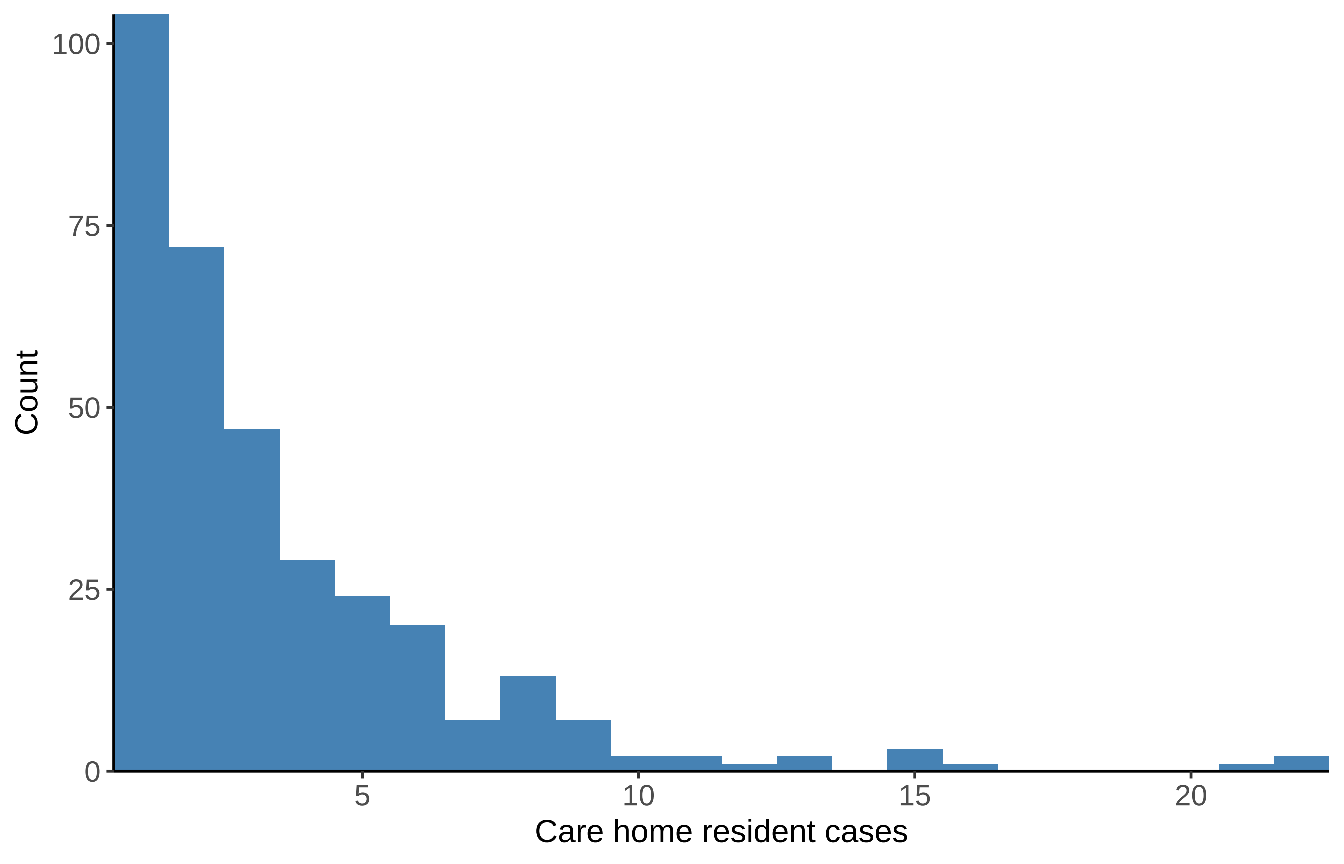

### Fig 3

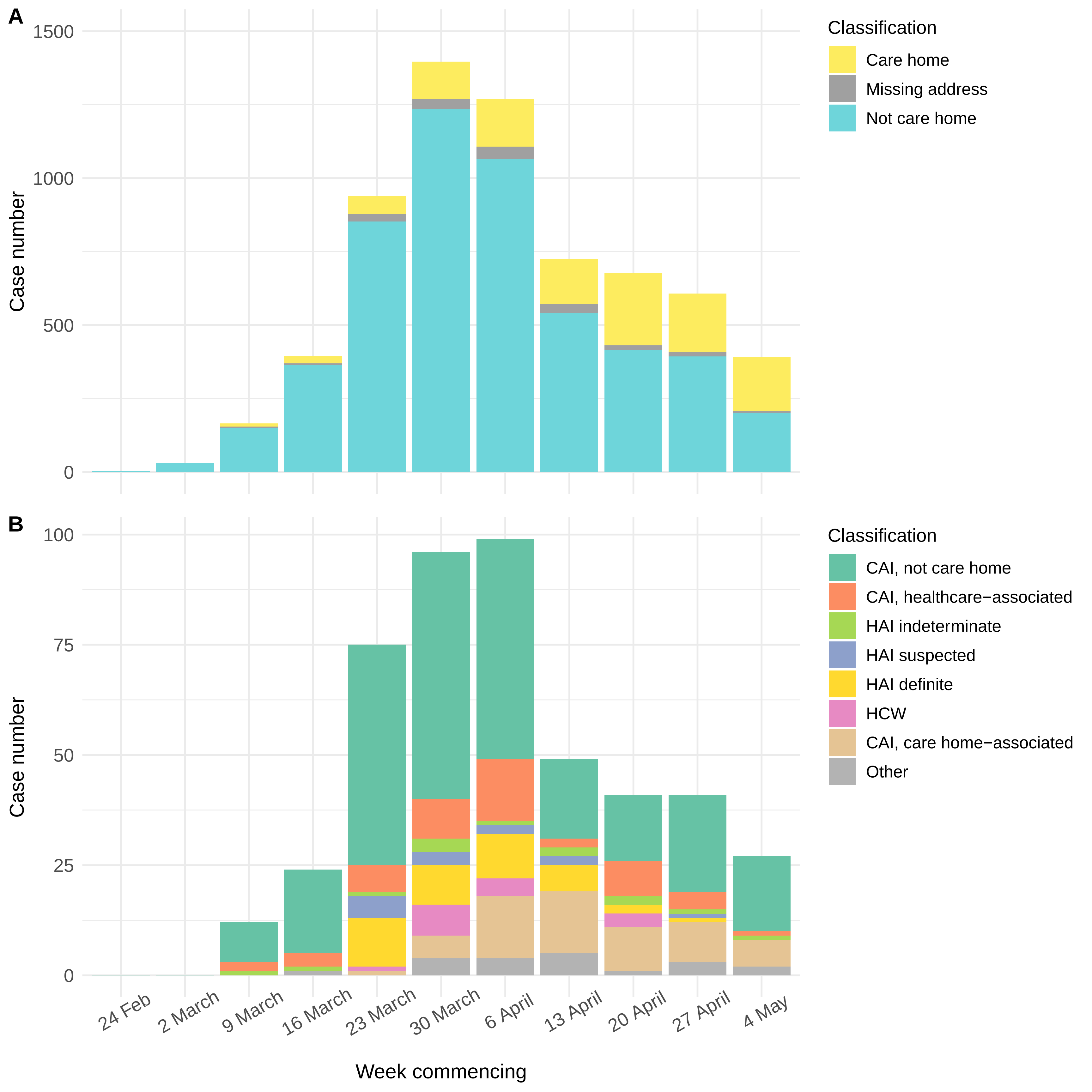

### Fig 3 Supp 1

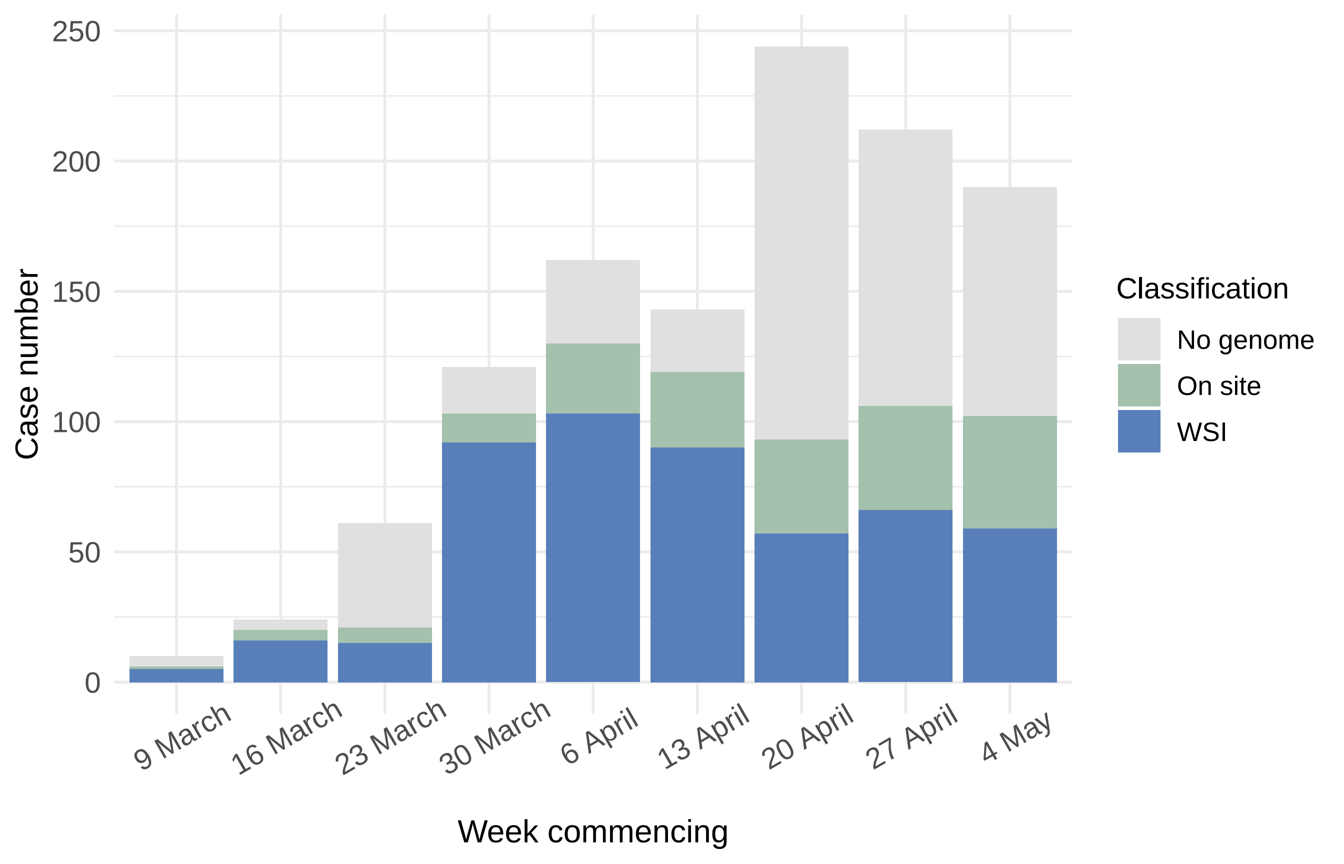

### Fig 4

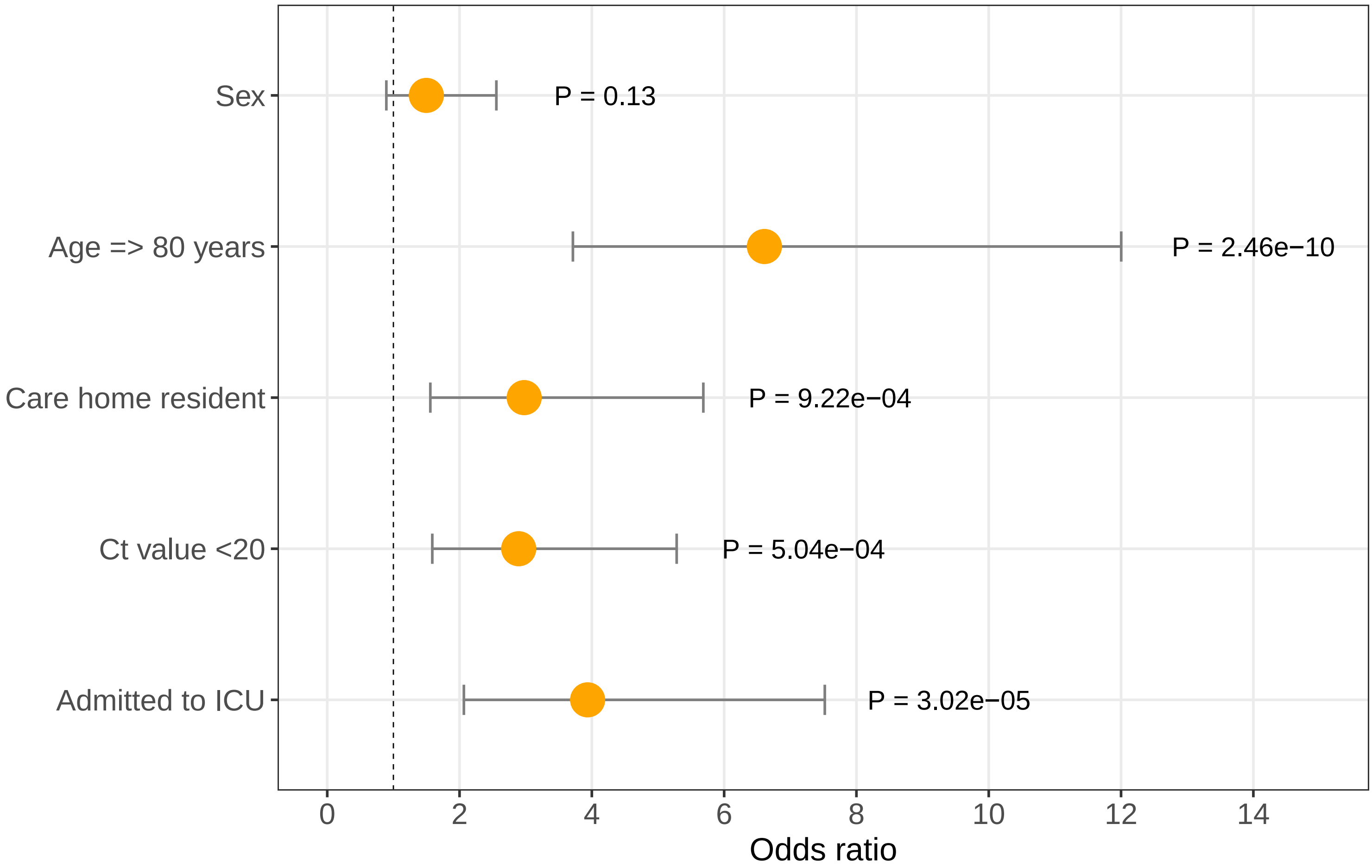

### Fig 4 Supp 1

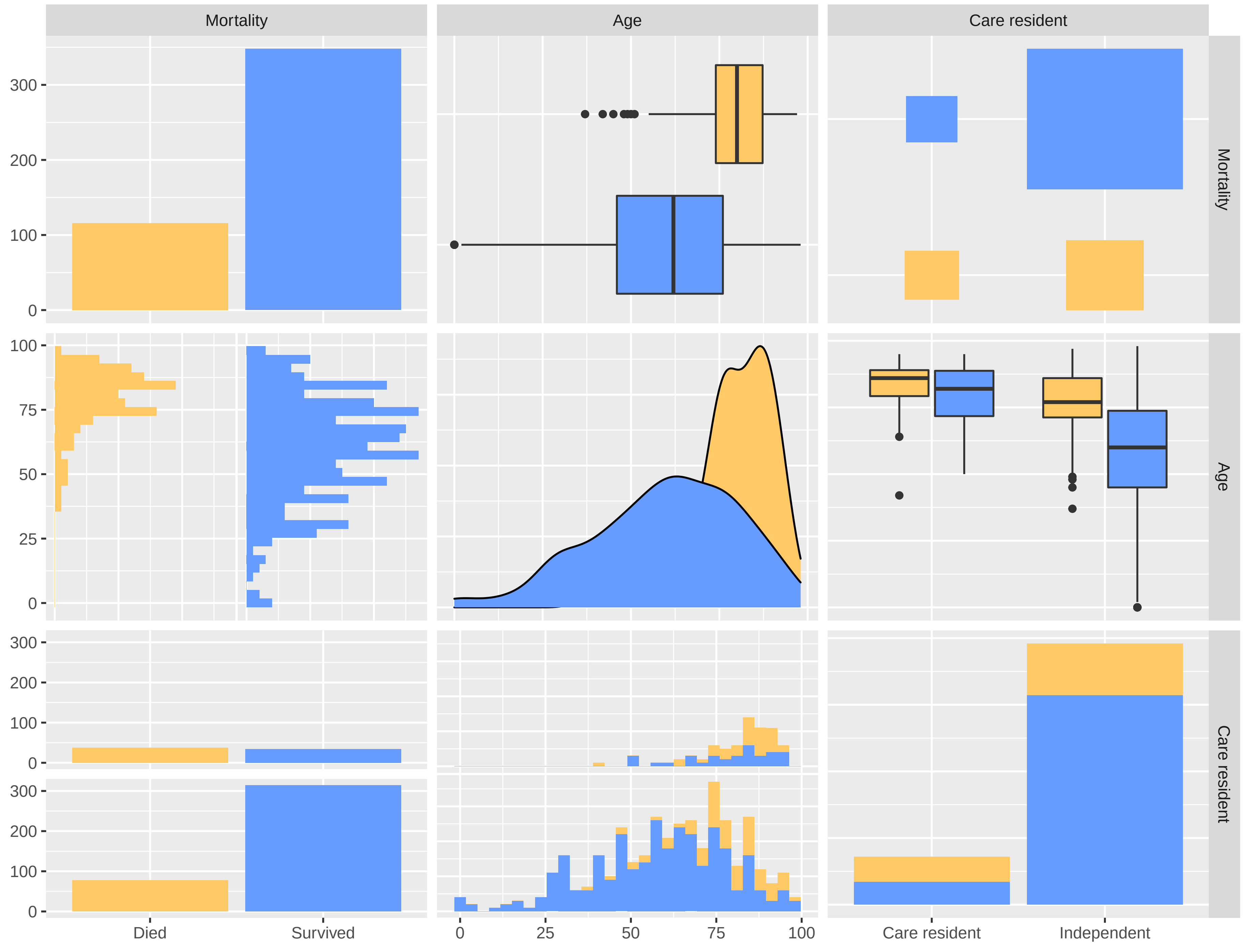

### Fig 5

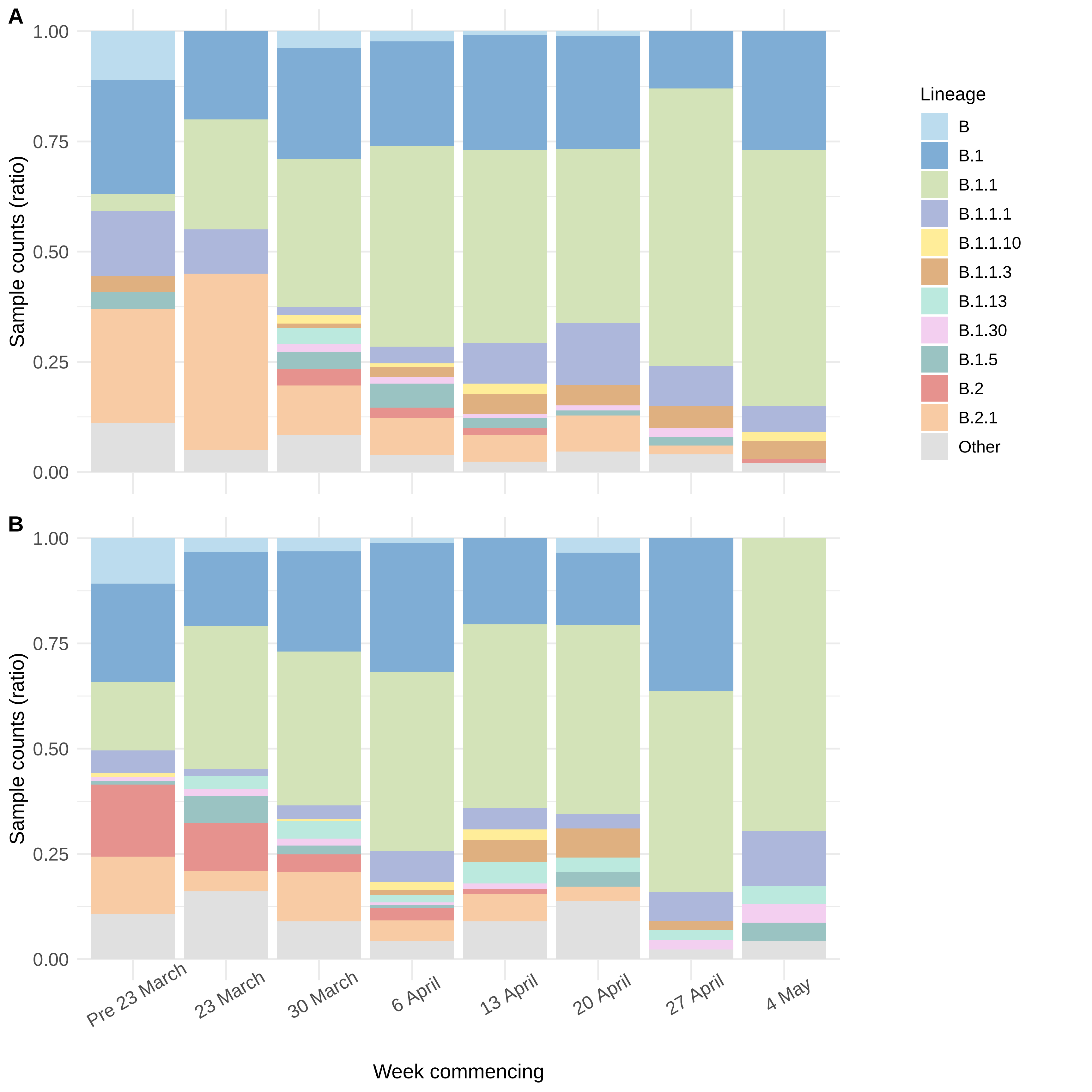

### Fig 5 Supp 1

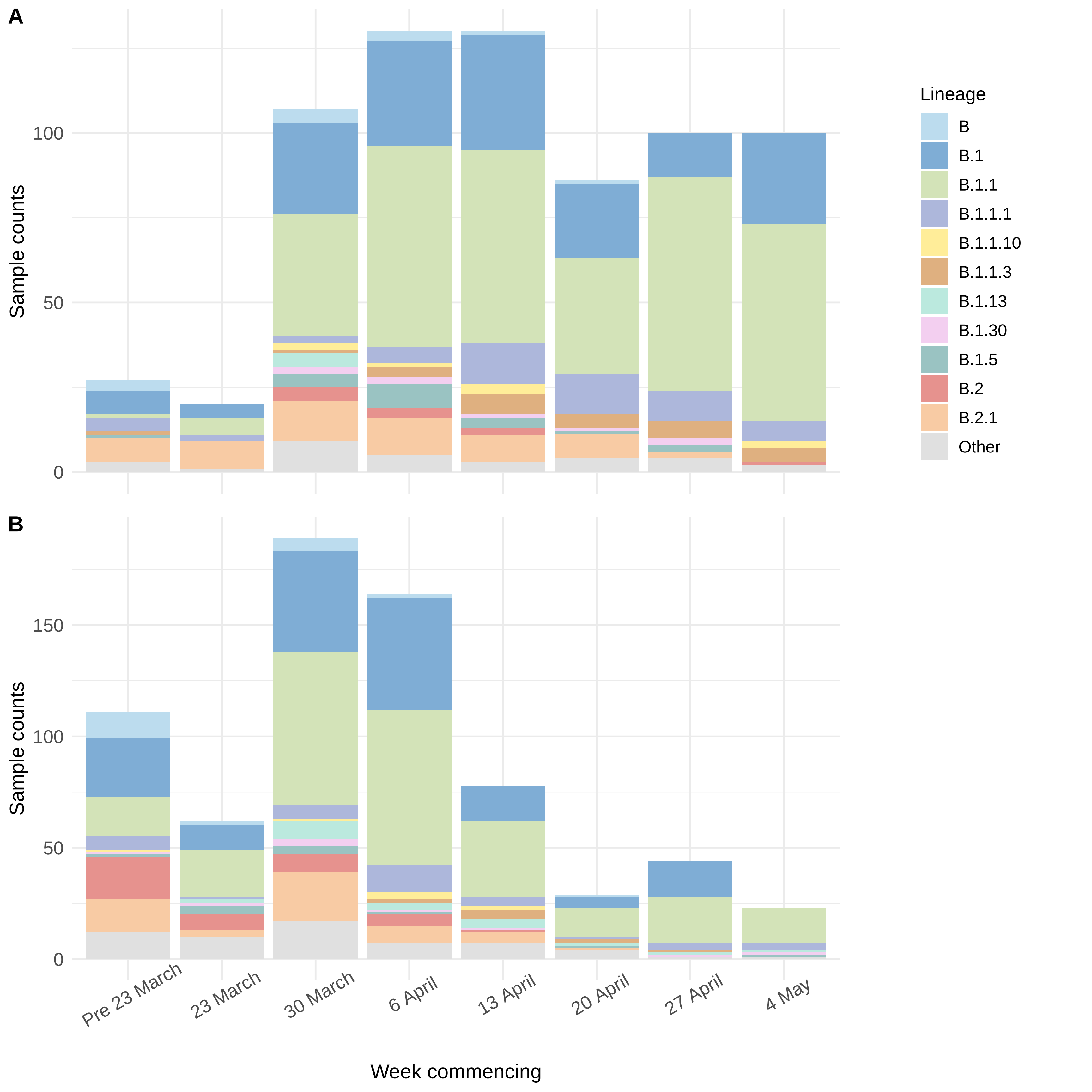

### Fig 5 Supp 2

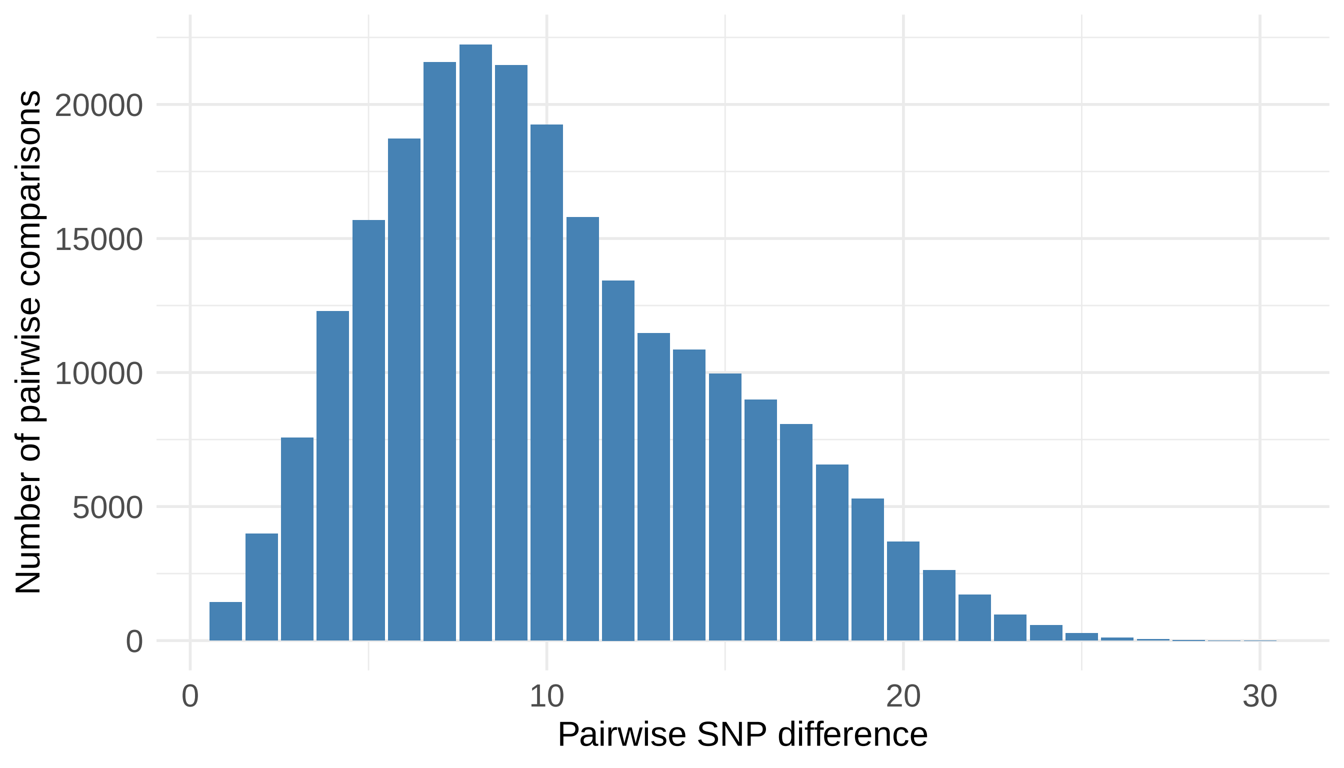

### Fig 6

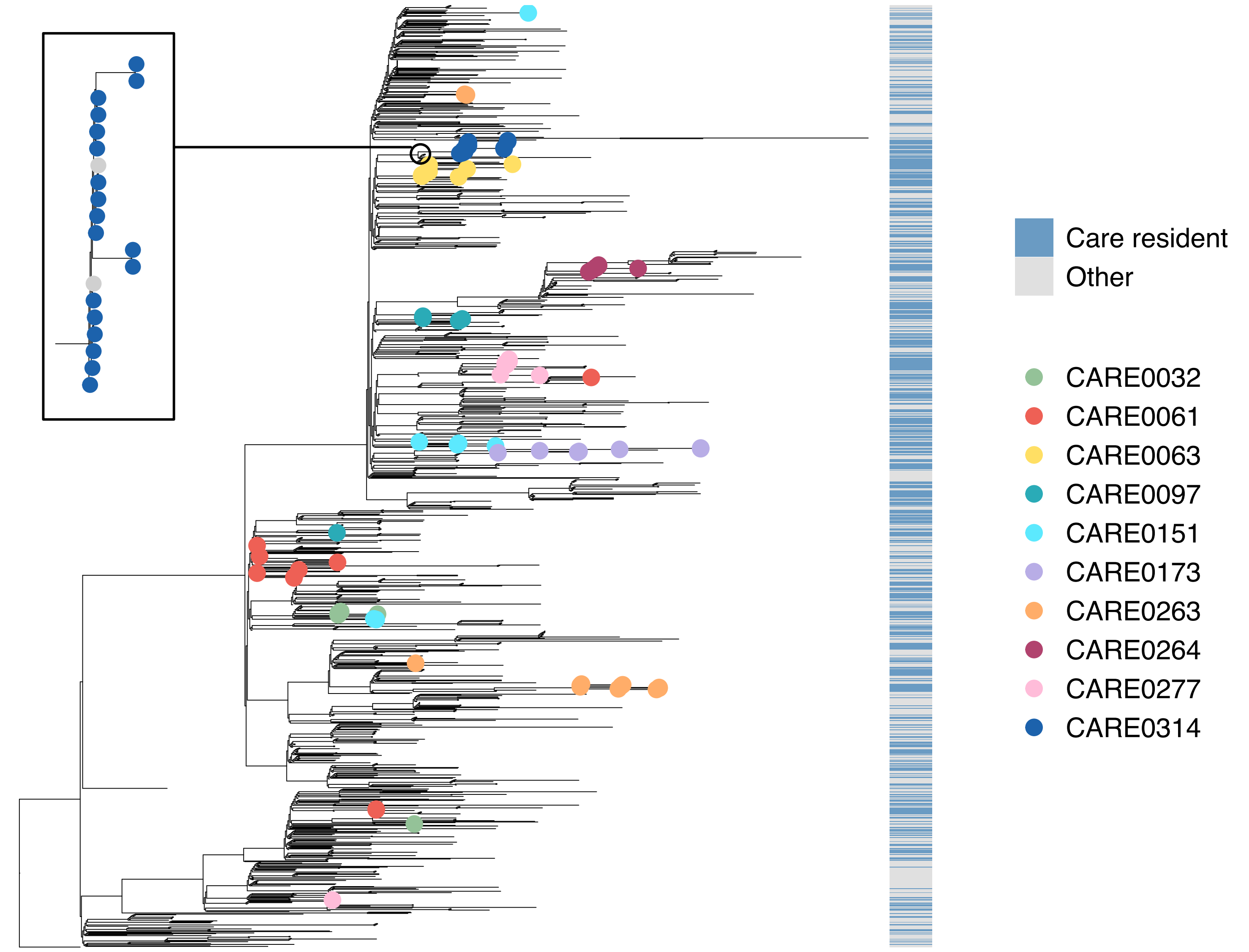

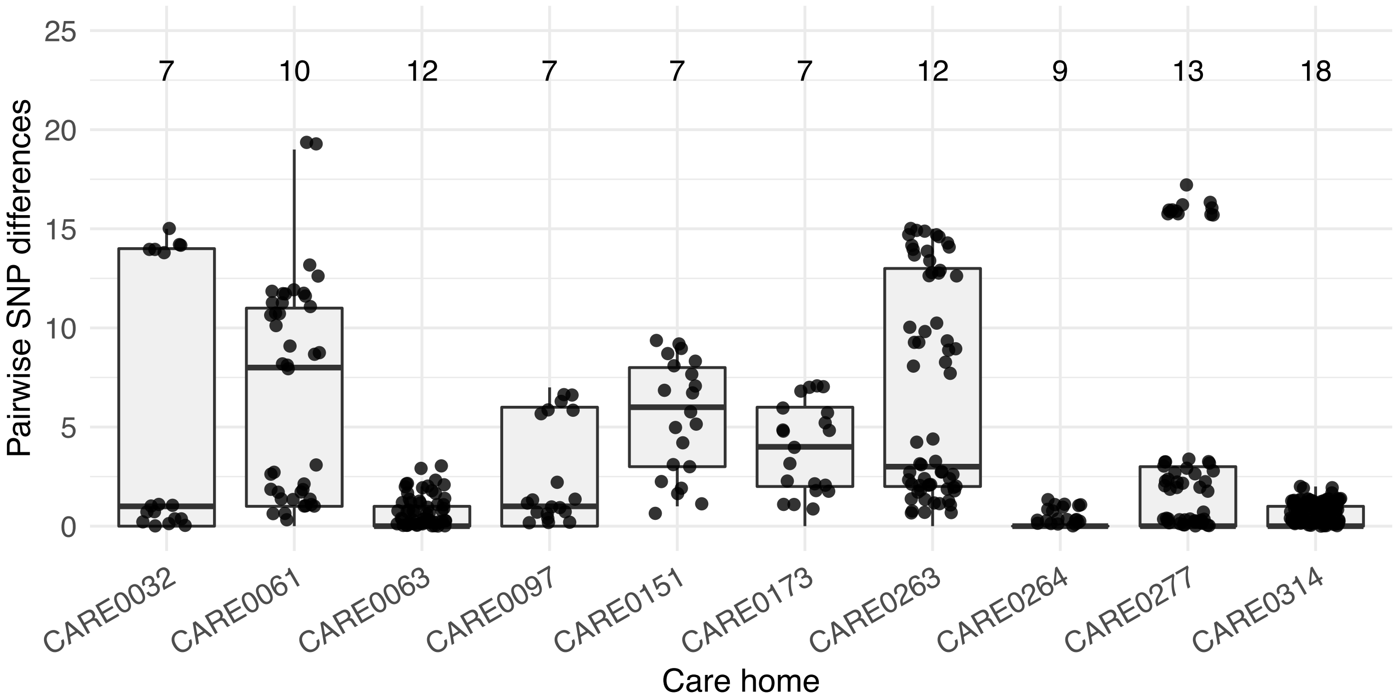

### Fig 6 Supp 1

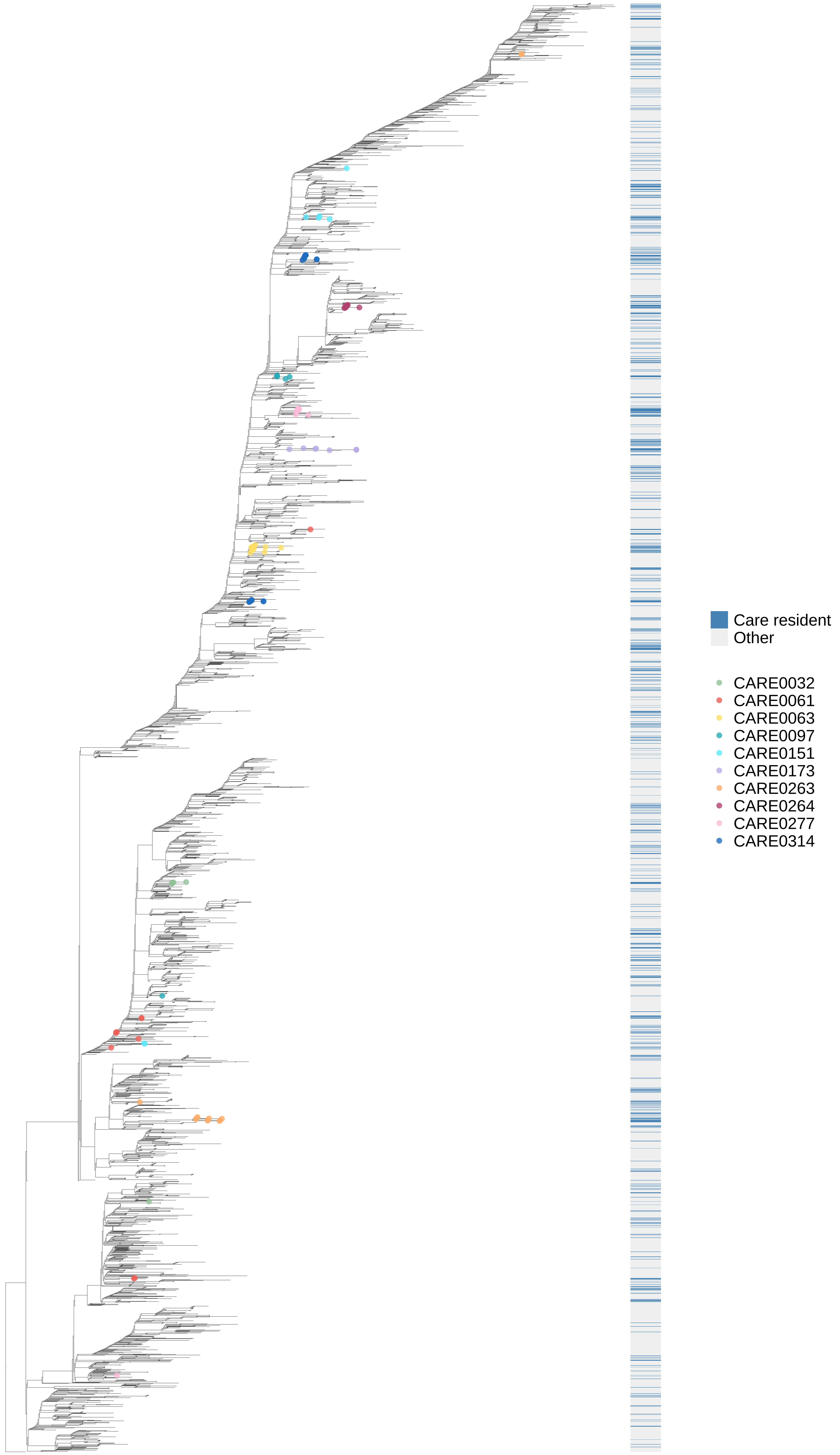

### Fig 7

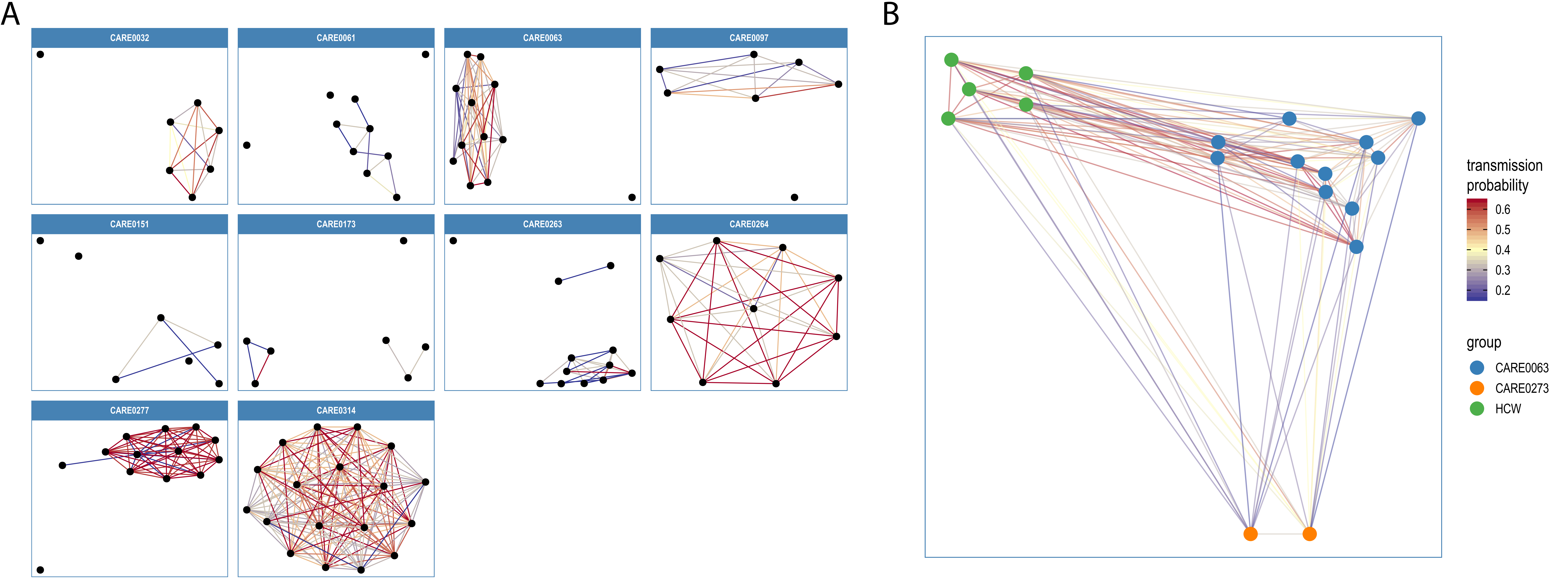

### Fig 7 Supp 1

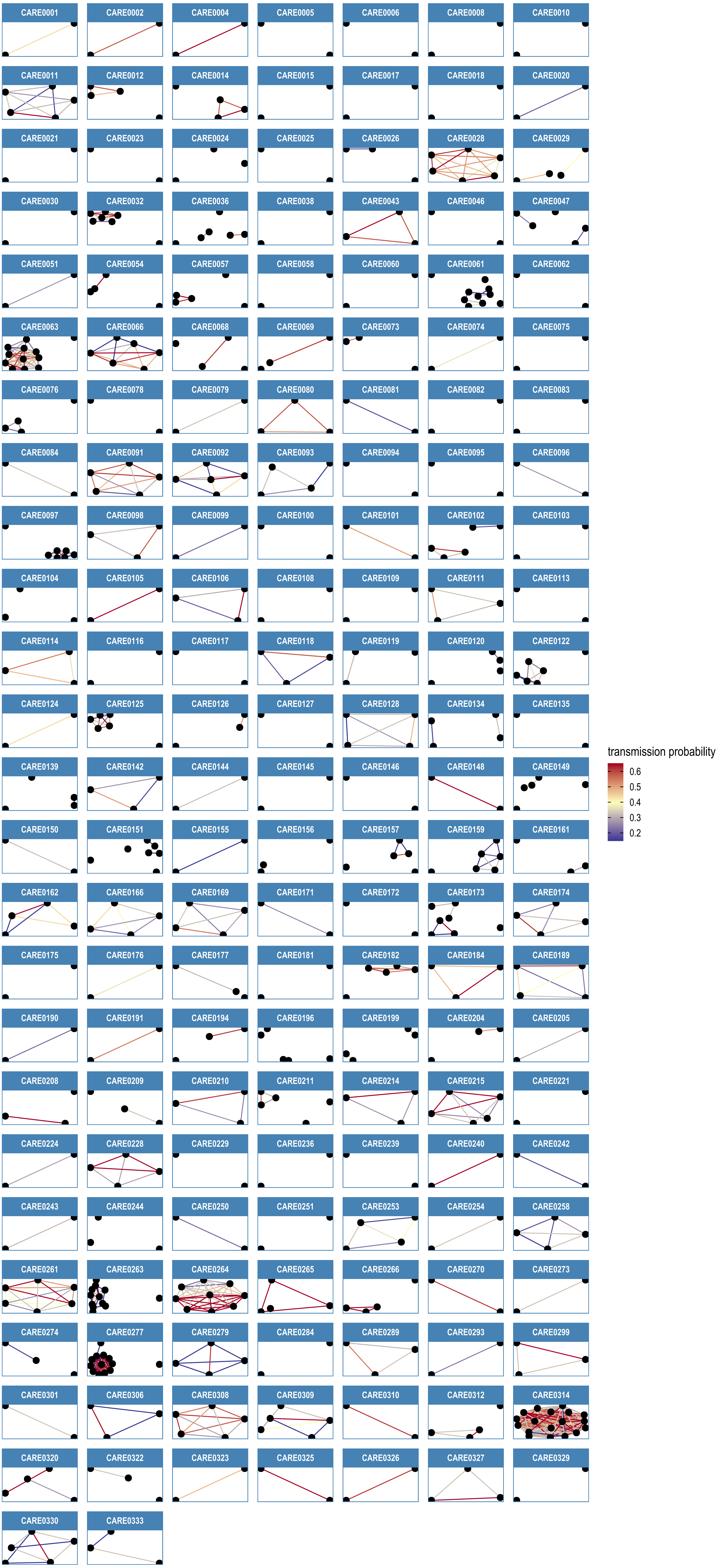

### Fig 7 Supp 2

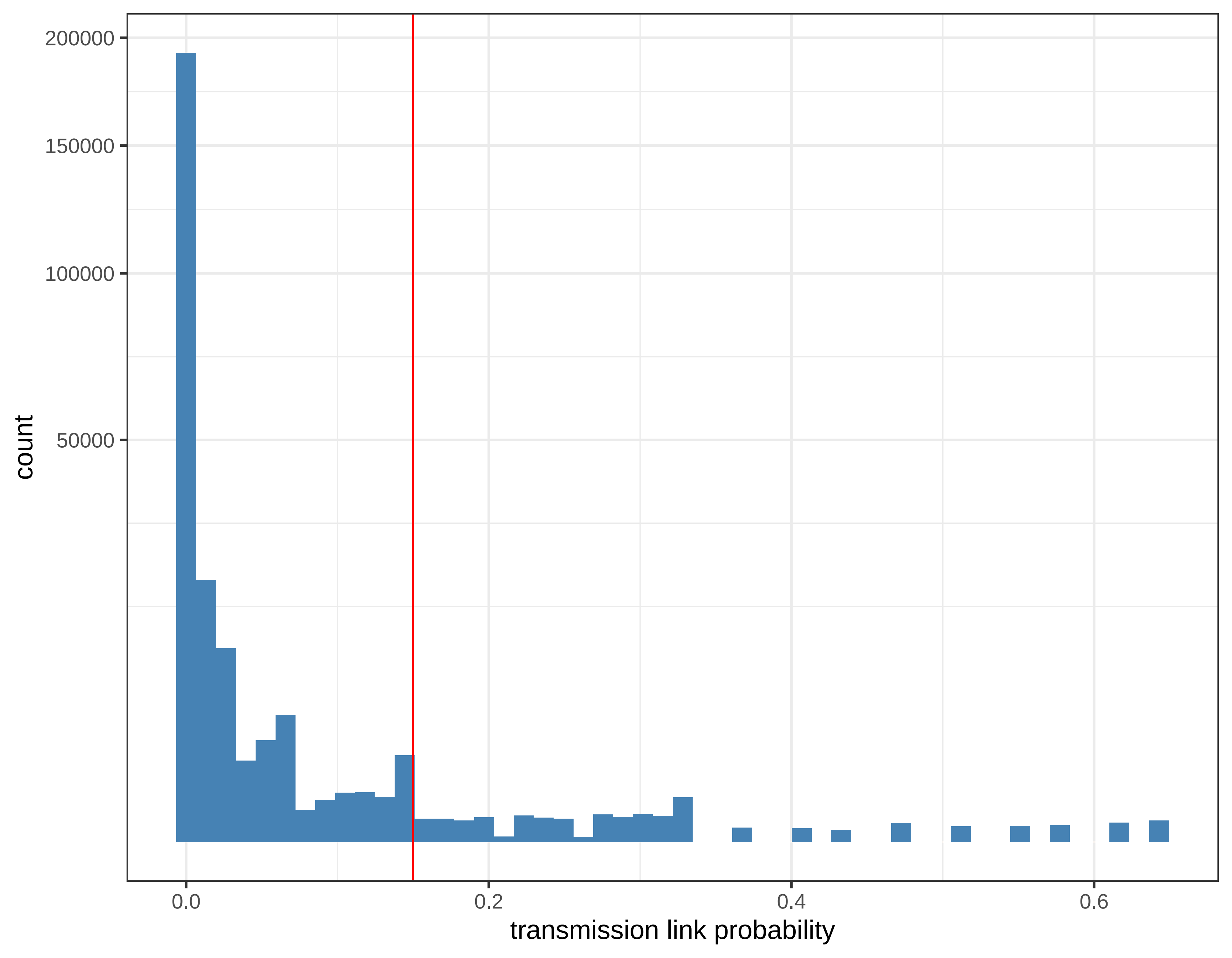

### Fig 7 Supp 3

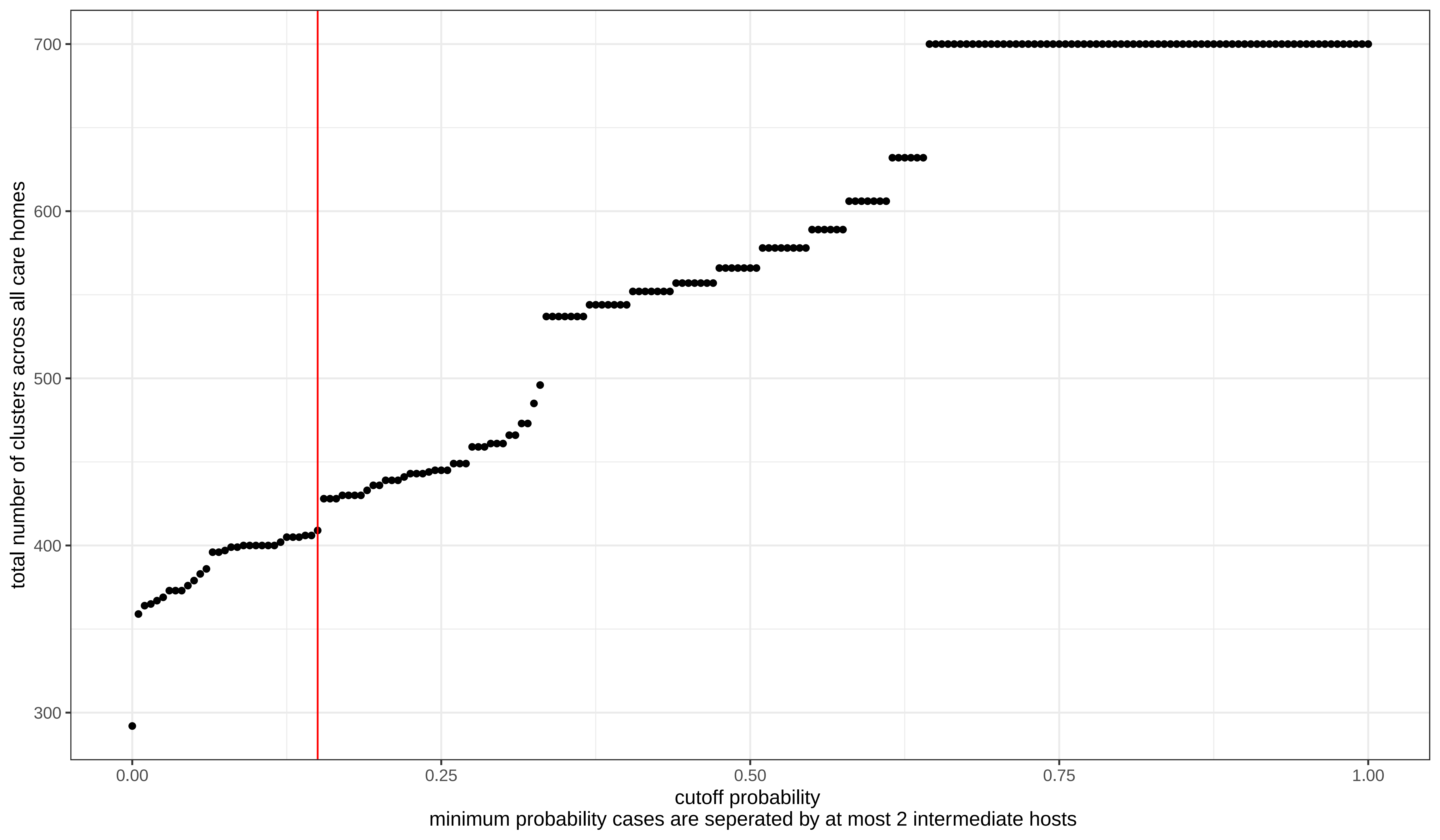

### Fig 7 Supp 4

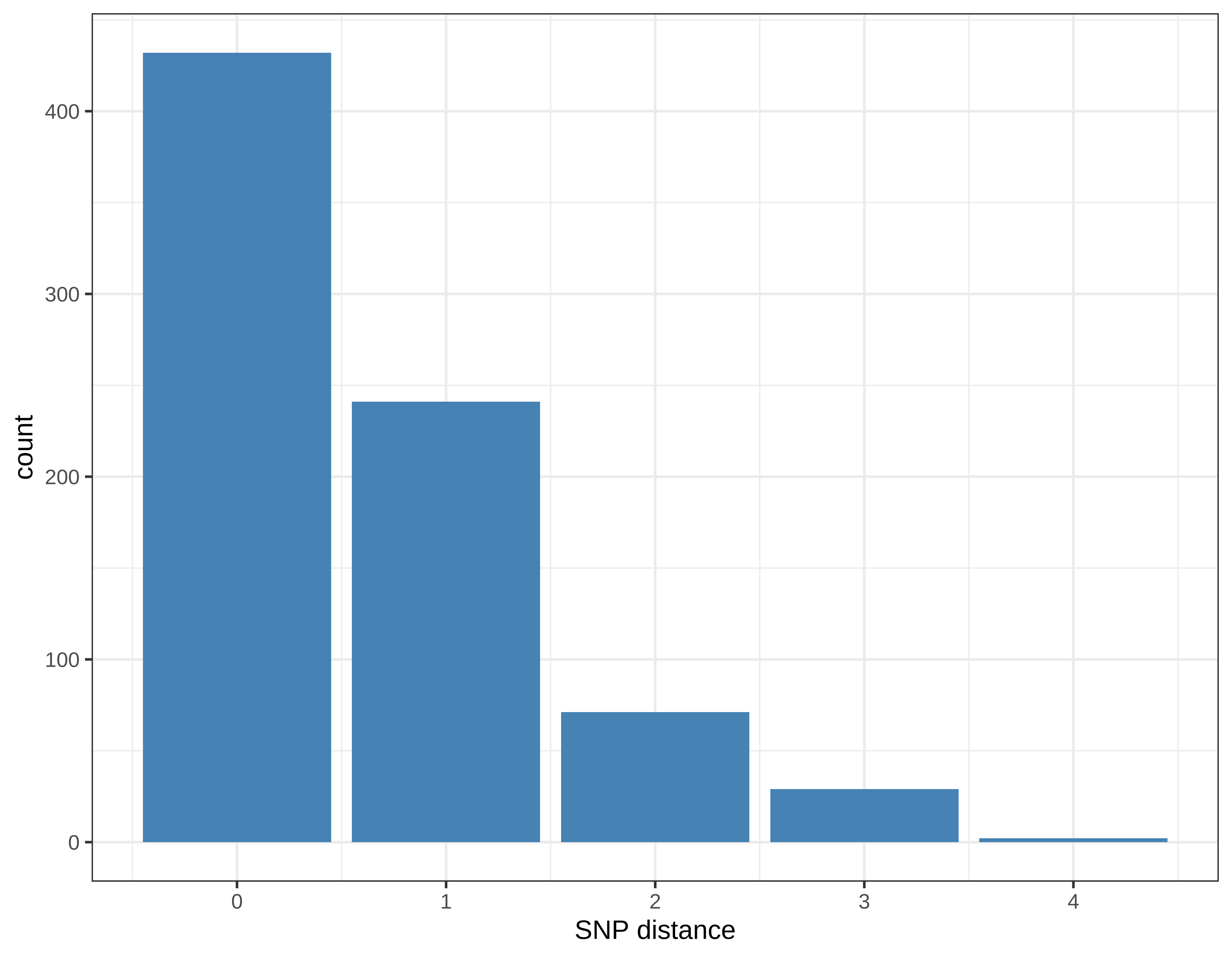

### Fig 7 Supp 5

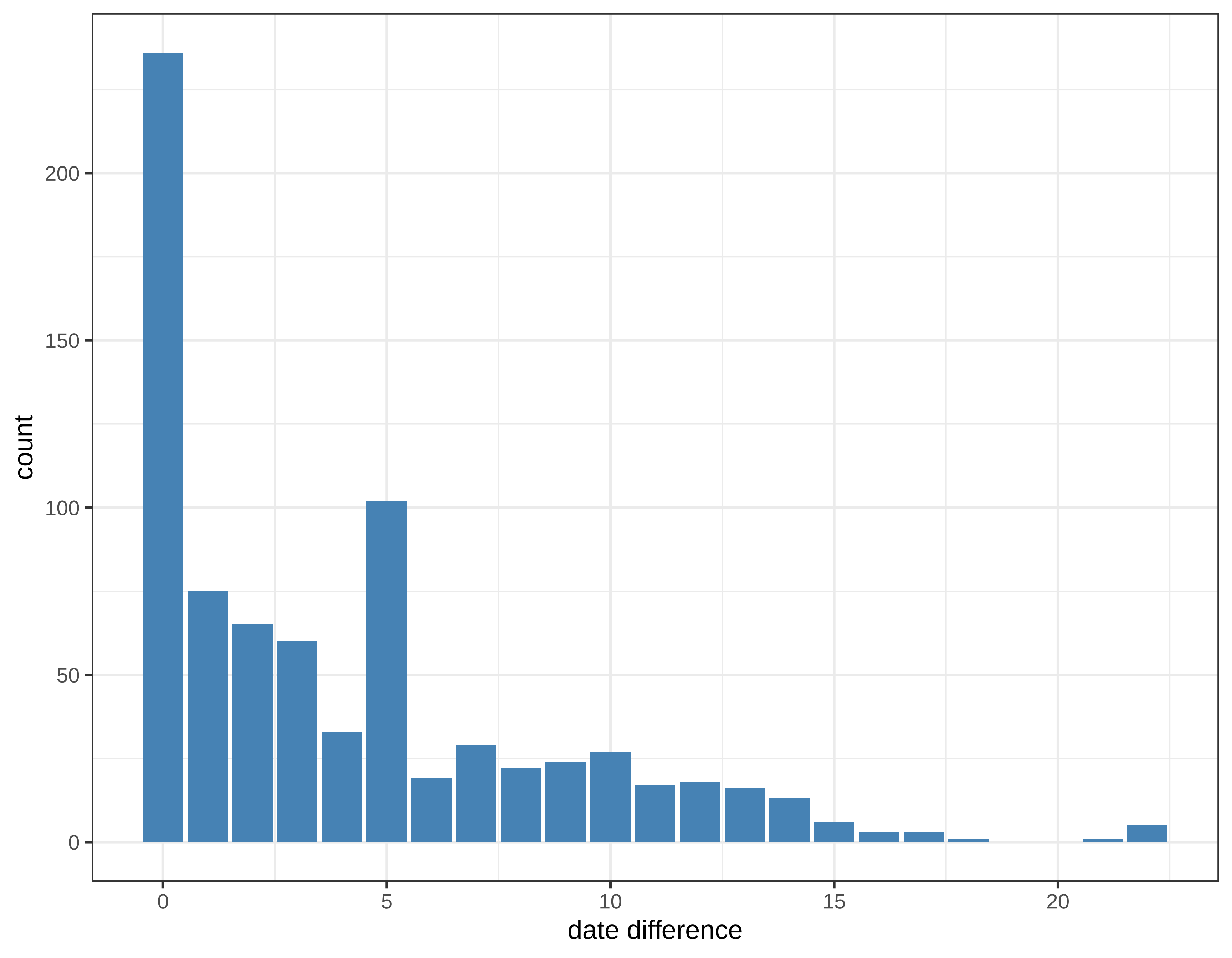

### Fig 7 Supp 6

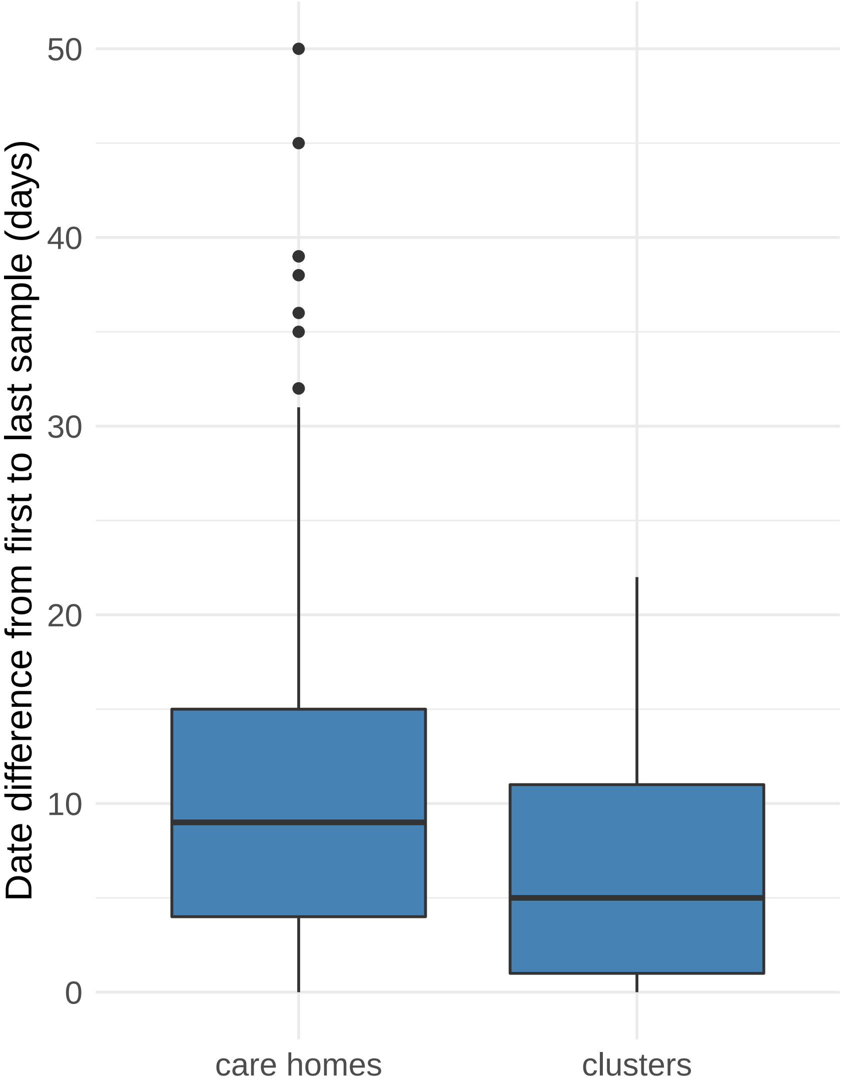

### Fig 7 Supp 7

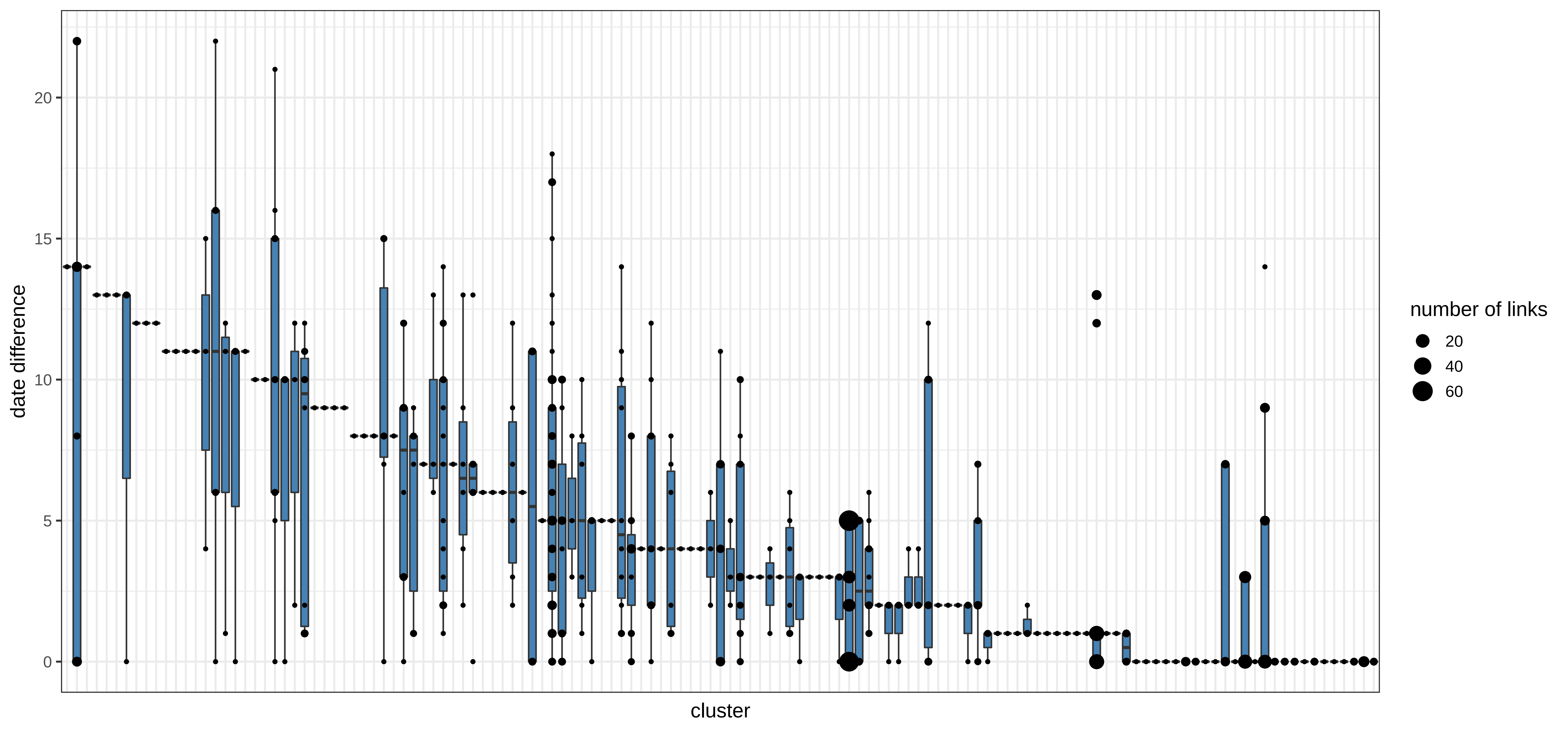
